# Unraveling the role of plasma proteins in dementia: insights from two cohort studies in the UK, with causal evidence from Mendelian randomization

**DOI:** 10.1101/2024.06.04.24308415

**Authors:** Jessica Gong, Dylan M. Williams, Shaun Scholes, Sarah Assaad, Feifei Bu, Shabina Hayat, Paola Zaninotto, Andrew Steptoe

## Abstract

Population-based proteomics offer a groundbreaking avenue to predict dementia onset. This study employed a proteome-wide, data-driven approach to investigate protein-dementia associations in 229 incident all-cause dementia (ACD) among 3,249 participants from the English Longitudinal Study of Ageing (ELSA) over a median 9.8-year follow-up, then validated in 1,506 incident ACD among 52,745 individuals from the UK Biobank (UKB) over median 13.7 years. NEFL and RPS6KB1 were robustly associated with incident ACD; MMP12 was associated with vascular dementia in ELSA. Additional markers EDA2R and KIM1 (HAVCR1) were identified from sensitivity analyses. Combining NEFL and RPS6KB1 with other factors yielded high predictive accuracy (area under the curve (AUC)=0.871) for incident ACD. Replication in the UKB confirmed associations between identified proteins with various dementia subtypes. Results from reverse Mendelian Randomization also supported the role of several proteins as early dementia biomarkers. These findings underscore proteomics’ potential in identifying novel risk screening targets for dementia.

## INTRODUCTION

The understanding of Alzheimer’s disease and related dementia (ADRD) is increasingly shifting towards a systemic and multifactorial perspective.[1, 2] Circulating proteins, as pivotal agents in biological processes, offer direct insights into disease mechanisms and can serve as early indicators, regulators, and effectors in disease pathways. This renders their studies indispensable in both drug discovery and the development of diagnostics.[3, 4]

Mounting evidence supports the significance of proteomics profiling in exploring pathways involved in ADRD.[3, 5–7] At the molecular level, deviations in protein function or expression play a role in the pathogenesis of prodromal dementia,[3, 8, 9] while protein biomarkers can forecast disease onset several years before symptoms manifest.[3, 8–10] Remarkably, approximately 96% of currently approved drugs target proteins,[4, 11] underscoring the substantial added value of proteomic analysis in ADRD drug discovery.

Integration of large-scale proteomics data into population studies represents a recent development,[3, 12] enabling cost-effective simultaneous measurement of multiple proteins on many samples.[3, 13, 14] This has led to the identification of distinct protein signatures relevant to ADRD susceptibility.[4, 9, 10, 15, 16] For instance, longitudinal analyses in the UK Whitehall II study spanning across two decades, demonstrated associations between 15 non-amyloid/non-tau-related proteins and cognitive decline and dementia.[4] Similarly, the Atherosclerosis Risk in Communities (ARIC) study in the US identified significant protein signatures for dementia, including immune and proteostasis/autophagy pathways.[10] Intriguingly, some of these associations were independent of known Alzheimer’s disease (AD) risk factors, suggesting novel potential targets for intervention.[4, 9, 10] Recent analyses based on data from the UK Biobank, which identified GFAP, NEFL, GDF15 and LTBP2 were most strongly associated with incident all-cause dementia (ACD), AD, and vascular dementia (VAD).[16] However, previous studies utilized the aptamer-based SomaScan platform in ARIC and Whitehall II,[4, 9, 10] and the SomaScan platform is deemed to have lower specificity compared to the Olink platform, which employs multiplexed antibody-based immunoassays proximity extension assay (PEA) technology.[17] Moreover, the UK Biobank study faced limitations such as a lack of external validation cohorts and evidence triangulation with causal inference.[16]

In this current study, we employed the large-scale Olink proteomics platform and a robust dementia algorithm to assess the proteomic signature of dementia risk in over 3,000 older adults using data from the English Longitudinal Study of Ageing (ELSA) as the discovery cohort. We validated these findings using Olink proteomics data from over 50,000 participants from the UK Biobank.[18] Two-sample bi-directional Mendelian randomization (MR) and drug target MR (or cis-MR) were utilized to infer causality between protein concentration and dementia outcomes, leveraging summary statistics from large genome-wide association study (GWAS) consortia.

## RESULTS

### Analysis 1: Protein-dementia associations in the ELSA discovery cohort

The participant selection for the proteomics assay in ELSA is depicted in Supplementary Fig. 1. In 3,262 samples with proteomics assayed, based on the dementia algorithm, prevalent dementia cases were excluded (N = 13), resulting in a final sample of 3,249 in the analysis. The mean age was 63.4 years (SD = 9.2), 55% were women, and 97.2% were of white ethnicity (Supplementary Table 1). A total of 229 incident dementia cases were documented over a median follow-up of 9.8 years (min – max: 0.4 – 10.9 years). Specific details on the data sources where these cases were extracted from in the first instance are included in Supplementary Fig. 2. The normalized protein expression (NPX) levels for participants with no dementia, incident dementia, and prevalent dementia for each protein are presented as box plots in Supplementary Fig. 3.

We initially assessed the relationship between the NPX value of 276 plasma proteins and ACD risk in the ELSA cohort, using Cox Proportional Hazard regression models. Unadjusted analyses revealed that 95 measured proteins were significantly associated with ACD (Supplementary Fig. 4). Among these, **NEFL** exhibited the strongest association with ACD, as indicated by the false discovery rate (FDR)-corrected p-value (denoted as P_FDR_ = 8.66 × 10^-37^, hazard ratio (HR) [95% confidence intervals (CI)]: **3.01 [2.63, 3.44]**)), followed by **EDA2R, SCARF2, LAYN, PGF, DCN, GFR-alpha-1, BNP, UNC5C, Dkk-4, KIM1** (also known as HAVCR1)**, TNFRSF12A, CADM3, TRAIL-R2, VWC2**, and **MMP12.**

In the minimally adjusted models (adjusted for age, sex, and ethnicity), **NEFL** (P_FDR_ = 0.0002; HR [95% CI]: **1.55 [1.30, 1.85]**), **RPS6KB1** (P_FDR_ = 0.003; HR [95% CI]: **1.34 [1.18, 1.53]**), **EDA2R** (P_FDR_ = 0.046; HR [95% CI]: **1.43 [1.19, 1.72]**) and **KIM1** (P_FDR_ = 0.049; HR [95% CI]: **1.31 [1.14, 1.50]**) were significantly associated with ACD (Supplementary Fig. 5).

In the fully adjusted models (adjusted for age, sex, ethnicity, education, smoking status, depression, presence of cardiovascular diseases, body mass index (BMI), systolic blood pressure, low-density lipoprotein (LDL) cholesterol), **NEFL** (P_FDR_ = 0.0008; HR [95% CI]: **1.54 [1.29, 1.84]**) and **RPS6KB1** (P_FDR_ = 0.01; HR [95% CI]: **1.33 [1.16, 1.52]**) remained significantly associated with ACD (Fig. 1).

**Figure 1.**
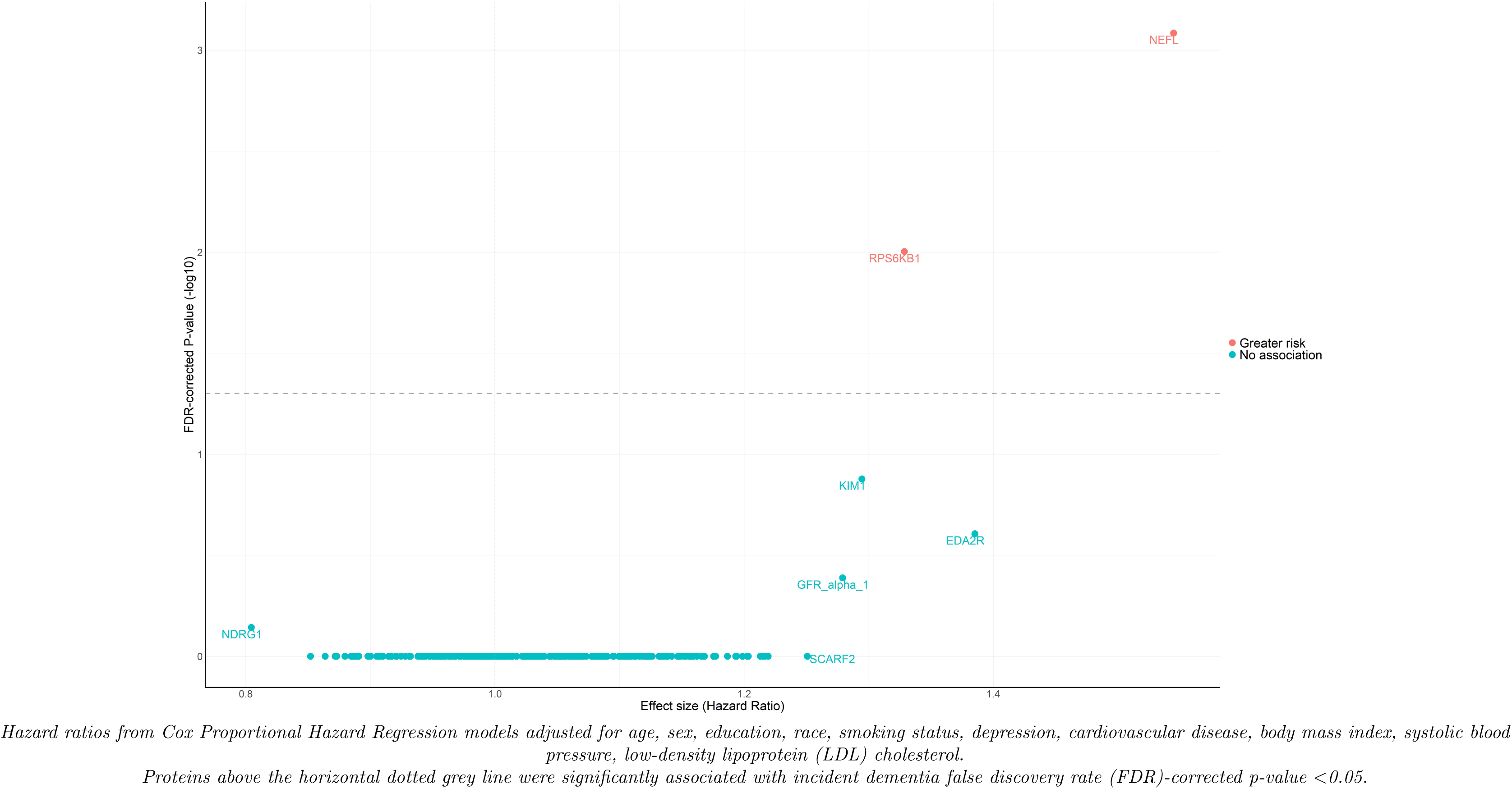
Volcano plot showing the Hazard ratio (x-axis) and two-sided false discovery rate (FDR)-corrected P values (y-axis) for the association of protein concentration with incident all-cause dementia using imputed data.

Sensitivity analyses demonstrated the robustness of the association between **NEFL** and **ACD**, with significance persisting after excluding participants in other ethnic groups other than white participants (P_FDR_ = 0.001; HR [95% CI]: **1.53 [1.28, 1.83]**) (Supplementary Fig. 6), APOE ε4 carriers (P_FDR_ = 0.009; HR [95% CI]: **1.61 [1.29, 2.01]**) (Supplementary Fig. 7), cases of dementia occurring within the first year of follow-up (P_FDR_ = 0.005; HR [95% CI]: **1.50 [1.25, 1.80]**) (Supplementary Fig. 8), participants aged <60 years (P_FDR_ = 0.011; HR [95% CI]: **1.47 [1.22, 1.78]**) (Supplementary Fig. 9), and when death was considered as a competing risk in Fine-Gray regression models (P_FDR_ = 0.002; HR [95% CI]: **1.50 [1.26, 1.80]**) (Supplementary Fig. 10). Similarly, **RPS6KB1** exhibited a robust association with **ACD**, which remained significant after excluding other ethnic groups (P_FDR_ = 0.004; HR [95% CI]: **1.34 [1.18, 1.53]**), cases of dementia occurring within the first year of follow-up (P_FDR_ = 0.025; HR [95% CI]: **1.31 [1.15, 1.50]**), participants aged <60 years (P_FDR_ = 0.031; HR [95% CI]: **1.31 [1.14, 1.50]**), and it was significantly associated with ACD in the Fine-Gray competing risk model (P_FDR_ = 0.014; HR [95% CI]: **1.33 [1.16, 1.52]**). However, the significance in association between RPS6KB1 and ACD based on P_FDR_ attenuated after excluding APOE ε4 carriers (P_FDR_ = 0.961; HR [95% CI]: **1.28 [1.08, 1.50]**). After excluding other ethnic groups, **KIM1** was additionally associated with **ACD** (P_FDR_ = 0.027; HR [95% CI]: **1.32 [1.15, 1.53]**). No evidence was found for any sex differences in the protein-dementia associations when sex was fitted as an interaction term in the Cox models (all P_FDR_ for interaction > 0.05).

When assessed by dementia subtypes, 89 incident AD and 41 cases of VAD were documented. After full adjustment, no protein was found to be significantly associated with AD indicated by P_FDR_ < 0.05 (Supplementary Fig. 11). **MMP12** was found to be associated with **VAD** (P_FDR_ = 0.046; HR [95% CI]: **2.06 [1.41, 2.99]**) (Supplementary Fig. 12). Albeit being non-significant after FDR correction, of the proteins significantly associated with ACD, based on nominal statistical significance (denoted as P_uncorrected_ < 0.05), **RPS6KB1** was associated with **AD** (P_uncorrected_ = 0.006; HR [95% CI]: **1.29 [1.07, 1.55]**); and **NEFL** was associated with **VAD** (P_uncorrected_ = 0.001; HR [95% CI]: **1.98 [1.31, 2.99]**).

For predicting incident ACD, plasma NEFL and RPS6KB1 parsimonious models yielded modest Area Under the Receiver Operating Characteristic Curve (AUC) values [95% CI] of 0.787 [0.757, 0.815], and 0.609 [0.571, 0.647], respectively (Fig. 2). We also evaluated the performance of these two plasma proteins in combination with other measures, including demographic predictors (age, sex, ethnicity, education), APOE ε4 status, and memory score (a combined test score of immediate– and delayed-recall). When NEFL was combined with these predictors, the model achieved an accuracy of AUC [95% CI] = 0.866 [0.840, 0.888]. Comparatively, when RPS6KB1 was combined with other predictors, the model achieved a comparable accuracy of AUC [95% CI] = 0.866 [0.842, 0.891]. NEFL and RPS6KB1 in combination with all the other predictors yielded AUC [95% CI] = 0.871 [0.845, 0.894]. The XGBoost models revealed that age (mean |SHAP| = 0.047) and memory score (mean |SHAP| = 0.017) were the most important features contributing to the prediction ACD onset. Additionally, the protein markers RPS6KB1 (mean |SHAP| = 0.009) and NEFL (mean |SHAP| = 0.008) emerged as the most prominent protein markers in predicting ACD (Fig. 3). The SHAP plot also illustrated that individuals with elevated levels of RPS6KB1 were more predisposed to developing ACD, while those with lower levels were more likely to remain ACD-free.

**Figure 2.**
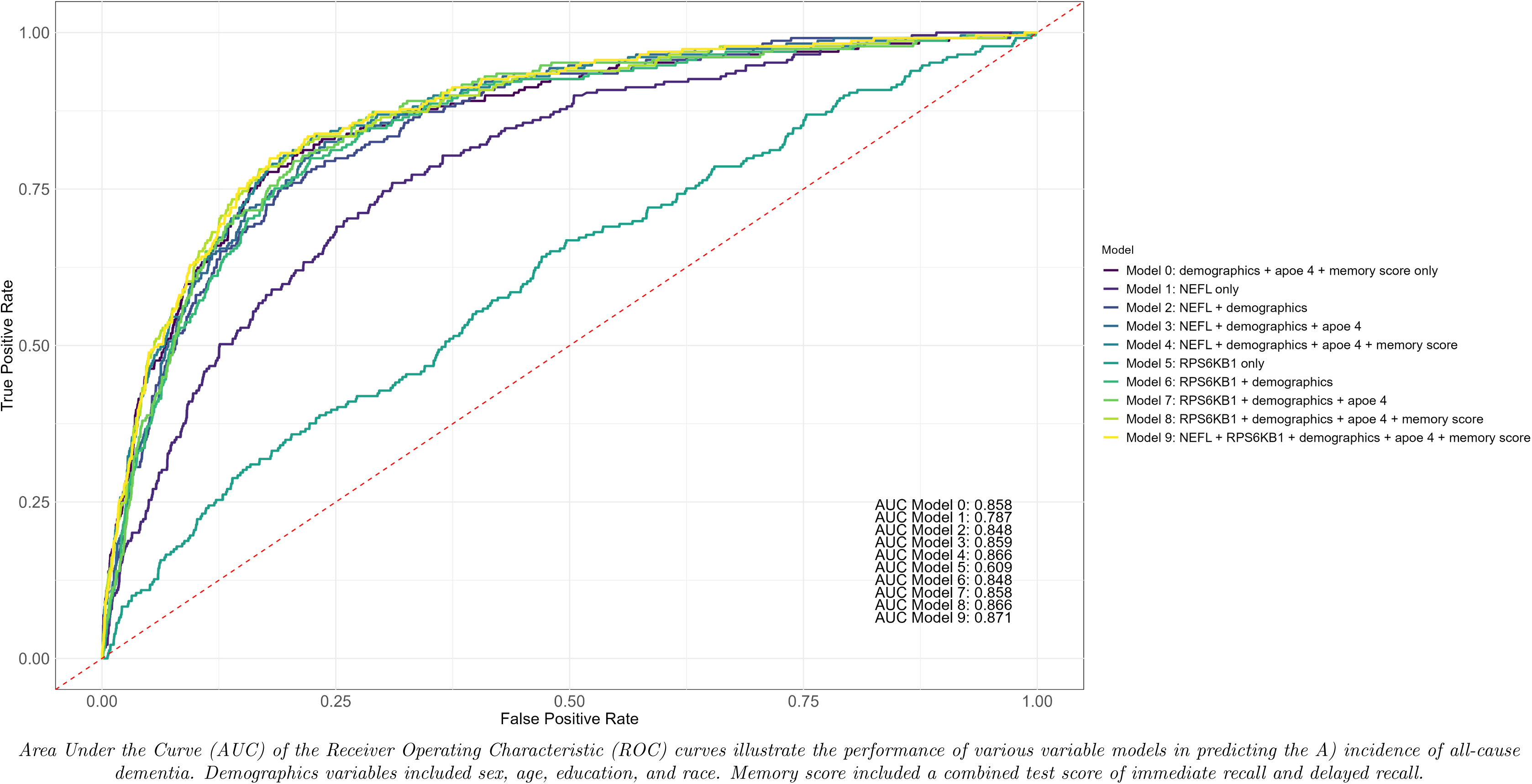
Predictive accuracy of NEFL and RPS6KB1, alone or in combination with demographic variables, APOE 4 status, and memory score for all-cause dementia.

**Figure 3.**
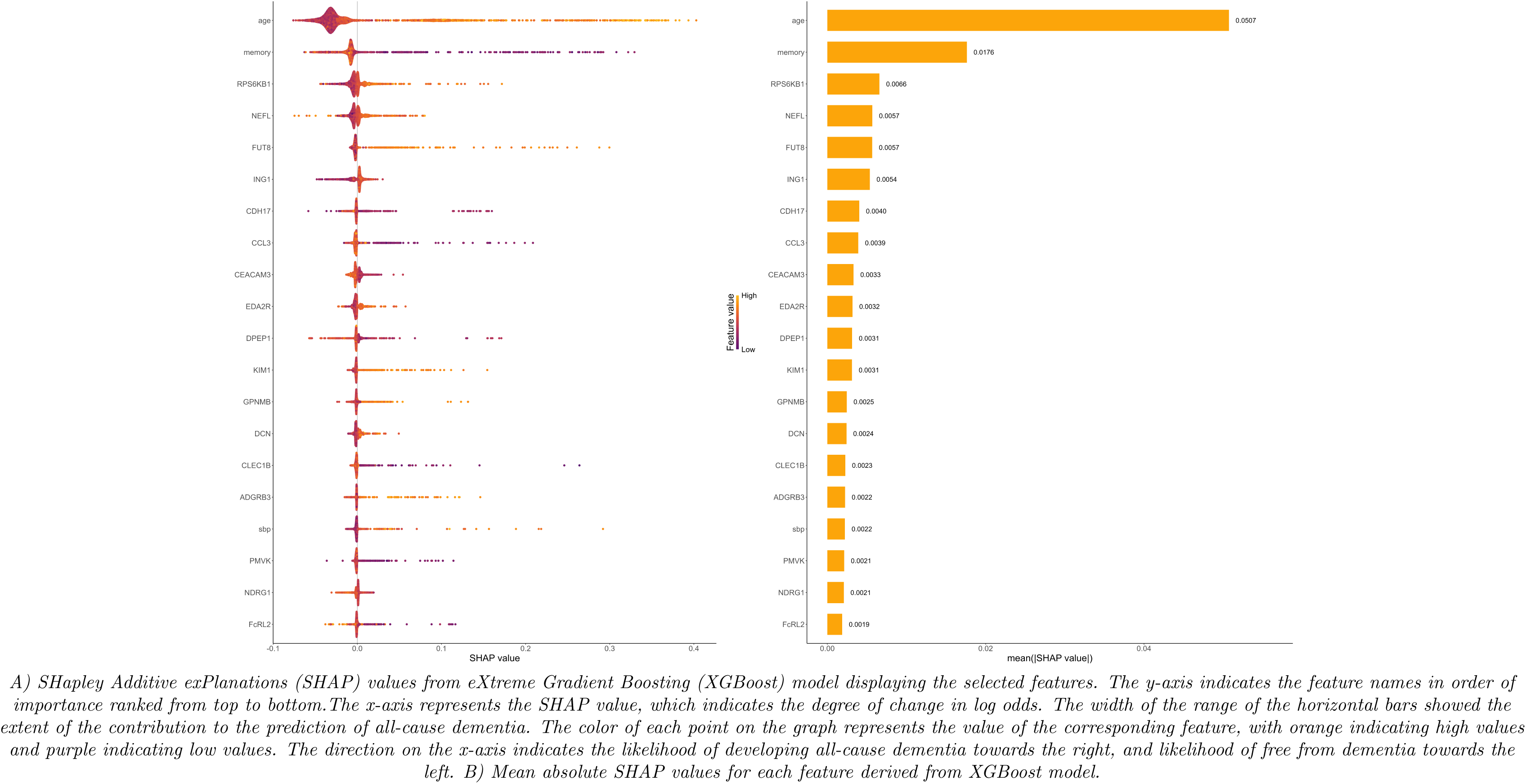
Protein importance ranking using XGBoost decision tree-based machine learning algorithm and SHAP visualization for selected features on all-cause dementia.

### Analysis 2: Protein-dementia associations in the UK Biobank validation cohort

Based on the results from the main, sensitivity, and dementia subtype analyses conducted in ELSA discovery cohort, all identified proteins were selected for further validation analyses using proteomics data from UK Biobank validation cohort. However, RPS6KB1 was not assayed in the UK Biobank.

In the UK Biobank, which included 52,745 participants with proteomics assayed and without dementia at study baseline (53.9% women, 93.3% white ethnicity), the mean age was 56.8 years (SD = 8.2) (Supplementary Table 1). UK Biobank participants with proteomics assayed were, on average, younger than participants in ELSA. Over a median of 13.7 years (min – max: 0.03 – 16.8 years) of follow-up, a total of 1,506 incident ACD, 732 incident AD, 281 incident VAD, and 111 incident frontotemporal dementia (FTD) cases were recorded.

Using the same adjustment strategy for the Cox regression models in ELSA, **NEFL** was replicated in the UK Biobank for **ACD** (P_FDR_ = 1.02 × 10^-81^; HR [95% CI]: **1.87 [1.75, 1.99]**). NEFL was also associated with **AD** (P_FDR_ =1.89 × 10^-35^; HR [95% CI]: **1.81 [1.65, 1.99]**), **VAD** (P_FDR_ = 1.59 × 10^-17^; HR [95% CI]: **1.90 [1.64, 2.19]**), and **FTD** (P_FDR_ = 1.10 × 10^-21^; HR [95% CI]: **2.97 [2.30, 3.70]**) (Fig. 4; Supplementary Table 2). **KIM1** was replicated for **ACD** (P_FDR_ =3.15 × 10^-4^; HR [95% CI]: **1.13 [1.06, 1.20]**); and was also associated with **AD** (P_FDR_ = 0.077; HR [95% CI]: **1.11 [1.02, 1.21]**), and **VAD** (P_FDR_ = 1.13 × 10^-6^; HR [95% CI]: **1.44 [1.25, 1.66]**). **MMP12** was replicated for **VAD** (P_FDR_ = 6.85 × 10^-5^; HR [95% CI]: **1.36 [1.18, 1.56]**) and it was also significantly associated with **ACD** (P_FDR_ = 2.00 × 10^-6^; HR [95% CI]: **1.17 [1.10, 1.24]**). **EDA2R** was replicated for **ACD** (P_FDR_ = 3.18 × 10^-13^; HR [95% CI]: **1.31 [1.22, 1.40]**); and was also associated with **AD** (P_FDR_ = 6.06 × 10^-5^; HR [95% CI]: **1.25 [1.13, 1.39]**), and with **VAD** (P_FDR_ = 0.001; HR [95% CI]: **1.34 [1.15, 1.58]**).

**Figure 4.**
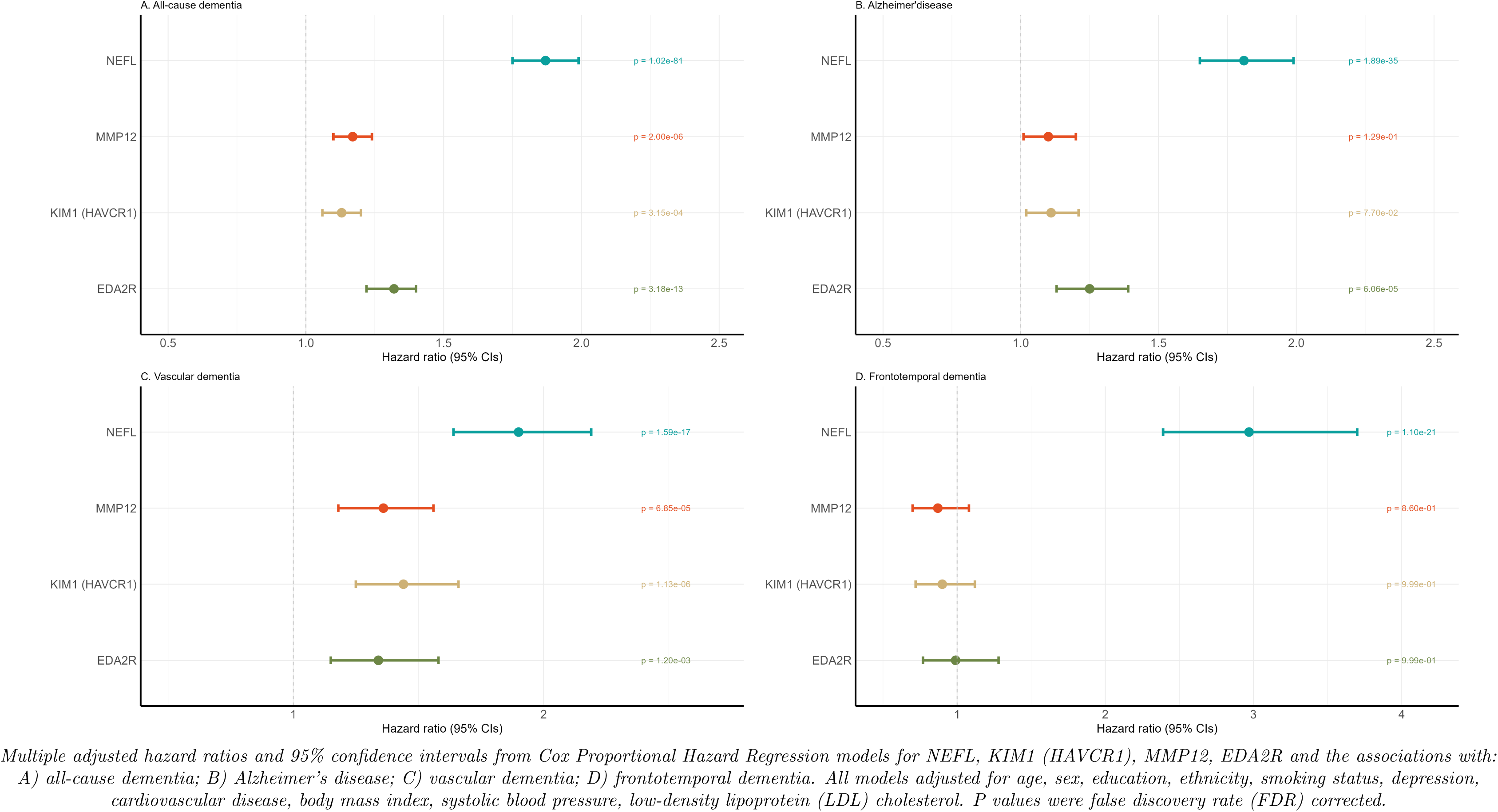
Forest Plots for the associations between identified proteins from ELSA and dementia and dementia subtypes validated in the UK Biobank.

### Analysis 3: Two-sample bi-directional MR

We then assessed the potential causal relationships between the circulating protein concentration, identified as significant based on the analyses described above, in relation to dementia outcomes, using two-sample bi-directional MR. Summary statistics for genetic variants associated with the circulating protein levels, protein quantitative trait locus (pQTL), that is also associated with dementia in GWAS were used to infer causality. Since RPS6KB1 was not assayed in the UK Biobank, MR analyses were therefore conducted for NEFL, KIM1, EDA2R, and MMP12.

For the GWAS used for dementia outcomes, three separate GWAS for AD (denoted as Kunkle 2019, Bellenguez 2022, FinnGen 2023), one GWAS for ACD (FinnGen 2023), and one GWAS for VAD (FinnGen 2023) were used.

In the forward direction MR (circulating protein concentration → dementia), there was evidence of a potential causal link from circulating **EDA2R** to **AD** (FinnGen 2023) (coefficient (β) [standard error (se)] = 0.259 [0.096], P = 0.007, based on the inverse-variance weighted (IVW) method), and **ACD** (β [se] = 0.232 [0.110], P = 0.035, based on IVW) (Fig. 5, Supplementary Table 3). However, these findings were less robust when methods such as MR-Egger were applied, suggesting the presence of horizontal pleiotropy.

**Figure 5.**
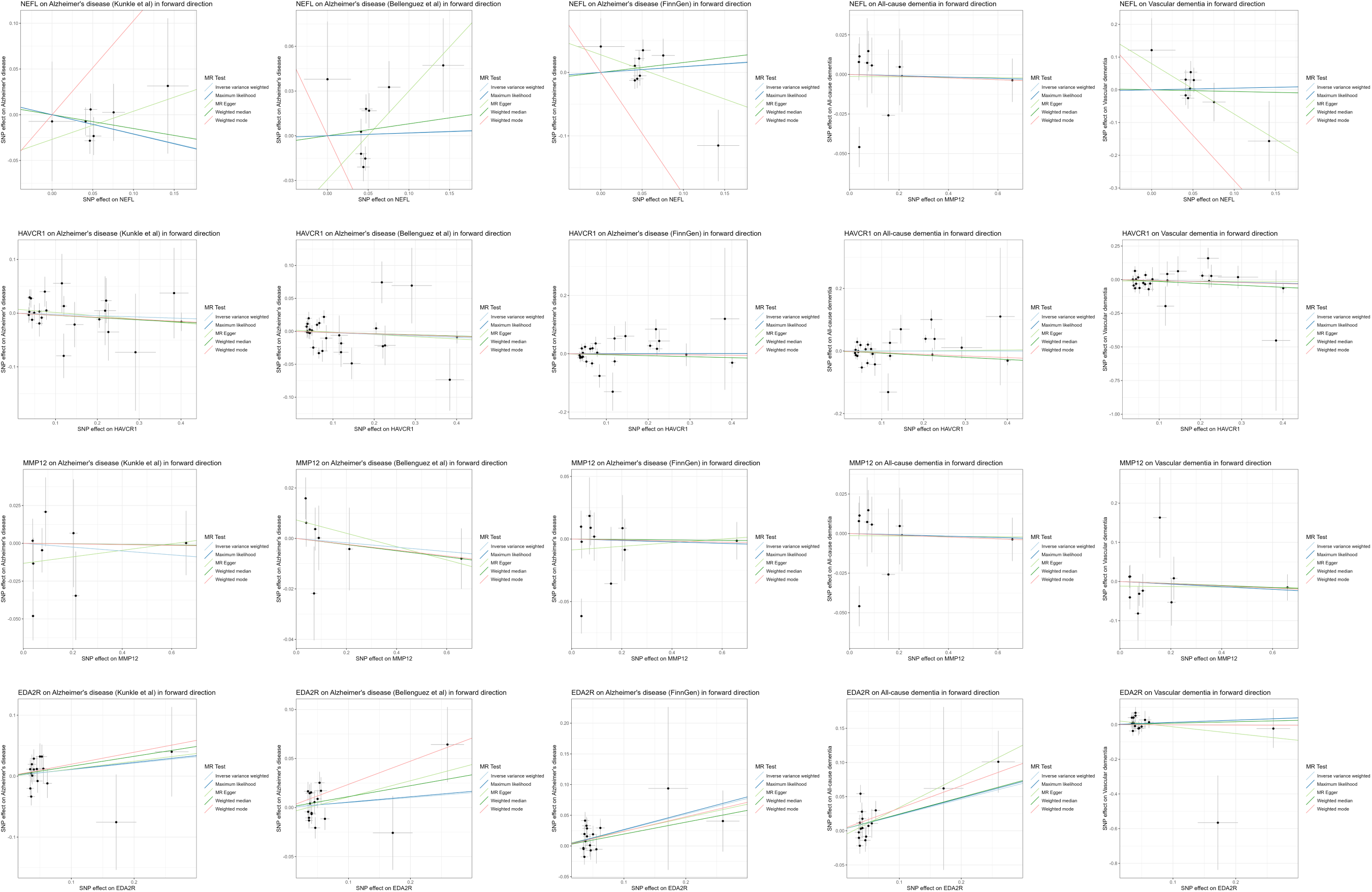
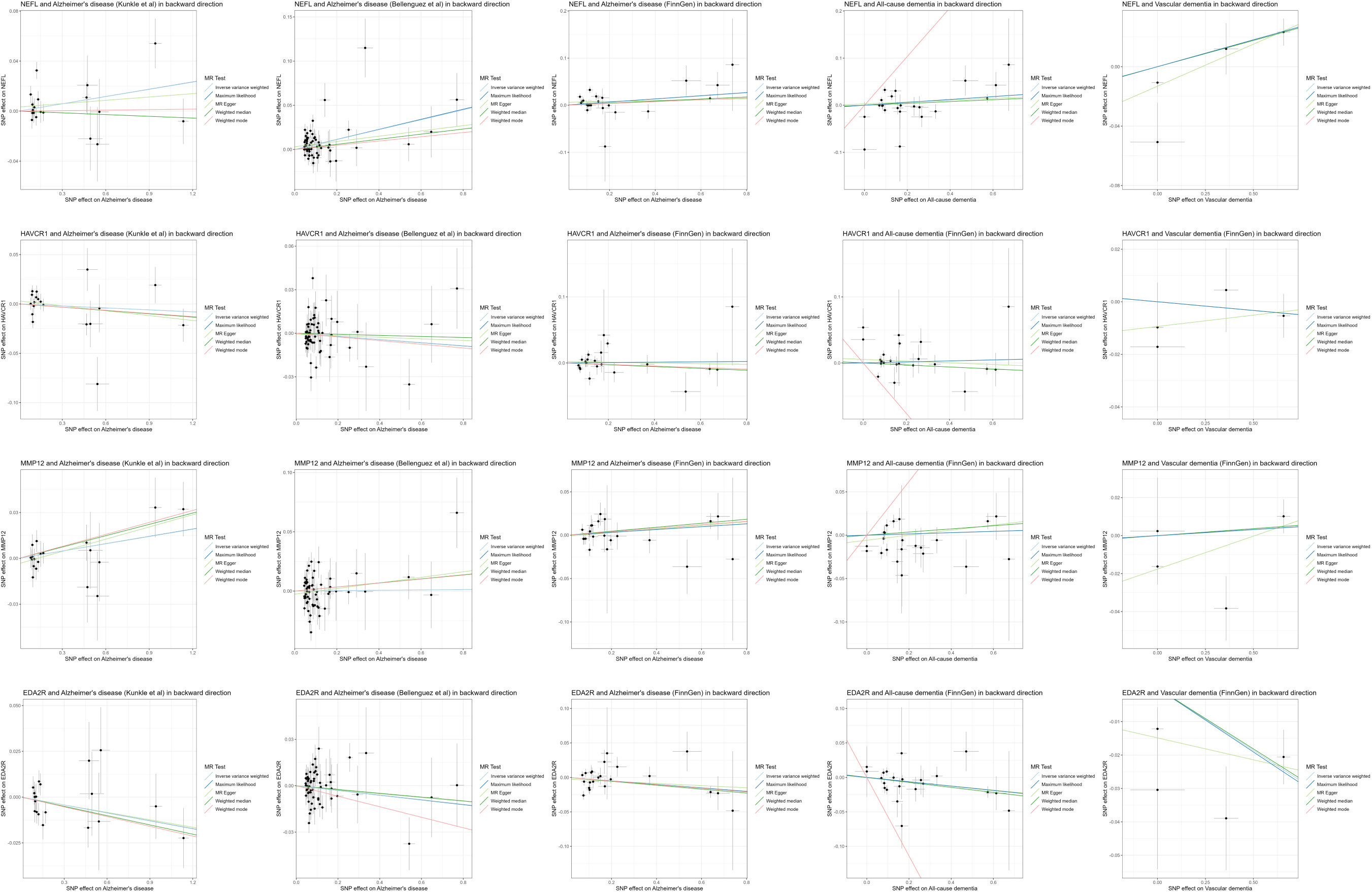
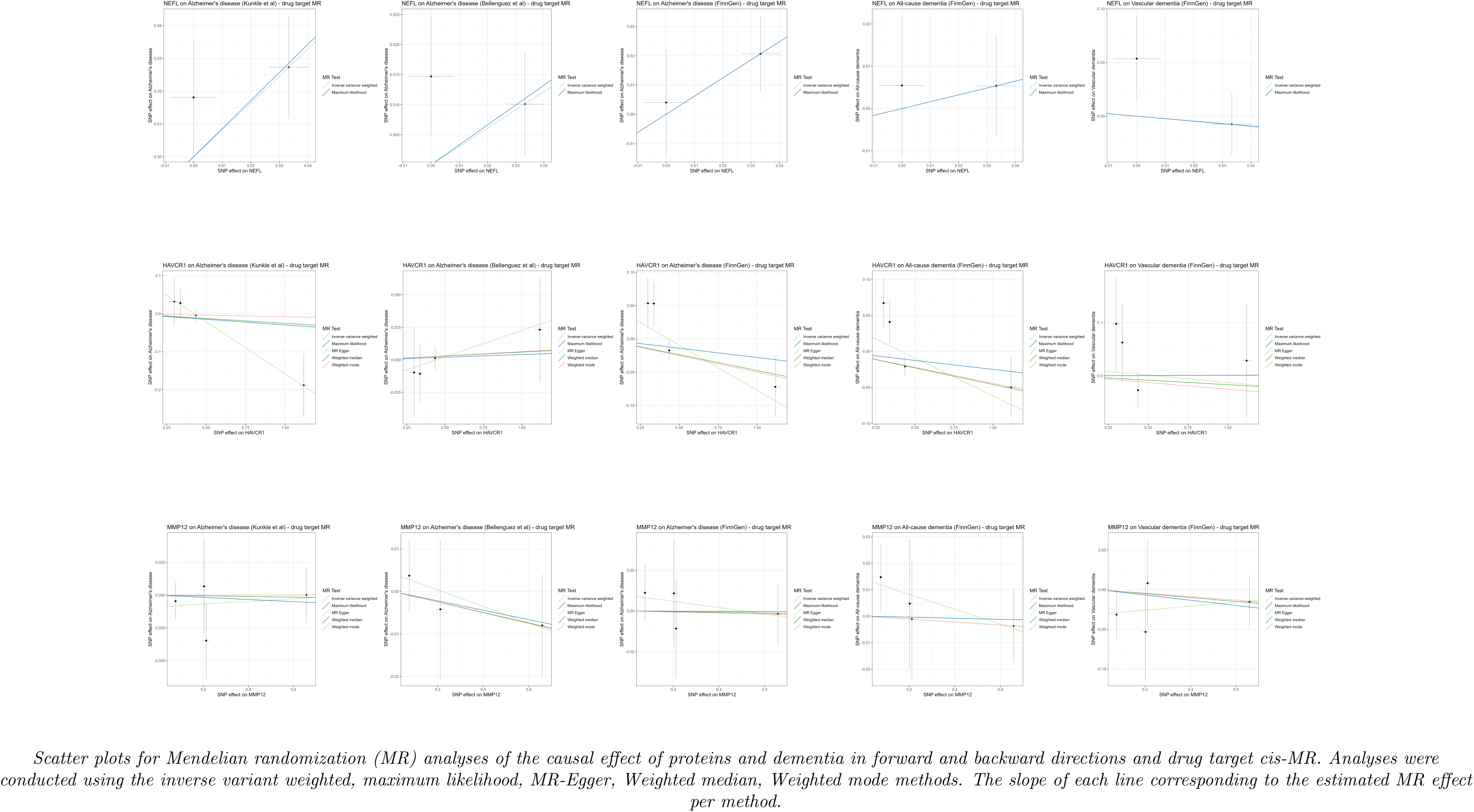
Two-sample bidirectional Mendelian randomisation and drug target Mendelian randomisation scatter plots for four proteins (NEFL, KIM1 (HAVCR1), MMP12, EDA2R) in five GWAS for Alzheimer’s disease, all-cause dementia, and vascular dementia.

MR analyses in the backward direction (dementia → circulating protein concentration) supported **AD** (Bellenguez 2022) ((β [se] = 0.056 [0.014]; P = 1.081 × 10^-4^, based on IVW), **AD** (FinnGen 2023) (β [se] = 0.033 [0.015]; P = 0.024, based on IVW), and **VAD** (β [se] = 0.036 ([0.014]; P = 0.008, based on maximum likelihood) as cause of altered **NEFL** abundance (Fig. 5, Supplementary Table 3). Furthermore, there was evidence supporting a causal link between **AD** (Kunkle 2019) and **MMP12**, demonstrated by MR-Egger, weighted median, and weighted mode methods (β [se] = 0.027 [0.012], P = 0.043; β [se] = 0.025 [0.012], P = 0.037, and β [se] = 0.026 [0.012], P = 0.037, respectively). There was evidence suggesting that **AD** (FinnGen 2023) (β [se] = 0.013 [0.049], P = 0.007 (based on IVW)), **ACD (**β [se] = –0.032 [0.012], P = 0.007 (based on IVW)), and **VAD** (β [se] = –0.036 (0.012), P = 0.003 (based on weighted median)) might have a causal link to altered **EDA2R** abundance.

### Analysis 4: Two sample drug target MR (cis-MR)

The instrument selection for drug target MR (cis-MR) relies on single nucleotide polymorphisms (SNPs) within or near the gene encoding region that regulates the protein of interest. However, the encoding region of EDA2R is located within the *X* chromosome, which precluded the analysis of drug target MR on EDA2R, as the sex chromosomes were excluded from GWAS summary statistics. Cis-MR was conducted for NEFL, KIM1, and MMP12.

In drug target MR, there was no causal evidence for any of the protein-dementia relationships (Fig. 5, Supplementary Table 4).

In sensitivity analysis using a less stringent instrument selection approach, results were largely consistent with the main analysis. There was some evidence indicating a causal relationship between **KIM1** and **AD** (FinnGen 2023) (β [se] = –0.102 [0.041], P = 0.037) and **ACD** (and β [se] = –0.094 [0.037], P = 0.036) based on MR-Egger.

### Analysis 5: Enrichment analysis

In Fig. 6 (also depicted in Supplementary Table 5), the enrichment analyses revealed several biological pathways potentially implicated for the identified proteins (NEFL, RPS6KB1, KIM1, EDA2R, and MMP12), including the immune system, cancers, and insulin signaling. Tissue expression analysis showed expression in the brain for NEFL and in the kidney for KIM1. Notably, one drug, LY2584702, which is a selective, adenosine triphosphate (ATP)-competitive p70S6K inhibitor, has been investigated in clinical trials for the treatment of renal cell carcinoma, metastases, neoplasm, and neuroendocrine tumors – where RPS6KB1 was shown to be implicated in the mechanisms of action of the drug.

**Figure 6.**
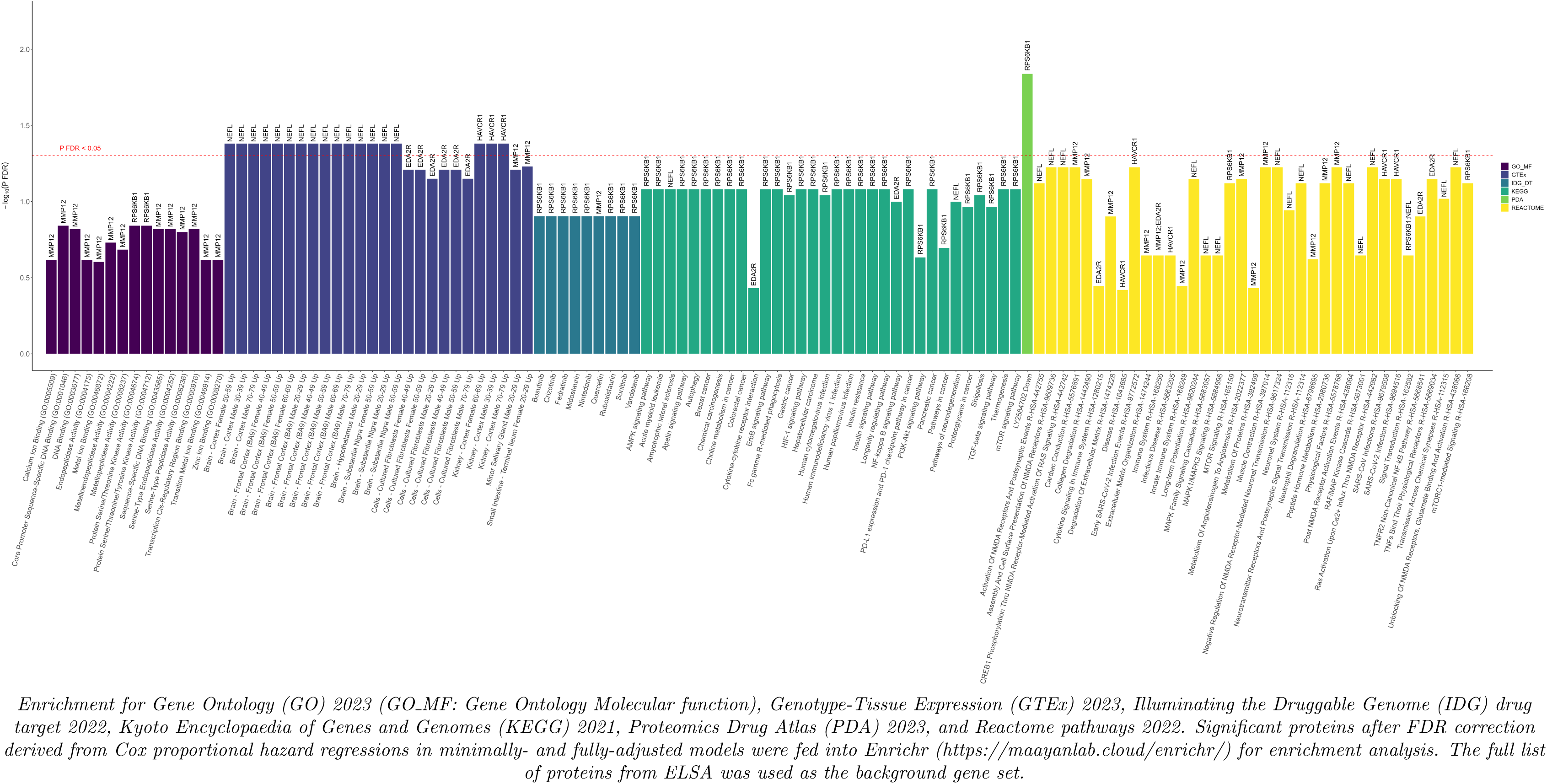
Enrichment analysis of the identified proteins.

Furthermore, upon searching the Open Targets platform for the identified proteins, we identified ten known small molecule drugs in clinical trials (including LY2584702) that are linked to two proteins (RPS6KB1 and MMP12), targeting various cancers, chronic hepatitis C infection, and chronic obstructive pulmonary disease (Supplementary Table 6).

## DISCUSSION

Through a broad proteomics study within the ELSA cohort, encompassing 276 proteins across 3,249 participants, we identified key plasma proteins linked to an elevated risk of incident ACD (NEFL and RPS6KB1) and vascular dementia (MMP12), based on fully adjusted models.

Secondary findings from minimally adjusted models and sensitivity analyses revealed EDA2R and KIM1 as additional significant markers. NEFL and RPS6KB1 individually displayed moderate predictive accuracy for ACD risk (AUC = 0.787 and 0.609, respectively), which yielded an AUC of 0.871 when combined with demographic, genetic, and cognitive factors. Notably, the XGboost machine learning algorithm further underscored RPS6KB1 and NEFL as the most important protein features in predicting ACD onset. These discoveries from ELSA were robustly replicated in the UK Biobank, where NEFL, MMP12, KIM1, and EDA2R were significantly associated with various ACD and dementia subtypes, including AD, VAD, and FTD. Furthermore, employing diverse MR approaches, several causal relationships were observed between AD and VAD with NEFL, AD with MMP12, and between AD, ACD, VAD, with EDA2R in the reverse direction. There was no evidence supporting causal relationships between proteins and dementia from cis-MR analyses.

Elevated NEFL were found to be associated with an increased risk of ACD in ELSA, and with ACD, AD, VAD, and FTD in the UK Biobank. Consistently, a previous study from the UK Biobank ranked NEFL as the most important protein associated with future dementia events out of 1463 protein markers.[16] NEFL is a marker of axonal injury,[19, 20] and is implicated in several biological mechanisms related to dementia,[21, 22] including neurodegeneration,[23, 24] inflammation,[25] central nervous system (CNS) injury,[26–28] and atherosclerosis.[29] It is a well-established (non-specific) marker of neurodegenerative diseases. While NEFL was found in our study to be causally linked to dementia based on the MR findings, the strongest indication was in the backwards direction, which points toward its role as a manifestation of prodromal dementia and anomalies in the brain, rather than a cause of dementia. This underscores the value of NEFL as an important diagnostic and early identification marker, as also demonstrated by the prediction models. It should be noted that the inconsistencies in MR findings from the current study across various AD GWAS could stem from the fact that the chosen genetic variant serving as the instrumental variable might exert a varied impact on the outcome within the represented population. Such differences may arise from variations in genetic backgrounds, environmental influences, or other population-specific factors.

RPS6KB1, functions as a serine/threonine-protein kinase, operating downstream of phosphoinositide 3-kinase (PI3K)/mammalian target of rapamycin (mTOR) signaling in response to growth factors and nutrients, promoting cell proliferation, growth, and progression through the cell cycle.[30] The mTOR complex 1 (mTORC1) signaling was found in a previous study to be implicated in the biological aging process,[31] such that the inhibition of mTOR may extend lifespan given that the mTOR activity becomes abnormally high with age.[31] In the nervous system, the mTOR pathway is implicated in the regulation of synaptic remodeling and long term potentiation.[32–35] Importantly, mTOR plays a crucial role in autophagy regulation in neurons,[35, 36] and the mTOR/p70S6K axis is shown to be essential in the early phases of plasticity for synaptic modifications and the formation of enduring memory.[37] Previous analyses of the ARIC study similarly highlighted the importance of autophagy signaling pathways in the two decades before dementia onset.[10] Interestingly, our sensitivity analysis which excluded APOE ε4 carriers observed an attenuation in association between RPS6KB1 and dementia after considering multiple testing. Previous literature highlighted the mechanisms affected by APOE ε4,[38] such that the presence of APOE ε4 may be necessary for the overactivation in mTOR pathway which subsequently lead to tau hyperphosphorylation and reduced Aβ clearance.[39] At a lower expression level, RPS6KB1 can facilitate the growth of damaged axons resulting from CNS injury.[40] In AD patients, there is a down-regulation of RPS6KB1,[40] and RPS6KB1 has also been implicated in the etiology of other complex neurological diseases including depression and autism.[41] Transcriptomic exploration has revealed the central role of RPS6KB1, and alterations in its co-expression occur during the initial stages of AD, which highlights its potential as a biomarker for the early diagnosis of AD.[42] Several small clinical trials of rapamycin are underway for investigating age-related diseases including AD,[31] with primary outcomes assessing the effects on cognitive performance, and biomarkers of aging.[31] From the Proteomics Drug Atlas, one drug (LY2584702) was found to target RPS6KB1, which is a highly selective adenosine triphosphate competitive inhibitor against p70S6 Kinase. However, the clinical trials associated with LY2584702 mostly target cancers. Further explorations are therefore needed to decipher the relationship between RPS6KB1 and the protein’s pharmacological properties.

Matrix metalloproteinases (MMPs) belong to a multigenic family of membrane-bound or secreted zinc-containing endopeptidases, which indirectly modulate the cellular processes through activation and inactivation of signaling molecules such as trophic factors cytokines, and receptor.[43, 44] MMPs play important roles in cell proliferation and death, neuroinflammation, neurodegeneration, and glial reactivity,[45] and are linked to their proteolytic disruption action on the blood-brain barrier.[46] Based on experimental models, A*β*_40_ contribute to the changes in blood-brain barrier (BBB) permeability, and increased expression of MMPs in transgenic human amyloid precursor protein (hAPP)-overexpressing mice, in turn compromises BBB integrity.[47] There is also an increased expression of MMPs in cerebrospinal fluid of AD patients.[48] Selective inhibitors for MMP12[49] was shown to reduce inflammation and delay of atherosclerosis progression.[50, 51] While observational studies demonstrated an association between high levels of plasma MMP12 and recurrent cardiovascular disease, MR yielded the inverse finding, such that genetic variants associated with higher MMP12 levels are associated with decreased risks of coronary artery disease and large artery atherosclerotic stroke, discouraging potential clinical trials of MMP12 inhibitors.[3, 52] For dementia, the associations between MMP12 and VAD risk in the UK Biobank, and MMP12 and AD risk in the ARIC cohort was similarly highlighted.[10, 16] At an elevated dosage, this medication can penetrate the BBB and manifest an inhibitory effect on metalloproteinase activity within the brain,[53] and was shown to decrease some seizure-related parameters.[53]

In our study, although the significant findings from minimally adjusted models for KIM1 and EDA2R, attenuated after full adjustments, results from the MR analyses showed some possible causal links between these proteins and dementia. We acknowledge however that some of these MR results may be biased by horizontal pleiotropy with the proteins affecting multiple diseases,[54] possibly via immune, renal, and metabolic disease pathways,[54–56] which subsequently contribute to the risk of dementia.[54] There was also evidence from previous studies indicating higher levels of EDA2R were associated a smaller total brain volume, smaller grey matter volume, and less normal-appearing white matter volume.[55]

The current study exhibits robustness through several key strengths. Firstly, it draws upon two extensive population-based cohorts with prolonged follow-up, employing data-driven proteome-wide methods that yield high-throughput and reliable proteomics data. The selection of the protein panel in ELSA is also noteworthy for its focused curation on dementia-related markers, enhancing the study’s precision in investigating associations with dementia risk in a nationally representative sample of older adults. The findings through the inclusion of the UK Biobank cohort encompassing a broader selection of proteins enhance the validity of our results. Both cohorts are well-characterized longitudinal cohorts, which enabled adjustment for wide range of factors. Furthermore, our study benefits from applying a robust and comprehensive dementia algorithm in ELSA, which integrated information from various sources, bolstered by details on medication use and informant-solicited information, which has been reported to correlates better with objective cognitive performance than self-report alone as well as medication which particularly captured those with younger onset dementia,[57] allowing for accurate identification of incident dementia cases and the exclusion of prevalent cases. Another significant strength lies in our approach to assessing protein-dementia associations through a range of established methods for evidence triangulation. Additionally, the utilization of Olink antibody-based PEA is recognized for its superior specificity in proteomics assays.[17] The integration of proteogenomic in MR analyses was an additional strength.[12]

Some limitations should be acknowledged. First, several circulating protein markers potentially relevant to dementia, for example, proteins such as GFAP and GDF-15, were not assayed in ELSA. There is also limited specification, or a lack of protein measurements from the A/T/N classification framework,[58] such as beta-amyloid, p-tau217 and p-tau181. Third, another source of limitation is that the algorithm used for dementia ascertainment lacked information extracted from primary care data, and uncertainties exist ascertaining dementia subtypes. Nevertheless, it is important to note that AD and VAD pathology often co-exist on a population level, and many dementia patients exhibit mixed neuropathology.[59] Fourth, it is important to note that we lacked external validation cohorts for RPS6KB1. Lastly, there are inherent assumptions in MR analyses, and for drug target MR specifically, genetics might not directly inform on specific pharmacological aspects of drug exposure.

In conclusion, in our proteome-wide study from two UK-based large-scale population-based cohorts, we demonstrated the utility of data-driven proteomics analyses in identifying novel targets for dementia. MR analyses leveraging extensive GWAS data substantiated some of these protein-dementia relationships with causal evidence. Looking forward, given the cost-effectiveness and minimal invasiveness of peripheral blood-based biomarkers, developing a panel of blood-based protein markers could greatly enhance dementia diagnostics in routine clinical practice. Integrating large-scale population-based proteomics with other omics, such as genomics,[12] offers immense potential for deeper biological insights into diseases. Additionally, several proteins identified in our study emerge as promising candidates for future clinical trials, addressing the urgent need for more treatment options amid the growing global burden of dementia.

## ONLINE METHODS

### ELSA discovery cohort study population

ELSA is a nationally representative sample of men and women aged 50 years and over living in the community in England. Data collection started in 2002-2003, with participants re-interviewed every two years. Details of survey design are available elsewhere.[60] Blood sample collection in ELSA took place for the first time in wave 2 nurse visit in 2004-2005 and subsequently in every four-year interval. The blood collected from wave 4 nurse visits in 2008-2009 were used for the proteomics profiling (thus forming the baseline sample of this study), thereby affording a unique temporal perspective that allows for the exploration of the relationship between protein concentrations and ADRD over a span exceeding 10 years. The following exclusion criteria were applied: 1) participants who died within 2 years of the wave 4 nurse visit; or 2) participants lost to follow-up (missing at ≥ 2 waves). A total of 3,305 available plasma samples from wave 4 were retrieved for the proteomics profiling.

### Measurement of plasma proteins in ELSA

The proteomics dataset in ELSA was curated with a specific focus on investigating the underlying biological processes associated with ADRD and cognitive decline. The comprehensive assays encompassed an extensive array of cardiovascular and inflammatory markers, in addition to markers integral to neurological processes such as axon guidance, neurogenesis, and synapse assembly. These analyses were conducted utilizing the Olink proteomics platform, the antibody based Olink PEA technology.[61]

We used Olink Target 96 Neurology, Cardiovascular II and Neurology Exploratory arrays in these analyses. The cardiovascular array includes major inflammatory and cardiometabolic pathways relevant to dementia risk, while the neurology array has been designed to include proteins implicated in brain aging. Frozen samples were shipped to Olink for aliquoting, plating, and assays. These assays include a built-in quality control based on four internal controls that are spiked into all samples, and external controls.

Following stringent data quality control (Supplementary Methods), proteins were measured across three panels containing 276 proteins. Proteins were presented as NPX values, the arbitrary unit on log^2^ scale from Olink proteomics.

### Covariates assessment in ELSA

Baseline (at wave 4, 2008-2009) sociodemographic and socioeconomic covariates included age (in years), biological sex (male vs female) and ethnicity (white vs other ethnic groups) were self-reported. The age that participants left formal education was coded as follows: none, age 14 or under, 15, 16, 17, 18, 19 or over. Smoking status was self-reported and was categorized as never, former, and current smoker. Physician-diagnosed cardiovascular disease (heart attack, angina, or heart failure) was self-reported. Depression was also self-reported. BMI was calculated using participant’s height and weight measured during the wave 4 nurse visit. Three measurements were taken of systolic and diastolic pressure on the respondent’s right arm while they were seated, and the average of the three measurements was used. LDL cholesterol was assayed using the blood sample collected by the nurse. The APOE genotype in ELSA was derived from the analysis of two specific SNPs, namely rs7412 and rs429358. To determine these genotypes, two TaqMan assays from Assay-On-Demand, a product of Applied Biosystems and Gene service Ltd in Cambridge, UK, were employed. These assays were conducted on a 7900HT analyzer, manufactured by Applied Biosystems, and the genotypes were determined using the Sequence Detection Software (version 2.0), also from Applied Biosystems. The quality control of genome-wide genotyping has been described elsewhere.[62] Episodic memory was assessed at wave 4, evaluated through the immediate and delayed recall tasks of the Consortium to Establish a Registry for AD (CERAD). Participants were presented with a ten-word list and tasked with recalling it both immediately and after a delay. The scores from these tests were then aggregated to compute a memory score.

### Dementia algorithm in ELSA

The standardized algorithm for identifying dementia cases relied on five primary data sources: 1) coded information extracted from interviews across all waves using participant self-reported physician diagnosis of AD and dementia; 2) caregivers who completed a modified short-form Informant Questionnaire on Cognitive Decline in the Elderly (IQCODE); 3) medication data collected during nurse visits (wave 6, 8, and 9); 4) linked data from hospital admissions (NHS Hospital Episode Statistics) and 5) mortality records (Office for National Statistics Mortality Statistics). All data sources were integrated into the algorithm development process.

Regarding self-reported diagnosis of dementia, during each interview, participants were asked whether a medical professional had informed them of a diagnosis of AD, dementia, organic brain senility, or any other serious memory condition. A positive response indicates the presence of dementia.

Caregivers completed a modified short-form IQCODE on behalf of individuals unable to respond independently. Caregivers (acting as proxies) were instructed to assess the current functional performance of the participant, comparing it to that of two years prior, instead of the standard 10-year interval. Consistent with prior research, individuals with an IQCODE score exceeding 3.38 were classified as having dementia.[63]

Medication data, encoded for analysis, was collected during nurse visits in waves 6 (2012-2013) and wave 8 (2016-2017), and wave 9 (2018-2019). In waves 8 and 9, two mutually exclusive subsets of the sample underwent nurse visits. As specified in the British National Formulary (BNF), we specifically considered four common drugs for dementia: Donepezil hydrochloride (0411000D0), Galantamine (0411000F0), Memantine hydrochloride (0411000G0), and Rivastigmine (0411000E0).

The 10th revision of the International Statistical Classification of Diseases and Related Health Problems (ICD-10) was employed to extract dementia-related outcomes from the National Health Service (NHS) Hospital Episode Statistics (HES) data (see Supplementary Table 7A for ICD-10 codes used for ascertaining all-cause dementia); HES data covered the period between March 1997 and January 2018. Mortality statistics provided by the Office for National Statistics (ONS) were accessible for the period between April 2002 and April 2018. The methodology used to determine ACD using the mortality-linked data mirrored the approach taken with HES data based on ICD codes.

Dementia subtypes, specifically AD and VAD, were also derived (Supplementary Table 7B). In the case of AD, all four data sources were utilized: self-reported physician-diagnosis of AD during each study interview, medication usage based on BNF from nurse visits, and the presence of ICD-10 codes in HES and mortality data. For ascertaining VAD, ICD-10 codes from HES and mortality data were used.

The computation of time-to-event for dementia cases depended on the data source where the event of dementia was first recorded. The date of censoring for non-dementia cases varied across participant groups. Details can be found in the Supplementary Methods.

### UK Biobank for protein validation

UK Biobank is a large population-based cohort from the UK with over half a million participants aged 40 – 69 years, recruited between 2006 and 2010.[18] The UK Biobank Pharma Proteomics Project (UKB-PPP) is a consortium of 13 biopharmaceutical companies which funded the proteomic profiling on blood plasma samples. Proteomics profiling was conducted in 54,219 participants at study baseline, with 2,923 unique proteins assayed using the antibody based Olink Explore 3072 PEA, across eight Olink panels (Cardiometabolic I, Cardiometabolic II, Inflammation I, Inflammation II, Neurology I, Neurology II, and Oncology I, Oncology II). Consortium members opted for samples enriched in specific diseases of interest, while the remaining population was randomly sampled using a stratified approach based on age, sex, and recruitment center.[18] The current analysis excluded those with dementia at study baseline, which yielded a total sample of 52,745 individuals.

We have attempted, where possible, to derive similar variables for both the ELSA and biobank cohorts with consideration of the level of missingness, to maximize comparability. Participant’s age was derived based on date of birth and date of attending an initial assessment center. Participant’s biological sex was acquired from central registry at recruitment and contains a mixture of the sex recorded by the NHS and self-reported sex. Ethnicity was self-reported and categorized into White, Mixed, Asian, or Asian British, and Black or Black British. Highest qualification was determined by the answers provided to the question: “Which of the following qualifications do you have?”, with options included: College or University degree; NVQ (National Vocational Qualification) or HND (Higher National Diploma) or HNC (Higher National Certificate) or equivalent; other professional qualifications e.g.: nursing; A levels/AS levels; O levels/GCSEs (General Certificate of Secondary Education) or equivalent; CSEs (General Certificate of Secondary Education) or equivalent; or none of the above. Smoking status was self-reported and constructed using information about the current/past smoking status of the participant and categorized as never, former, and current smoker. Self-reported medical conditions were solicited through the touchscreen questionnaire as well as during verbal interview conducted by a trained nurse, and the presence of cardiovascular disease (heart attack, angina, or heart failure) were defined if the participant reported any of these conditions. Depression was affirmatory if the participant confirmed to have any of probable recurrent major depression (severe), probable recurrent major depression (moderate), or single probable major depression episode, if reported on the questionnaire or nurse-administered verbal interview. BMI was constructed from height and weight measured during the initial assessment center visit using an Omron device. Two automatic readings of blood pressure were taken a few moments apart, using an Omron HEM-7015IT digital blood pressure monitor, and the average of the readings were used. LDL cholesterol was measured by enzymatic protective selection analysis on a Beckman Coulter AU5800 from the blood sample collected at recruitment.

ACD, AD, VAD and FTD were defined by the UK Biobank dementia algorithm.[64] The last date of censoring was 31^st^ December 2022 (last date of linkage to death and inpatient records). Baseline dementia was removed.

## Statistical analysis

### Analysis 1: Protein-dementia associations in the ELSA discovery cohort

We used Cox proportional hazards regression models to evaluate the associations between each plasma protein NPX value (scaled) and incident dementia using the ‘survival’ R package. Rank-based inverse normal transformation was first applied to the protein levels and scaled to have a mean of 0 and standard deviation of 1 prior to all analyses.[65]

All proteins had ≤ 6% missing. Missing protein measurements were imputed using the K-nearest neighbor (k = 57) imputation using the ‘impute’ R package,[66] which works by identifying the nearest 57 individuals defined using Euclidean distances and imputing with their medians, with k calculated from the square root of the total sample size (N = 3,249). Missingness was imputed for proteomics and clinical data independently, where clinical data was imputed using the Multiple Imputation by Chained Equations procedure (‘mice’ R package),[67] with 30 imputed datasets and 10 iterations.

Based on the normalized and imputed protein data, we first assessed the protein-dementia associations without any model adjustments by pooling the estimates from all 30 imputed datasets. The models were further adjusted for age, sex, and ethnicity for the minimally-adjusted model; and adjusted for age, sex, ethnicity, education, smoking status, depression, cardiovascular disease, BMI, systolic blood pressure, and LDL cholesterol for the fully-adjusted models for each protein NPX value associated with the risk of incident dementia, with P_FDR_ set at a cut-off of 0.05, this translates to an uncorrected p-value of 0.00018 (with 276 tests for significance for 276 proteins). All p-values were two-sided. FDR-corrected p-values (denoted as P_FDR_) were reported and displayed using volcano plot, accompanying the HR. The Cox proportional hazard regression analyses were repeated for AD and VAD as the outcome of interest separately.

A series of sensitivity analyses were conducted using the same adjustment methods as the main Cox regression for incident ACD, by excluding the following: 1) dementia cases that occurred during the first year of follow-up to reduce the possibility of reverse causation bias; 2) other ethnic groups other than white ethnicity; 3) APOE ε4 carriers; and 4) participants <60 years of age. We also conducted competing risk of death using Fine-Gray regression models[68] with the same set of covariate adjustments, given that death may preclude dementia from occurring. The competing risk models, which estimated sub-distribution HRs, were conducted for incident ACD accounting for all-cause mortality as a competing risk, incorporating time to event data.[68, 69] Lastly, sex differences in the protein-dementia associations were assessed by fitting sex as an interaction term in the Cox regression models.[70]

Next, receiver operating characteristic (ROC) analyses were conducted to assess the precision of the identified proteins from the fully adjusted Cox regression models in predicting incident ACD. These analyses were performed independently and in combination with additional factors including age, sex, ethnicity, education, APOE ε4 status, and memory score. To evaluate the performance of the Cox models, bootstrapping was performed to assess the stability of the AUC estimates. A total of 2000 bootstrap resamples were generated. Bootstrapping involves resampling with replacement from the original dataset to estimate the sampling distribution of a statistic. We used the bootstrapped samples to compute 95% CI for the AUC of each model, utilizing the R packages ‘caret’[71], ‘boot’[72] and ‘pROC’.[73] The mean AUC value across all bootstrap resamples was calculated to estimate the overall predictive performance of the survival model.

We utilized eXtreme Gradient Boosting (XGBoost), an advanced decision-tree ensemble machine learning algorithm within a gradient boosting framework, to identify the most important features predictive of future ACD onset.[74] To ensure comprehensive analysis and validation, we incorporated all available protein, demographic, and clinical predictors into our models, using imputed data to handle any missing values. The XGBoost model feature importance scores were used to select key predictors. To explain the outputs of our XGBoost models and to provide interpretability to the feature importance rankings, we employed SHapley Additive exPlanations (SHAP) values. SHAP values offer a consistent and theoretically sound method for interpreting the contributions of individual features to the model’s predictions.[75] Furthermore, this process of evidence triangulation with XGBoost and SHAP values was conducted to validate and complement the findings from our Cox regression models. By comparing and cross validating, the results from these two different methodological approaches, we aimed to enhance the robustness and reliability of our predictive model for future ACD onset.

### Analysis 2: Protein-dementia associations in the UK Biobank validation cohort

The significant proteins from the main and sensitivity analyses based on ELSA were then validated using the UK Biobank proteomics data, if assayed. The reason for choosing ELSA as the discovery cohort and UK Biobank as the validation cohort is that ELSA’s target panels were specifically designed to investigate proteomic signatures related to cognitive decline and dementia, resulting in a more focused selection of proteins. In contrast, UK Biobank offers a broader protein selection, allowing for effective validation.

The UKB-PPP proteomics samples underwent sample selection, processing, and quality control procedures.[18] Missing protein measurements for the remaining individuals were imputed using K-nearest neighbor imputation (k = 230), the protein levels were normalized and scaled akin to the analyses in ELSA proteomics data described above.[65] Clinical data was imputed with 30 imputed datasets and 10 iterations, and was imputed separately to the proteomics data.

Data was processed and analyzed using the R Studio Workbench on the UK Biobank Research Analysis Platform, under application No. 71702.

### Analysis 3: Two-sample bi-directional MR

MR pertains to employing genetic variants to explore causal connections regarding the influence of modifiable exposures on a range of outcomes. Anchored in Mendel’s laws of inheritance and instrumental variable estimation techniques, MR principles facilitate the inference of causal effects, even in the presence of unobserved confounding factors. MR studies are often compared to naturally occurring randomized trials, resembling a randomized control trial where genetic factors are inherently assigned at random by nature.[76] The MR approach rests on three key assumptions: 1) the instruments are correlated with the exposure; 2) the instruments are linked to the outcome solely through the studied exposure (exclusion restriction assumption); and 3) the instruments are independent of other factors influencing the outcome (independence assumption).

To infer causality between dementia-associated proteins and AD, ACD, and VAD, we performed bidirectional two-sample MR, leveraging summary statistics from different GWAS consortia. For AD, we used GWAS summary statistics derived from three consortia: (International Genomics of Alzheimer’s Project (IGAP) (Kunkle et al 2019),[77] European Alzheimer & Dementia Biobank (EADB) consortium (Bellenguez et al 2022),[78] and the FinnGen 2023 study);[79] For ACD and vascular dementia GWAS were both derived from the FinnGen 2023 study.[79] Summary details of the dementia GWAS used were included in Supplementary Table 8.

Selection of instruments to proxy for altered protein abundance were derived using pQTL mapping proteins that identifies genetic associations in participants of European ancestry from the UK Biobank based on Olink data (https://doi.org/10.7303/syn51364943).[18] The effects of protein pQTL were standardized to align with the same effect allele.

To ensure the three MR assumptions were not violated, instruments for the exposure were selected by using association at genome-wide significance (P < 5 × 10^−8^). SNP with a minor allele frequency (MAF) < 5% were excluded. linkage disequilibrium (LD) clumping was done with a window size of 10,000 kilobases [kb] at an R^2^ threshold of 0.001. In cases where a requested SNP from the exposure GWAS is not found in the outcome GWAS, a proxy SNP with a LD coefficient of R^2^ > 0.8 to the requested missing target SNP were sought as a substitute, if available, by querying the LDLink web server. LD proxies are determined using data from the 1000 Genomes European sample. The returned information includes the effect of the proxy SNP on the outcome, along with details such as the proxy SNP itself, the effect allele of the proxy SNP, and the corresponding allele for the target SNP. The effect of a SNP on an outcome and exposure were then harmonized to be relative to the same allele. We specify the inference of positive strand alleles by utilizing allele frequencies for palindromes instead of eliminating palindromic variants. F-statistics were used to assess the SNP-exposure strength (F > 10).

We first performed the MR in the forward direction (circulating protein concentration → dementia) on the harmonized effects to estimate the effect of genetically proxied protein abundance on genetic liability to the dementia outcome of interest to be relative to the same allele. Given the study outcomes were AD and related dementia, variants in the *APOE* region (human genome reference builds GRCh38 – chromosome: 19, base pair position: 44,407,913 – 45,408,821) were removed due to its pleiotropic nature and large effect size. We then calculated the effects for each individual variant by employing a two-term Taylor series expansion of the Wald ratio. Subsequently, we utilized the weighted delta inverse-variance weighted (IVW) method to conduct a meta-analysis of individual SNP effects, to estimate the combined effect of the Wald ratios. Sensitivity analyses included using various MR methods, including MR-Egger, Weighted Median, Maximum Likelihood, Weighted Mode, and leave-one-out methods.[76]

The same method was then applied to the backward MR (dementia → circulating protein concentration), with SNPs extracted from the same GWAS as described above, except with dementia being the exposure data and protein being the outcome.

All analyses were conducted using human genome reference build GRCh38. In instances where the genome build was based on GRCh37 assembly Hg19, a lift-over process was executed to convert genome coordinates and annotations to the GRCh38 using a designated alignment. All MR analyses were conducted using ‘TwoSampleMR’, ‘MendelianRandomization’ and ‘LDlinkR’ R packages.[80]

### Analysis 4: Two sample drug target MR (cis-MR)

Next, we used a two-sample MR study design, based predominantly on genetic variants located in or near genes that encode the relevant drug targets, to infer causality from protein concentration → dementia (cis-MR).[81–84] Cis-MR is considered to be less susceptible to pleiotropy, and the potential effect of a drug by analyzing the genomic locus encoding protein targets, which may be informative for drug trial design.[82, 84]

The selection of the primary cis-MR models included SNPs successfully harmonized with the gene encoding regions, with the flanking region being within 10-kilobase (kB) in either direction of the start and stop coordinates for the genes, according to Human Genome reference release GRCh38. LD clumping was then done to remove the excess of most highly correlated variants at each locus, with R^2^ threshold set at < 0.001, retaining the SNPs with the strongest associations with the protein of interest filtered by P < 5 × 10^−5^. The sensitivity analyses which took a more liberal approach, selecting valid instruments at the threshold of P < 5 × 10^−5^ and clumped at R^2^ < 0.01.

All analyses were done on human genome reference builds – GRCh38, lift over was performed from GRCh 37 assembly Hg19 to GRCh 38, using CrossMap 0.7.0 in Python (version 3.12). Analysis 1 to 4 were done using R Studio (version 4.4.0).

### Analysis 5: Enrichment analysis

Enrichment analysis was conducted by searching open-source databases to further characterize the identified proteins from the Cox proportional hazard regressions. We employed the Enrichr,[85] which is a computational method infers knowledge about an input gene set by comparing it to annotated gene sets that represent existing biological knowledge. It determines if the input set of genes significantly overlaps with these annotated gene sets. We used the full set of ELSA proteins as the background gene set, to glean a deeper biological understanding.

We searched the following bioinformatics databases: Gene Ontology (GO): GO Molecular Function, GO Biological Process, and GO Cellular Component [86], Kyoto Encyclopedia of Genes and Genomes (KEGG) [87], Reactome Pathway Database (REACTOME) [88], Illuminating the Druggable Genome (IDG) [89], Proteomics Drug Atlas (PDA) [90], and Genotype-Tissue Expression (GTEx) [91]. Statistical significance was indicated if P_FDR_ < 0.05, considering multiple testing.

Furthermore, we utilized the Open Targets platform (https://www.opentargets.org/) for the systematic identification of potential therapeutic drug targets among the identified proteins. [92]

## Supporting information

This file contains all supplementary methods, supplementary tables, and supplementary figures.

## Acknowledgement

The National Institute of Aging (NIA) (grant No. [R01AG17644]) funded the proteomics data curation in ELSA. J.G. is supported by the NIA (grant No. [R01AG17644]). D.M.W. is supported by the British Medical Research Council (MRC) (grant No. [MC_UU_00019/3]).

This research has been conducted using the UK Biobank Resource under application number 71702. This work uses data provided by patients and collected by the NHS as part of their care and support. Copyright © 2023, NHS England. Re-used with the permission of the NHS England and UK Biobank. All rights reserved.

We would like to thank the participants in ELSA for their contribution to the research. We additionally want to acknowledge the participants and investigators of the FinnGen study. We would like to thank Olink representatives and the Newcastle laboratory for their support for data procurement and data pre-processing.

## Conflict of interests

Olink had no part in designing the study or analyzing the data. No conflicts of interest to be declared from any of the authors.

## Author contributions

The study’s conceptualization and design were undertaken by A.S., P.Z., and J.G. Proteomics data curation in ELSA was a joint effort between J.G. and A.S. J.G. handled all data processing and analysis and wrote the initial manuscript draft. A.S. and D.M.W. provided oversight and guidance for statistical analyses and theoretical frameworks. D.M.W. provided access to the UK Biobank data, under project application No. 71702. J.G. led the analysis and interpretation of data, as well as manuscript revisions. J.G. led the development of the dementia algorithm in ELSA, with contributions from A.S., S.S., S.A., F.B., and S.H. All authors participated in the manuscript’s revision process and collectively accepted responsibility for its submission for publication.

## Data availability

The ELSA data is available on UK Data Service. The proteomics data in ELSA will be deposited on UK Data Service upon publication. All data from the UK Biobank, including the proteomics data is available by directly submitting a project application to the UK Biobank. All GWAS summary statistics are available online: https://doi.org/10.7303/syn51364943; https://www.finngen.fi/en/access_results; https://gwas.mrcieu.ac.uk/.

## Code availability

The codes used for all analyses are available on GitHub repository: https://github.com/jgong94/ELSA_proteomics_dementia.

## Notes

### Competing Interest Statement

The authors have declared no competing interest.

### Author Declarations

Ethical consent has been obtained for all waves and components of ELSA, by the National Research Ethics Service. UK Biobank has approval from the North West Multi-centre Research Ethics Committee (MREC) as a Research Tissue Bank (RTB) approval.

