## Supplementary material for "Unraveling the role of plasma proteins in dementia: insights from two cohort studies in the UK, with causal evidence from Mendelian randomization": This file contains all supplementary methods, supplementary tables, and supplementary figures.

**Supplementary Materials**

**Content**

**Supplementary Methods**


| | |
|---|---|
| ELSA proteomics data quality control pipeline. | Page 3-4 |
| Time to event censoring for dementia in ELSA. | Page 5-6 |


**Supplementary Tables**


| | |
|---|---|
| Supplementary Table 1. Demographic and phenotypic information of the ELSA and UK Biobank participants. | Page 7-8 |
| Supplementary Table 2. Multiple-adjusted hazard ratios and 95% confidence intervals for the associations between identified proteins and dementia outcomes in the UK Biobank. | Page 9 |
| Supplementary Table 3. Two sample bi-directional Mendelian randomization between proteins and dementia. | Page 10-13 |
| Supplementary Table 4. Two sample cis-Mendelian randomization between proteins and dementia. | Page 14-16 |
| Supplementary Table 5. Enrichment analysis for the identified proteins. | Page 17-20 |
| Supplementary Table 6. Drugs linked to the identified proteins associated with dementia, drug type, their potential mechanisms of action, current target disease, and the status and phase of the trials. | Page 21 |
| Supplementary Table 7A. International classification of disease codes for definite dementia as recorded in hospital admission and mortality records in ELSA. | Page 22 |
| Supplementary Table 7B: International classification of disease codes by dementia sub-type for definite dementia cases in ELSA. | Page 23 |
| Supplementary Table 8. Summary information of dementia GWAS datasets. | Page 24 |


**Supplementary Figures**


| | |
|---|---|
| Supplementary Figure 1. Flow diagram for ELSA proteomics project sample selection and quality control pipeline. | Page 25 |
| Supplementary Figure 2. Dementia cases by data source in ELSA. | Page 26 |
| Supplementary Figure 3. Boxplot for all proteins in ELSA by dementia status. | Page 27-29 |
| Supplementary Figure 4. Volcano plot showing the unadjusted HR (x axis) and two-sided P values (y axis) for the association between protein concentration with incident all-cause dementia using imputed data. | Page 30 |
| Supplementary Figure 5. Volcano plot showing the sex-, age- and ethnicity-adjusted HR (x axis) and two-sided P values (y axis) for the association between protein concentration with incident all-cause dementia using imputed data. | Page 31 |
| Supplementary Figure 6. Volcano plot showing the fully adjusted HR (x axis) and two-sided P values (y axis) for the association between protein concentration with incident all-cause dementia, excluding other ethnic groups using imputed data. | Page 32 |
| Supplementary Figure 7. Volcano plot showing the fully adjusted HR (x axis) and two-sided P values (y axis) for the association between protein concentration with incident all-cause dementia, excluding APOE 4 carriers using imputed data. | Page 33 |
| Supplementary Figure 8. Volcano plot showing the fully adjusted HR (x axis) and two-sided P values (y axis) for the association between protein concentration with incident all-cause dementia, after reducing the possibility of reverse causation bias by excluding all-cause dementia cases that occurred during the first year of follow-up using imputed data. | Page 34 |

| | |
|---|---|
| Supplementary Figure 9. Volcano plot showing the fully adjusted HR (x axis) and two-sided P values (y axis) for the association between protein concentration with incident all-cause dementia, excluding participants <60 years using imputed data. | Page 35 |
| Supplementary Figure 10. Volcano plot showing the fully adjusted sub-distribution HR (x axis) and two-sided P values (y axis) for the association between protein concentration with incident all-cause dementia, using Fine-Gray competing risk regression using imputed data. | Page 36 |
| Supplementary Figure 11. Volcano plot showing the fully adjusted HR (x axis) and two-sided P values (y axis) for the association between protein concentration with incident Alzheimer's disease using imputed data. | Page 37 |
| Supplementary Figure 12. Volcano plot showing the fully adjusted HR (x axis) and two-sided P values (y axis) for the association between protein concentration with incident vascular dementia using imputed data. | Page 38 |

### Supplementary Methods

#### ELSA proteomics data quality control pipeline

This section provides a summary of the study design and quality control (QC) protocol implemented by the proteomics data curation within the English Longitudinal Study of Ageing (ELSA) for normalized protein expression (.NPX) data generated using the antibody based Olink™ Proximity Extension Assay (PEA) in blood plasma samples derived from 3262 ELSA participants in Wave 4 in 2008.

Data is presented as normalized protein expression (NPX) values, Olink Proteomics' arbitrary unit on $log_2$ scale. Data points for samples that did not pass QC are written in red text in the raw data files derived from Olink.

The protocol follows the following curation and quality control pipeline:

1. Data pre-processing and internal quality control by Olink

Four internal controls are added to each sample to monitor the quality of assay performance, as well as the quality of individual samples. The QC is performed in two steps:

- Each sample plate is evaluated on the standard deviation (SD) of the internal controls. This should be below 0.2 NPX. Only data from sample plate that pass this quality control will be reported.
- The quality of each sample is assessed by evaluating the deviation from the median value of the controls for each individual sample. Samples that deviate less than 0.3 NPX from the median pass the quality control.

2. Importing data and removing Olink control samples

3. Removing data with quality control warnings or assay warnings

Samples that did not pass the QC are indicated in columns named "QC Warning". Data points from samples that do not pass QC should be treated with caution.

4. Outlier sample detection and removal

IQR-median and principal component analyses (PCA) are used to detect and visualize outliers. Generates PCA projection of all samples from NPX data along two principal components (Default PC2 vs PC1) colored by the variable QC_Warning and including the percentage of explained variance. By default, the values scaled and cantered in the PCA and proteins with missing NPX values removed from the corresponding assay(s). Imputation by median value is done for assays with missingness <10% and for multi-plate projects.

Based on these analyses, we then removed: 1) samples with a PC1 or PC2 value more than 5 SD from the mean, and 2) samples with median concentrations (NPX values) across proteins that were more than 5 SD from the mean median, or those with IQR (NPX) across proteins that were great than 5 SD from the mean IQR. The removal of outliers is panel specific.

**Time to event and time to censoring for dementia in ELSA**

The term "time to event" pertains to the duration until a specific predefined endpoint of interest occurs. The methodology for determining the time to event in participants with a documented event of dementia varied by the source where the dementia event was initially established. This divergence in assessment methods is summarized as follows:

1. *Interview:* If the earliest date of diagnosed dementia was reported at the time of interview, we calculated the time to event by determining the midpoint between the interview date when doctor-diagnosed dementia was first reported and the date of the preceding interview. Participants who reported diagnosed dementia at the first interview were classified as prevalent cases.
2. *IQCODE:* If the earliest date corresponded to dementia reported through the proxy/informant method using IQCODE data (and considering that the questionnaire solicited information pertaining to the preceding two years), the time to event was computed as one year preceding the interview date where the IQCODE score was calculated as $< 3.38$.
3. *Hospital Episode Statistics (HES):* When the earliest date of dementia was derived from HES, we used the exact date of the dementia-related admission (HES datasets included appointments at Outpatient and Admitted Patient Care services and attendance at Accident and Emergency units).
4. *Mortality statistics:* In cases where the earliest date of dementia was established through linkage to mortality data, we used the date of death.
5. *Medications*: When the earliest date of dementia was deduced from medication information obtained during the nurse visits, we calculated the time to event by determining the midpoint between the date of the nurse visit when dementia medication usage was recorded and the date of the interview prior to the nurse visit.

For the participants not recorded as having dementia, the date of censoring varied depending on conditions such as the consent given for linkage to hospital episodes and mortality data:

1. If the participant's death was confirmed by mortality data, they were censored at the date of death.

2. If the participant provided consent for mortality statistics linkage (and did not later revoke it) and was not recorded as having died, they were censored on 15th April 2018, the last date of linkage).
3. If the participant consented to HES linkage (and did not later revoke it) and was not recorded as having died, they were censored on 15th January 2018, the last date of linkage.
4. For participants who did not consent to HES or mortality linkage, the date of censoring was set at the date of their last interview.

### Supplementary Tables

**Supplementary Table 1. Demographic and phenotypic information of the ELSA and UK Biobank participants.**

| | Overall | ACD | AD | VAD | FTD |
|---|---|---|---|---|---|
| **ELSA, study wave 4 (2008 – 2009)** | | | | | |
| **Sample N** | 3249 | 229 | 89 | 41 | - |
| **Age (years)**, Mean (SD) | 63.4 (9.20) | 75.1 (9.40) | 73.9 (9.25) | 76.5 (9.59) | - |
| **Sex**, N (%) | | | | | - |
| Female | 1786 (54.97) | 137 (59.83) | 57 (64.04) | 20 (48.78) | - |
| Male | 1463 (45.03) | 92 (40.17) | 32 (35.96) | 21 (51.22) | - |
| **Ethnicity**, N (%) | | | | | - |
| White | 3157 (97.17) | 226 (98.69) | 89 (100.00) | 41 (100.00) | - |
| Other ethnic groups | 92 (2.83) | 3 (1.31) | 0 (0.00) | 0 (0.00) | - |
| **Age completed full-time education**, N (%) | | | | | |
| None | 16 (0.49) | 3 (1.31) | 2 (2.25) | 0 (0.00) | - |
| Age 14 or under | 271 (8.34) | 62 (27.07) | 21 (23.60) | 14 (34.15) | - |
| Age 15 | 1023 (31.49) | 72 (31.44) | 28 (31.46) | 11 (26.83) | - |
| Age 16 | 745 (22.93) | 33 (14.41) | 9 (10.11) | 7 (17.07) | - |
| Age 17 | 265 (8.16) | 15 (6.55) | 7 (7.87) | 1 (2.44) | - |
| Age 18 | 248 (7.63) | 12 (5.24) | 5 (5.62) | 1 (2.44) | - |
| Age 19 or over | 631 (19.42) | 27 (11.79) | 16 (17.98) | 7 (17.07) | - |
| **Smoking status**, N (%) | | | | | |
| Never smoker | 1319 (40.60) | 96 (41.92) | 35 (39.33) | 14 (34.15) | - |
| Former smoker | 1475 (45.40) | 109 (47.60) | 45 (50.56) | 23 (56.10) | - |
| Current smoker | 448 (13.79) | 24 (10.48) | 9 (10.11) | 4 (9.76) | - |
| **Cardiovascular disease**, N (%) | 259 (7.97) | 42 (18.34) | 11 (12.36) | 9 (21.95) | - |
| **Depression**, N (%) | 198 (6.09) | 14 (6.11) | 6 (6.74) | 3 (7.32) | |
| **Body mass index**, Mean (SD) | 28.11 (5.07) | 27.66 (5.05) | 26.65 (4.83) | 28.64 (5.14) | - |
| **Systolic blood pressure**, Mean (SD) | 131.7 (17.3) | 136.4 (21.6) | 133.5 (16.6) | 146.2 (22.3) | - |
| **LDL cholesterol**, Mean (SD) | 3.28 (1.02) | 2.96 (1.07) | 3.18 (1.08) | 2.70 (1.12) | - |
| **UK Biobank, study baseline (2006 – 2010)** | | | | | |
| **Sample N** | 52745 | 1506 | 732 | 281 | 111 |
| **Age (years)**, Mean (SD) | 56.8 (8.21) | 64.5 (5.10) | 65.0 (4.97) | 65.0 (4.53) | 62.1 (6.04) |
| **Sex**, N (%) | | | | | |
| Female | 28436 (53.9) | 732 (48.61) | 402 (54.92) | 110 (39.15) | 48 (43.24) |

| | | | | | |
|---|---|---|---|---|---|
| Male | 24309 (46.1) | 774 (51.39) | 330 (45.08) | 171 (60.85) | 63 (56.76) |
| **Ethnicity**, N (%) | | | | | |
| White | 49194 (93.3) | 1448 (96.15) | 703 (96.04) | 274 (97.51) | 109 (98.20) |
| Black | 1210 (2.29) | 26 (1.73) | 15 (2.05) | 2 (0.71) | 1 (0.90) |
| Asian | 1132 (2.15) | 9 (0.60) | 3 (0.41) | 2 (0.71) | 0 (0.00) |
| Other or mixed | 958 (1.82) | 15 (1.00) | 7 (0.96) | 2 (0.71) | 1 (0.90) |
| **Educational attainment**, N (%) | | | | | |
| College or University degree | 16887 (32.02) | 336 (22.31) | 154 (21.04) | 46 (16.37) | 31 (27.93) |
| A levels/AS levels | 5796 (10.99) | 125 (8.30) | 60 (8.20) | 32 (11.39) | 8 (7.21) |
| O levels/GCSEs or equivalent | 10894 (20.65) | 291 (19.32) | 148 (20.22) | 58 (20.64) | 14 (12.61) |
| CSEs or equivalent | 2813 (5.33) | 25 (1.66) | 9 (1.23) | 1 (0.36) | 4 (3.60) |
| NVQ or HND or HNC or equivalent | 3485 (6.61) | 96 (6.37) | 42 (5.74) | 27 (9.61) | 7 (6.31) |
| Other professional qualifications e.g.: nursing, teaching | 2780 (5.27) | 88 (5.84) | 42 (5.74) | 16 (5.69) | 10 (9.01) |
| None of the above | 9295 (17.62) | 493 (32.74) | 249 (34.02) | 93 (33.10) | 34 (30.63) |
| **Smoking status**, N (%) | | | | | |
| Never smoker | 28533 (54.10) | 722 (47.94) | 363 (49.59) | 116 (41.28) | 54 (48.65) |
| Former smoker | 18390 (34.87) | 636 (42.23) | 302 (41.26) | 133 (47.33) | 43 (38.74) |
| Current smoker | 5567 (10.55) | 139 (9.23) | 61 (8.33) | 30 (10.68) | 13 (11.71) |
| **Cardiovascular disease**, N (%) | 2796 (5.30) | 201 (13.35) | 76 (10.38) | 53 (18.86) | 13 (11.71) |
| **Depression,** N (%) | 5835 (11.06) | 177 (11.75) | 70 (9.56) | 39 (13.88) | 23 (20.72) |
| **Body mass index**, Mean (SD) | 27.47 (4.81) | 27.77 (4.92) | 27.40 (4.84) | 28.55 (4.99) | 27.58 (4.95) |
| **Systolic blood pressure**, Mean (SD) | 139.7 (19.7) | 146.0 (20.5) | 146.8 (20.6) | 146.1 (20.3) | 142.2 (19.5) |
| **LDL cholesterol**, Mean (SD) | 3.53 (0.88) | 3.38 (0.93) | 3.42 (0.94) | 3.31 (1.01) | 3.58 (0.94) |

*ELSA, English Longitudinal Study of Ageing; SD, standard deviation; ACD, all-cause dementia; AD, Alzheimer's disease; VAD, vascular dementia; FTD, frontotemporal dementia; LDL, low-density lipoprotein; GCSE, General Certificate of Secondary Education; CSE, Certificate of Secondary Education; NVQ, National Vocational Qualification; HND, Higher National Diploma; HNC, Higher National Certificate.*

**Supplementary Table 2. Multiple-adjusted hazard ratios and 95% confidence intervals for the associations between identified proteins and dementia outcomes in the UK Biobank.**

| | UK Biobank | | |
|---|---|---|---|
| | **HR (95% CI)** | **$P_{FDR}$** | **$P_{Uncorrected}$** |
| All-cause dementia | | | |
| NEFL | 1.87 (1.75, 1.99) | $1.02 \times 10^{-81}$ | $2.54 \times 10^{-82}$ |
| KIM1 | 1.13 (1.06, 1.20) | $3.15 \times 10^{-4}$ | $7.87 \times 10^{-5}$ |
| MMP12 | 1.17 (1.10, 1.24) | $2.00 \times 10^{-6}$ | $4.99 \times 10^{-7}$ |
| EDA2R | 1.31 (1.22, 1.40) | $3.18 \times 10^{-13}$ | $7.94 \times 10^{-14}$ |
| Alzheimer's disease | | | |
| NEFL | 1.81 (1.65, 1.99) | $1.89 \times 10^{-35}$ | $4.72 \times 10^{-36}$ |
| KIM1 | 1.11 (1.02, 1.21) | 0.077 | 0.019 |
| MMP12 | 1.10 (1.01, 1.20) | 0.129 | 0.032 |
| EDA2R | 1.25 (1.13, 1.39) | $6.06 \times 10^{-5}$ | $1.52 \times 10^{-5}$ |
| Vascular dementia | | | |
| NEFL | 1.90 (1.64, 2.19) | $1.59 \times 10^{-17}$ | $3.99 \times 10^{-18}$ |
| KIM1 | 1.44 (1.25, 1.66) | $1.13 \times 10^{-6}$ | $2.82 \times 10^{-7}$ |
| MMP12 | 1.36 (1.18, 1.56) | $6.85 \times 10^{-5}$ | $1.71 \times 10^{-5}$ |
| EDA2R | 1.34 (1.15, 1.58) | $1.20 \times 10^{-3}$ | $3.01 \times 10^{-4}$ |
| Frontotemporal dementia | | | |
| NEFL | 2.97 (2.39, 3.70) | $1.10 \times 10^{-21}$ | $2.75 \times 10^{-22}$ |
| KIM1 | 0.90 (0.72, 1.12) | 0.999 | 0.347 |
| MMP12 | 0.87 (0.70, 1.08) | 0.860 | 0.215 |
| EDA2R | 0.99 (0.77, 1.28) | 0.999 | 0.938 |

*All models adjusted for age, sex, education, ethnicity, smoking status, depression, cardiovascular disease, body mass index, systolic blood pressure, LDL cholesterol. P values were false discovery rate (FDR) corrected.*

**Supplementary Table 3. Two sample bi-directional Mendelian randomization between proteins and dementia.**

| Gene name | Outcome trait | MR methods | Forward (protein → dementia) | | | Backward (dementia → protein) | | |
|---|---|---|---|---|---|---|---|---|
| | | | No of SNP | Slope (SE) | P value | No of SNP | Slope (SE) | P value |
| NEFL | Alzheimer's disease, *Kunkle et al 2019* | Inverse variant weighting | 7 | -0.208 (0.157) | 0.185 | 19 | 0.019 (0.013) | 0.134 |
| NEFL | Alzheimer's disease, *Kunkle et al 2019* | MR-Egger | 7 | 0.307 (0.561) | 0.608 | 19 | 0.009 (0.018) | 0.628 |
| NEFL | Alzheimer's disease, *Kunkle et al 2019* | Weighted Median | 7 | -0.151 (0.213) | 0.479 | 19 | -0.005 (0.013) | 0.724 |
| NEFL | Alzheimer's disease, *Kunkle et al 2019* | Maximum-likelihood | 7 | -0.211 (0.159) | 0.709 | 19 | 0.019 (0.009) | 0.032 |
| NEFL | Alzheimer's disease, *Kunkle et al 2019* | Weighted Mode | 7 | 1.028 (2.627) | 0.182 | 19 | 0.001 (0.015) | 0.930 |
| NEFL | Alzheimer's disease, *Kunkle et al 2019* | Leave-one-out | 7 | -0.208 (0.157) | 0.185 | 19 | 0.019 (0.013) | 0.134 |
| NEFL | Alzheimer's disease, *Bellenguez et al 2022* | Inverse variant weighting | 9 | 0.017 (0.125) | 0.890 | 59 | 0.056 (0.014) | $1.081\times10^{-4}$ |
| NEFL | Alzheimer's disease, *Bellenguez et al 2022* | MR-Egger | 9 | 0.592 (0.417) | 0.200 | 59 | 0.030 (0.025) | 0.238 |
| NEFL | Alzheimer's disease, *Bellenguez et al 2022* | Weighted Median | 9 | 0.080 (0.128) | 0.532 | 59 | 0.029 (0.018) | 0.100 |
| NEFL | Alzheimer's disease, *Bellenguez et al 2022* | Maximum-likelihood | 9 | 0.020 (0.077) | 0.803 | 59 | 0.057 (0.011) | $4.831\times10^{-7}$ |
| NEFL | Alzheimer's disease, *Bellenguez et al 2022* | Weighted Mode | 9 | -1.156 (1.990) | 0.577 | 59 | 0.024 (0.020) | 0.233 |
| NEFL | Alzheimer's disease, *Bellenguez et al 2022* | Leave-one-out | 9 | 0.017 (0.125) | 0.890 | 59 | 0.056 (0.014) | $1.081\times10^{-4}$ |
| NEFL | Alzheimer's disease, *FinnGen 2023* | Inverse variant weighting | 9 | 0.086 (0.158) | 0.584 | 22 | 0.033 (0.015) | 0.024 |
| NEFL | Alzheimer's disease, *FinnGen 2023* | MR-Egger | 9 | -0.464 (0.569) | 0.442 | 22 | 0.010 (0.022) | 0.655 |
| NEFL | Alzheimer's disease, *FinnGen 2023* | Weighted Median | 9 | 0.156 (0.167) | 0.352 | 22 | 0.022 (0.013) | 0.099 |
| NEFL | Alzheimer's disease, *FinnGen 2023* | Maximum-likelihood | 9 | 0.091 (0.120) | 0.448 | 22 | 0.034 (0.010) | 0.001 |
| NEFL | Alzheimer's disease, *FinnGen 2023* | Weighted Mode | 9 | -1.921 (3.008) | 0.541 | 22 | 0.019 (0.014) | 0.183 |
| NEFL | Alzheimer's disease, *FinnGen 2023* | Leave-one-out | 9 | 0.086 (0.158) | 0.584 | 22 | 0.033 (0.015) | 0.024 |
| NEFL | All-cause dementia, *FinnGen 2023* | Inverse variant weighting | 9 | 0.024 (0.175) | 0.889 | 22 | 0.030 (0.017) | 0.080 |
| NEFL | All-cause dementia, *FinnGen 2023* | MR-Egger | 9 | -0.608 (0.630) | 0.366 | 22 | 0.019 (0.026) | 0.470 |
| NEFL | All-cause dementia, *FinnGen 2023* | Weighted Median | 9 | 0.235 (0.158) | 0.136 | 22 | 0.019 (0.015) | 0.189 |
| NEFL | All-cause dementia, *FinnGen 2023* | Maximum-likelihood | 9 | 0.027 (0.111) | 0.807 | 22 | 0.031 (0.012) | 0.010 |
| NEFL | All-cause dementia, *FinnGen 2023* | Weighted Mode | 9 | -1.852 (2.811) | 0.528 | - | - | - |
| NEFL | All-cause dementia, *FinnGen 2023* | Leave-one-out | 9 | 0.024 (0.175) | 0.889 | 22 | 0.030 (0.017) | 0.080 |
| NEFL | Vascular dementia, *FinnGen 2023* | Inverse variant weighting | 9 | 0.050 (0.269) | 0.854 | 4 | 0.035 (0.019) | 0.058 |
| NEFL | Vascular dementia, *FinnGen 2023* | MR-Egger | 9 | -1.541 (0.927) | 0.140 | 4 | 0.057 (0.018) | 0.090 |
| NEFL | Vascular dementia, *FinnGen 2023* | Weighted Median | 9 | -0.052 (0.343) | 0.880 | 4 | 0.035 (0.014) | 0.009 |
| NEFL | Vascular dementia, *FinnGen 2023* | Maximum-likelihood | 9 | 0.052 (0.263) | 0.844 | 4 | 0.036 (0.014) | 0.008 |
| NEFL | Vascular dementia, *FinnGen 2023* | Weighted Mode | 9 | -2.777 (5.385) | 0.620 | - | - | - |
| NEFL | Vascular dementia, *FinnGen 2023* | Leave-one-out | 9 | -0.050 (0.027) | 0.853 | 4 | 0.035 (0.019) | 0.058 |
| KIM1 | Alzheimer's disease, *Kunkle et al 2019* | Inverse variant weighting | 22 | -0.024 (0.032) | 0.451 | 19 | -0.007 (0.013) | 0.594 |
| KIM1 | Alzheimer's disease, *Kunkle et al 2019* | MR-Egger | 22 | -0.064 (0.045) | 0.174 | 19 | -0.017 (0.018) | 0.362 |

| | | | | | | | | |
|---|---|---|---|---|---|---|---|---|
| KIM1 (HAVCR1) | Alzheimer's disease, *Kunkle et al 2019* | Weighted Median | 22 | -0.042 (0.039) | 0.277 | 19 | -0.011 (0.012) | 0.367 |
| KIM1 (HAVCR1) | Alzheimer's disease, *Kunkle et al 2019* | Maximum-likelihood | 22 | -0.024 (0.032) | 0.452 | 19 | -0.007 (0.008) | 0.424 |
| KIM1 (HAVCR1) | Alzheimer's disease, *Kunkle et al 2019* | Weighted Mode | 22 | -0.039 (0.039) | 0.332 | 19 | -0.011 (0.012) | 0.377 |
| KIM1 (HAVCR1) | Alzheimer's disease, *Kunkle et al 2019* | Leave-one-out | 22 | -0.024 (0.032) | 0.451 | 19 | -0.007 (0.013) | 0.594 |
| KIM1 (HAVCR1) | Alzheimer's disease, *Bellenguez et al 2022* | Inverse variant weighting | 27 | -0.022 (0.026) | 0.393 | 59 | -0.011 (0.014) | 0.444 |
| KIM1 (HAVCR1) | Alzheimer's disease, *Bellenguez et al 2022* | MR-Egger | 27 | -0.034 (0.038) | 0.372 | 59 | -0.005 (0.024) | 0.825 |
| KIM1 (HAVCR1) | Alzheimer's disease, *Bellenguez et al 2022* | Weighted Median | 27 | -0.018 (0.023) | 0.432 | 59 | -0.004 (0.017) | 0.831 |
| KIM1 (HAVCR1) | Alzheimer's disease, *Bellenguez et al 2022* | Maximum-likelihood | 27 | -0.023 (0.018) | 0.196 | 59 | -0.011 (0.010) | 0.296 |
| KIM1 (HAVCR1) | Alzheimer's disease, *Bellenguez et al 2022* | Weighted Mode | 27 | -0.019 (0.022) | 0.391 | 59 | -0.013 (0.021) | 0.557 |
| KIM1 (HAVCR1) | Alzheimer's disease, *Bellenguez et al 2022* | Leave-one-out | 27 | -0.022 (0.026) | 0.393 | 59 | -0.011 (0.014) | 0.444 |
| KIM1 (HAVCR1) | Alzheimer's disease, *FinnGen 2023* | Inverse variant weighting | 26 | 0.002 (0.040) | 0.966 | 22 | 0.003 (0.014) | 0.839 |
| KIM1 (HAVCR1) | Alzheimer's disease, *FinnGen 2023* | MR-Egger | 26 | 0.002 (0.058) | 0.971 | 22 | -0.006 (0.021) | 0.789 |
| KIM1 (HAVCR1) | Alzheimer's disease, *FinnGen 2023* | Weighted Median | 26 | -0.034 (0.043) | 0.430 | 22 | -0.014 (0.012) | 0.265 |
| KIM1 (HAVCR1) | Alzheimer's disease, *FinnGen 2023* | Maximum-likelihood | 26 | 0.002 (0.028) | 0.951 | 22 | 0.003 (0.009) | 0.767 |
| KIM1 (HAVCR1) | Alzheimer's disease, *FinnGen 2023* | Weighted Mode | 26 | -0.013 (0.037) | 0.716 | 22 | -0.012 (0.012) | 0.334 |
| KIM1 (HAVCR1) | Alzheimer's disease, *FinnGen 2023* | Leave-one-out | 26 | 0.002 (0.040) | 0.966 | 22 | 0.003 (0.014) | 0.839 |
| KIM1 (HAVCR1) | All-cause dementia, *FinnGen 2023* | Inverse variant weighting | 26 | -0.001 (0.042) | 0.999 | 22 | 0.007 (0.017) | 0.659 |
| KIM1 (HAVCR1) | All-cause dementia, *FinnGen 2023* | MR-Egger | 26 | 0.014 (0.062) | 0.823 | 22 | -0.013 (0.025) | 0.613 |
| KIM1 (HAVCR1) | All-cause dementia, *FinnGen 2023* | Weighted Median | 26 | -0.068 (0.038) | 0.073 | 22 | -0.015 (0.014) | 0.271 |
| KIM1 (HAVCR1) | All-cause dementia, *FinnGen 2023* | Maximum-likelihood | 26 | -0.001 (0.025) | 0.998 | 22 | 0.008 (0.011) | 0.484 |
| KIM1 (HAVCR1) | All-cause dementia, *FinnGen 2023* | Weighted Mode | 26 | -0.053 (0.036) | 0.153 | 22 | -0.382 (1.619) | 0.816 |
| KIM1 (HAVCR1) | All-cause dementia, *FinnGen 2023* | Leave-one-out | 26 | -0.001 (0.042) | 0.999 | 22 | 0.007 (0.017) | 0.659 |
| KIM1 (HAVCR1) | Vascular dementia, *FinnGen 2023* | Inverse variant weighting | 26 | -0.067 (0.067) | 0.315 | 4 | -0.007 (0.012) | 0.590 |
| KIM1 (HAVCR1) | Vascular dementia, *FinnGen 2023* | MR-Egger | 26 | -0.010 (0.097) | 0.918 | 4 | 0.009 (0.017) | 0.658 |
| KIM1 (HAVCR1) | Vascular dementia, *FinnGen 2023* | Weighted Median | 26 | -0.142 (0.085) | 0.093 | 4 | -0.007 (0.012) | 0.594 |
| KIM1 (HAVCR1) | Vascular dementia, *FinnGen 2023* | Maximum-likelihood | 26 | -0.068 (0.061) | 0.270 | 4 | -0.007 (0.012) | 0.589 |
| KIM1 (HAVCR1) | Vascular dementia, *FinnGen 2023* | Weighted Mode | 26 | -0.074 (0.077) | 0.345 | - | - | - |
| KIM1 (HAVCR1) | Vascular dementia, *FinnGen 2023* | Leave-one-out | 26 | -0.067 (0.067) | 0.315 | 4 | -0.007 (0.012) | 0.590 |
| MMP12 | Alzheimer's disease, *Kunkle et al 2019* | Inverse variant weighting | 10 | -0.013 (0.039) | 0.745 | 19 | 0.016 (0.009) | 0.072 |
| MMP12 | Alzheimer's disease, *Kunkle et al 2019* | MR-Egger | 10 | 0.022 (0.045) | 0.649 | 19 | 0.027 (0.012) | 0.043 |
| MMP12 | Alzheimer's disease, *Kunkle et al 2019* | Weighted Median | 10 | -0.002 (0.032) | 0.954 | 19 | 0.025 (0.012) | 0.037 |
| MMP12 | Alzheimer's disease, *Kunkle et al 2019* | Maximum-likelihood | 10 | -0.013 (0.030) | 0.673 | 19 | 0.016 (0.009) | 0.072 |
| MMP12 | Alzheimer's disease, *Kunkle et al 2019* | Weighted Mode | 10 | -0.002 (0.030) | 0.944 | 19 | 0.026 (0.012) | 0.037 |
| MMP12 | Alzheimer's disease, *Kunkle et al 2019* | Leave-one-out | 10 | -0.013 (0.039) | 0.745 | 19 | 0.016 (0.009) | 0.072 |
| MMP12 | Alzheimer's disease, *Bellenguez et al 2022* | Inverse variant weighting | 8 | -0.017 (0.046) | 0.708 | 59 | 0.002 (0.016) | 0.916 |

| | | | | | | | | |
|---|---|---|---|---|---|---|---|---|
| MMP12 | Alzheimer's disease, *Bellenguez et al 2022* | MR-Egger | 8 | 0.003 (0.058) | 0.965 | 59 | 0.024 (0.028) | 0.396 |
| MMP12 | Alzheimer's disease, *Bellenguez et al 2022* | Weighted Median | 8 | -0.012 (0.018) | 0.491 | 59 | 0.017 (0.018) | 0.333 |
| MMP12 | Alzheimer's disease, *Bellenguez et al 2022* | Maximum-likelihood | 8 | -0.017 (0.017) | 0.312 | 59 | 0.002 (0.011) | 0.880 |
| MMP12 | Alzheimer's disease, *Bellenguez et al 2022* | Weighted Mode | 8 | -0.009 (0.018) | 0.629 | 59 | 0.017 (0.019) | 0.386 |
| MMP12 | Alzheimer's disease, *Bellenguez et al 2022* | Leave-one-out | 8 | -0.017 (0.046) | 0.708 | 59 | 0.002 (0.016) | 0.916 |
| MMP12 | Alzheimer's disease, *FinnGen 2023* | Inverse variant weighting | 10 | -0.006 (0.033) | 0.862 | 22 | 0.016 (0.011) | 0.134 |
| MMP12 | Alzheimer's disease, *FinnGen 2023* | MR-Egger | 10 | 0.014 (0.042) | 0.739 | 22 | 0.021 (0.017) | 0.230 |
| MMP12 | Alzheimer's disease, *FinnGen 2023* | Weighted Median | 10 | -0.002 (0.023) | 0.936 | 22 | 0.023 (0.013) | 0.082 |
| MMP12 | Alzheimer's disease, *FinnGen 2023* | Maximum-likelihood | 10 | -0.006 (0.022) | 0.789 | 22 | 0.016 (0.010) | 0.104 |
| MMP12 | Alzheimer's disease, *FinnGen 2023* | Weighted Mode | 10 | -0.004 (0.023) | 0.858 | 22 | 0.020 (0.014) | 0.177 |
| MMP12 | Alzheimer's disease, *FinnGen 2023* | Leave-one-out | 10 | -0.006 (0.033) | 0.862 | 22 | 0.016 (0.011) | 0.134 |
| MMP12 | All-cause dementia, *FinnGen 2023* | Inverse variant weighting | 10 | -0.004 (0.027) | 0.875 | 22 | 0.007 (0.015) | 0.625 |
| MMP12 | All-cause dementia, *FinnGen 2023* | MR-Egger | 10 | -0.001 (0.035) | 0.975 | 22 | 0.029 (0.022) | 0.195 |
| MMP12 | All-cause dementia, *FinnGen 2023* | Weighted Median | 10 | -0.006 (0.021) | 0.794 | 22 | 0.018 (0.015) | 0.235 |
| MMP12 | All-cause dementia, *FinnGen 2023* | Maximum-likelihood | 10 | -0.004 (0.020) | 0.833 | 22 | 0.007 (0.012) | 0.521 |
| MMP12 | All-cause dementia, *FinnGen 2023* | Weighted Mode | 10 | -0.006 (0.021) | 0.799 | - | - | - |
| MMP12 | All-cause dementia, *FinnGen 2023* | Leave-one-out | 10 | -0.004 (0.027) | 0.875 | 22 | 0.007 (0.015) | 0.625 |
| MMP12 | Vascular dementia, *FinnGen 2023* | Inverse variant weighting | 10 | -0.033 (0.048) | 0.490 | 4 | 0.006 (0.025) | 0.808 |
| MMP12 | Vascular dementia, *FinnGen 2023* | MR-Egger | 10 | -0.006 (0.059) | 0.926 | 4 | 0.035 (0.026) | 0.313 |
| MMP12 | Vascular dementia, *FinnGen 2023* | Weighted Median | 10 | -0.025 (0.049) | 0.607 | 4 | 0.007 (0.013) | 0.584 |
| MMP12 | Vascular dementia, *FinnGen 2023* | Maximum-likelihood | 10 | -0.034 (0.048) | 0.488 | 4 | 0.006 (0.013) | 0.629 |
| MMP12 | Vascular dementia, *FinnGen 2023* | Weighted Mode | 10 | -0.028 (0.050) | 0.586 | 4 | - | - |
| MMP12 | Vascular dementia, *FinnGen 2023* | Leave-one-out | 10 | -0.033 (0.048) | 0.490 | 4 | 0.006 (0.025) | 0.808 |
| EDA2R | Alzheimer's disease, *Kunkle et al 2019* | Inverse variant weighting | 16 | 0.106 (0.113) | 0.348 | 19 | -0.014 (0.008) | 0.085 |
| EDA2R | Alzheimer's disease, *Kunkle et al 2019* | MR-Egger | 16 | 0.127 (0.312) | 0.689 | 19 | -0.013 (0.012) | 0.269 |
| EDA2R | Alzheimer's disease, *Kunkle et al 2019* | Weighted Median | 16 | 0.163 (0.141) | 0.250 | 19 | -0.017 (0.012) | 0.143 |
| EDA2R | Alzheimer's disease, *Kunkle et al 2019* | Maximum-likelihood | 16 | 0.112 (0.102) | 0.399 | 19 | -0.014 (0.008) | 0.075 |
| EDA2R | Alzheimer's disease, *Kunkle et al 2019* | Weighted Mode | 16 | 0.196 (0.226) | 0.270 | 19 | -0.018 (0.011) | 0.134 |
| EDA2R | Alzheimer's disease, *Kunkle et al 2019* | Leave-one-out | 16 | 0.106 (0.113) | 0.348 | 19 | -0.014 (0.008) | 0.085 |
| EDA2R | Alzheimer's disease, *Bellenguez et al 2022* | Inverse variant weighting | 17 | 0.051 (0.070) | 0.471 | 59 | -0.015 (0.012) | 0.205 |
| EDA2R | Alzheimer's disease, *Bellenguez et al 2022* | MR-Egger | 17 | 0.166 (0.186) | 0.386 | 59 | -0.012 (0.021) | 0.568 |
| EDA2R | Alzheimer's disease, *Bellenguez et al 2022* | Weighted Median | 17 | 0.111 (0.084) | 0.185 | 59 | -0.012 (0.017) | 0.467 |
| EDA2R | Alzheimer's disease, *Bellenguez et al 2022* | Maximum-likelihood | 17 | 0.055 (0.054) | 0.309 | 59 | -0.015 (0.010) | 0.128 |
| EDA2R | Alzheimer's disease, *Bellenguez et al 2022* | Weighted Mode | 17 | 0.237 (0.169) | 0.180 | 59 | -0.034 (0.024) | 0.155 |
| EDA2R | Alzheimer's disease, *Bellenguez et al 2022* | Leave-one-out | 17 | 0.051 (0.070) | 0.471 | 59 | -0.015 (0.012) | 0.205 |

| | | | | | | | | |
|---|---|---|---|---|---|---|---|---|
| EDA2R | Alzheimer's disease, *FinnGen 2023* | Inverse variant weighting | 16 | 0.259 (0.096) | 0.007 | 22 | 0.013 (0.049) | 0.049 |
| EDA2R | Alzheimer's disease, *FinnGen 2023* | MR-Egger | 16 | 0.221 (0.244) | 0.382 | 22 | 0.020 (0.432) | 0.432 |
| EDA2R | Alzheimer's disease, *FinnGen 2023* | Weighted Median | 16 | 0.194 (0.122) | 0.114 | 22 | 0.012 (0.016) | 0.016 |
| EDA2R | Alzheimer's disease, *FinnGen 2023* | Maximum-likelihood | 16 | 0.267 (0.085) | 0.002 | 22 | 0.009 (0.006) | 0.006 |
| EDA2R | Alzheimer's disease, *FinnGen 2023* | Weighted Mode | 16 | 0.238 (0.169) | 0.181 | 22 | 0.012 (0.039) | 0.039 |
| EDA2R | Alzheimer's disease, *FinnGen 2023* | Leave-one-out | 16 | 0.259 (0.096) | 0.007 | 22 | 0.013 (0.049) | 0.049 |
| EDA2R | All-cause dementia, *FinnGen 2023* | Inverse variant weighting | 16 | 0.232 (0.110) | 0.035 | 22 | -0.032 (0.012) | 0.007 |
| EDA2R | All-cause dementia, *FinnGen 2023* | MR-Egger | 16 | 0.457 (0.272) | 0.115 | 22 | -0.030 (0.018) | 0.114 |
| EDA2R | All-cause dementia, *FinnGen 2023* | Weighted Median | 16 | 0.246 (0.111) | 0.027 | 22 | -0.036 (0.013) | 0.006 |
| EDA2R | All-cause dementia, *FinnGen 2023* | Maximum-likelihood | 16 | 0.241 (0.078) | 0.002 | 22 | -0.032 (0.011) | 0.003 |
| EDA2R | All-cause dementia, *FinnGen 2023* | Weighted Mode | 16 | 0.329 (0.143) | 0.036 | 22 | -0.570 (1.723) | 0.744 |
| EDA2R | All-cause dementia, *FinnGen 2023* | Leave-one-out | 16 | 0.232 (0.110) | 0.035 | 22 | -0.032 (0.012) | 0.007 |
| EDA2R | Vascular dementia, *FinnGen 2023* | Inverse variant weighting | 16 | 0.127 (0.194) | 0.512 | 4 | -0.037 (0.020) | 0.058 |
| EDA2R | Vascular dementia, *FinnGen 2023* | MR-Egger | 16 | -0.390 (0.474) | 0.424 | 4 | -0.013 (0.017) | 0.513 |
| EDA2R | Vascular dementia, *FinnGen 2023* | Weighted Median | 16 | 0.083 (0.247) | 0.737 | 4 | -0.036 (0.012) | 0.003 |
| EDA2R | Vascular dementia, *FinnGen 2023* | Maximum-likelihood | 16 | 0.131 (0.186) | 0.480 | 4 | -0.038 (0.012) | 0.002 |
| EDA2R | Vascular dementia, *FinnGen 2023* | Weighted Mode | 16 | -0.014 (0.317) | 0.966 | - | - | - |
| EDA2R | Vascular dementia, *FinnGen 2023* | Leave-one-out | 16 | 0.127 (0.194) | 0.512 | 4 | -0.037 (0.020) | 0.058 |

*MR, Mendelian randomization; SNP, single nucleotide polymorphism; SE, standard error.*

**Supplementary Table 4. Two sample cis-Mendelian randomization between proteins and dementia.**

| Gene name | Outcome trait | MR methods | Cis-MR (protein → dementia) Main analysis ($P < 5 \times 10^{-5}$, $R^2 < 0.001$) | | | Cis-MR (protein → dementia) Sensitivity analysis ($P < 5 \times 10^{-5}$, $R^2 < 0.01$) | | |
|---|---|---|---|---|---|---|---|---|
| | | | No of SNP | Slope (SE) | P value | No of SNP | Slope (SE) | P value |
| NEFL | Alzheimer's disease, *Kunkle et al 2019* | Inverse variant weighting | 2 | 0.820 (0.485) | 0.091 | 2 | 0.820 (0.485) | 0.091 |
| NEFL | Alzheimer's disease, *Kunkle et al 2019* | MR-Egger | - | - | - | - | - | - |
| NEFL | Alzheimer's disease, *Kunkle et al 2019* | Weighted Median | - | - | - | - | - | - |
| NEFL | Alzheimer's disease, *Kunkle et al 2019* | Maximum-likelihood | 2 | 0.857 (0.516) | 0.097 | 2 | 0.857 (0.516) | 0.097 |
| NEFL | Alzheimer's disease, *Kunkle et al 2019* | Weighted Mode | - | - | - | - | - | - |
| NEFL | Alzheimer's disease, *Kunkle et al 2019* | Leave-one-out | 2 | 0.820 (0.485) | 0.091 | 2 | 0.820 (0.485) | 0.091 |
| NEFL | Alzheimer's disease, *Bellenguez et al 2022* | Inverse variant weighting | 2 | 0.303 (0.372) | 0.414 | 2 | 0.303 (0.372) | 0.414 |
| NEFL | Alzheimer's disease, *Bellenguez et al 2022* | MR-Egger | - | - | - | - | - | - |
| NEFL | Alzheimer's disease, *Bellenguez et al 2022* | Weighted Median | - | - | - | - | - | - |
| NEFL | Alzheimer's disease, *Bellenguez et al 2022* | Maximum-likelihood | 2 | 0.331 (0.277) | 0.232 | 2 | 0.331 (0.277) | 0.232 |
| NEFL | Alzheimer's disease, *Bellenguez et al 2022* | Weighted Mode | - | - | - | - | - | - |
| NEFL | Alzheimer's disease, *Bellenguez et al 2022* | Leave-one-out | 2 | 0.303 (0.372) | 0.414 | 2 | 0.303 (0.372) | 0.414 |
| NEFL | Alzheimer's disease, *FinnGen 2023* | Inverse variant weighting | 2 | 0.620 (0.392) | 0.114 | 2 | 0.620 (0.392) | 0.114 |
| NEFL | Alzheimer's disease, *FinnGen 2023* | MR-Egger | - | - | - | - | - | - |
| NEFL | Alzheimer's disease, *FinnGen 2023* | Weighted Median | - | - | - | - | - | - |
| NEFL | Alzheimer's disease, *FinnGen 2023* | Maximum-likelihood | 2 | 0.621 (0.414) | 0.133 | 2 | 0.621 (0.414) | 0.133 |
| NEFL | Alzheimer's disease, *FinnGen 2023* | Weighted Mode | - | - | - | - | - | - |
| NEFL | Alzheimer's disease, *FinnGen 2023* | Leave-one-out | 2 | 0.620 (0.392) | 0.114 | 2 | 0.620 (0.392) | 0.114 |
| NEFL | All-cause dementia, *FinnGen 2023* | Inverse variant weighting | 2 | 0.161 (0.358) | 0.652 | 2 | 0.161 (0.358) | 0.652 |
| NEFL | All-cause dementia, *FinnGen 2023* | MR-Egger | - | - | - | - | - | - |
| NEFL | All-cause dementia, *FinnGen 2023* | Weighted Median | - | - | - | - | - | - |
| NEFL | All-cause dementia, *FinnGen 2023* | Maximum-likelihood | 2 | 0.162 (0.360) | 0.653 | 2 | 0.162 (0.360) | 0.653 |
| NEFL | All-cause dementia, *FinnGen 2023* | Weighted Mode | - | - | - | - | - | - |
| NEFL | All-cause dementia, *FinnGen 2023* | Leave-one-out | 2 | 0.161 (0.358) | 0.652 | 2 | 0.161 (0.358) | 0.652 |
| NEFL | Vascular dementia, *FinnGen 2023* | Inverse variant weighting | 2 | -0.222 (1.166) | 0.849 | 2 | -0.222 (1.166) | 0.849 |
| NEFL | Vascular dementia, *FinnGen 2023* | MR-Egger | - | - | - | - | - | - |
| NEFL | Vascular dementia, *FinnGen 2023* | Weighted Median | - | - | - | - | - | - |
| NEFL | Vascular dementia, *FinnGen 2023* | Maximum-likelihood | 2 | -0.235 (0.895) | 0.793 | 2 | -0.235 (0.895) | 0.793 |
| NEFL | Vascular dementia, *FinnGen 2023* | Weighted Mode | - | - | - | - | - | - |
| NEFL | Vascular dementia, *FinnGen 2023* | Leave-one-out | 2 | -0.222 (1.166) | 0.849 | 2 | -0.222 (1.166) | 0.849 |
| KIM1 (HAVCR1) | Alzheimer's disease, *Kunkle et al 2019* | Inverse variant weighting | 4 | -0.030 (0.042) | 0.473 | 9 | -0.006 (0.039) | 0.882 |

| | | | | | | | | |
|---|---|---|---|---|---|---|---|---|
| KIM1 (HAVCR1) | Alzheimer's disease, *Kunkle et al 2019* | MR-Egger | 4 | -0.274 (0.114) | 0.138 | 9 | 0.021 (0.101) | 0.837 |
| KIM1 (HAVCR1) | Alzheimer's disease, *Kunkle et al 2019* | Weighted Median | 4 | -0.025 (0.035) | 0.471 | 9 | -0.012 (0.036) | 0.731 |
| KIM1 (HAVCR1) | Alzheimer's disease, *Kunkle et al 2019* | Maximum-likelihood | 4 | -0.030 (0.032) | 0.354 | 9 | -0.006 (0.032) | 0.858 |
| KIM1 (HAVCR1) | Alzheimer's disease, *Kunkle et al 2019* | Weighted Mode | 4 | -0.008 (0.040) | 0.853 | 9 | -0.011 (0.036) | 0.767 |
| KIM1 (HAVCR1) | Alzheimer's disease, *Kunkle et al 2019* | Leave-one-out | 4 | -0.030 (0.042) | 0.473 | 9 | -0.006 (0.039) | 0.882 |
| KIM1 (HAVCR1) | Alzheimer's disease, *Bellenguez et al 2022* | Inverse variant weighting | 4 | 0.004 (0.017) | 0.814 | 10 | 0.003 (0.015) | 0.858 |
| KIM1 (HAVCR1) | Alzheimer's disease, *Bellenguez et al 2022* | MR-Egger | 4 | 0.040 (0.055) | 0.542 | 10 | 0.019 (0.030) | 0.556 |
| KIM1 (HAVCR1) | Alzheimer's disease, *Bellenguez et al 2022* | Weighted Median | 4 | 0.006 (0.020) | 0.761 | 10 | 0.005 (0.019) | 0.801 |
| KIM1 (HAVCR1) | Alzheimer's disease, *Bellenguez et al 2022* | Maximum-likelihood | 4 | 0.004 (0.017) | 0.814 | 10 | 0.003 (0.015) | 0.859 |
| KIM1 (HAVCR1) | Alzheimer's disease, *Bellenguez et al 2022* | Weighted Mode | 4 | 0.006 (0.019) | 0.766 | 10 | 0.006 (0.017) | 0.737 |
| KIM1 (HAVCR1) | Alzheimer's disease, *Bellenguez et al 2022* | Leave-one-out | 4 | 0.004 (0.017) | 0.814 | 10 | 0.003 (0.015) | 0.858 |
| KIM1 (HAVCR1) | Alzheimer's disease, *FinnGen 2023* | Inverse variant weighting | 4 | -0.028 (0.039) | 0.471 | 10 | -0.038 (0.023) | 0.090 |
| KIM1 (HAVCR1) | Alzheimer's disease, *FinnGen 2023* | MR-Egger | 4 | -0.135 (0.088) | 0.264 | 10 | -0.102 (0.041) | 0.037 |
| KIM1 (HAVCR1) | Alzheimer's disease, *FinnGen 2023* | Weighted Median | 4 | -0.047 (0.027) | 0.078 | 10 | -0.046 (0.027) | 0.086 |
| KIM1 (HAVCR1) | Alzheimer's disease, *FinnGen 2023* | Maximum-likelihood | 4 | -0.028 (0.025) | 0.267 | 10 | -0.038 (0.023) | 0.091 |
| KIM1 (HAVCR1) | Alzheimer's disease, *FinnGen 2023* | Weighted Mode | 4 | -0.050 (0.026) | 0.154 | 10 | -0.053 (0.026) | 0.072 |
| KIM1 (HAVCR1) | Alzheimer's disease, *FinnGen 2023* | Leave-one-out | 4 | -0.028 (0.039) | 0.471 | 10 | -0.038 (0.023) | 0.090 |
| KIM1 (HAVCR1) | All-cause dementia, *FinnGen 2023* | Inverse variant weighting | 4 | -0.025 (0.037) | 0.504 | 10 | -0.026 (0.021) | 0.219 |
| KIM1 (HAVCR1) | All-cause dementia, *FinnGen 2023* | MR-Egger | 4 | -0.105 (0.099) | 0.401 | 10 | -0.094 (0.037) | 0.036 |
| KIM1 (HAVCR1) | All-cause dementia, *FinnGen 2023* | Weighted Median | 4 | -0.046 (0.026) | 0.075 | 10 | -0.049 (0.024) | 0.043 |
| KIM1 (HAVCR1) | All-cause dementia, *FinnGen 2023* | Maximum-likelihood | 4 | -0.025 (0.023) | 0.278 | 10 | -0.026 (0.021) | 0.218 |
| KIM1 (HAVCR1) | All-cause dementia, *FinnGen 2023* | Weighted Mode | 4 | -0.044 (0.024) | 0.157 | 10 | -0.048 (0.022) | 0.060 |
| KIM1 (HAVCR1) | All-cause dementia, *FinnGen 2023* | Leave-one-out | 4 | -0.025 (0.037) | 0.504 | 10 | -0.026 (0.021) | 0.219 |
| KIM1 (HAVCR1) | Vascular dementia, *FinnGen 2023* | Inverse variant weighting | 4 | 0.001 (0.057) | 0.986 | 10 | 0.009 (0.058) | 0.878 |
| KIM1 (HAVCR1) | Vascular dementia, *FinnGen 2023* | MR-Egger | 4 | -0.028 (0.172) | 0.885 | 10 | -0.002 (0.112) | 0.983 |
| KIM1 (HAVCR1) | Vascular dementia, *FinnGen 2023* | Weighted Median | 4 | -0.017 (0.061) | 0.785 | 10 | -0.001 (0.062) | 0.993 |
| KIM1 (HAVCR1) | Vascular dementia, *FinnGen 2023* | Maximum-likelihood | 4 | 0.001 (0.057) | 0.986 | 10 | 0.009 (0.051) | 0.863 |
| KIM1 (HAVCR1) | Vascular dementia, *FinnGen 2023* | Weighted Mode | 4 | -0.025 (0.062) | 0.713 | 10 | -0.028 (0.056) | 0.630 |
| KIM1 (HAVCR1) | Vascular dementia, *FinnGen 2023* | Leave-one-out | 4 | 0.001 (0.057) | 0.986 | 10 | 0.009 (0.058) | 0.878 |
| MMP12 | Alzheimer's disease, *Kunkle et al 2019* | Inverse variant weighting | 4 | -0.008 (0.031) | 0.787 | 9 | -0.003 (0.027) | 0.919 |
| MMP12 | Alzheimer's disease, *Kunkle et al 2019* | MR-Egger | 4 | 0.010 (0.044) | 0.838 | 9 | -0.011 (0.045) | 0.819 |
| MMP12 | Alzheimer's disease, *Kunkle et al 2019* | Weighted Median | 4 | -0.003 (0.031) | 0.935 | 9 | 0.000 (0.030) | 0.989 |
| MMP12 | Alzheimer's disease, *Kunkle et al 2019* | Maximum-likelihood | 4 | -0.008 (0.031) | 0.787 | 9 | -0.003 (0.027) | 0.919 |
| MMP12 | Alzheimer's disease, *Kunkle et al 2019* | Weighted Mode | 4 | 0.000 (0.032) | 0.995 | 9 | -0.002 (0.028) | 0.950 |
| MMP12 | Alzheimer's disease, *Kunkle et al 2019* | Leave-one-out | 4 | -0.008 (0.031) | 0.787 | 9 | -0.003 (0.027) | 0.919 |

| | | | | | | | | |
|---|---|---|---|---|---|---|---|---|
| MMP12 | Alzheimer's disease, *Bellenguez et al 2022* | Inverse variant weighting | 3 | -0.011 (0.018) | 0.528 | 9 | -0.009 (0.015) | 0.546 |
| MMP12 | Alzheimer's disease, *Bellenguez et al 2022* | MR-Egger | 3 | -0.019 (0.025) | 0.580 | 9 | -0.009 (0.025) | 0.746 |
| MMP12 | Alzheimer's disease, *Bellenguez et al 2022* | Weighted Median | 3 | -0.012 (0.018) | 0.487 | 9 | -0.011 (0.017) | 0.512 |
| MMP12 | Alzheimer's disease, *Bellenguez et al 2022* | Maximum-likelihood | 3 | -0.011 (0.018) | 0.528 | 9 | -0.009 (0.015) | 0.545 |
| MMP12 | Alzheimer's disease, *Bellenguez et al 2022* | Weighted Mode | 3 | -0.013 (0.018) | 0.562 | 9 | -0.011 (0.017) | 0.514 |
| MMP12 | Alzheimer's disease, *Bellenguez et al 2022* | Leave-one-out | 3 | -0.011 (0.018) | 0.528 | 9 | -0.009 (0.015) | 0.546 |
| MMP12 | Alzheimer's disease, *FinnGen 2023* | Inverse variant weighting | 4 | -0.001 (0.022) | 0.981 | 9 | 0.010 (0.020) | 0.620 |
| MMP12 | Alzheimer's disease, *FinnGen 2023* | MR-Egger | 4 | -0.015 (0.034) | 0.701 | 9 | -0.003 (0.033) | 0.922 |
| MMP12 | Alzheimer's disease, *FinnGen 2023* | Weighted Median | 4 | -0.002 (0.022) | 0.935 | 9 | 0.005 (0.021) | 0.803 |
| MMP12 | Alzheimer's disease, *FinnGen 2023* | Maximum-likelihood | 4 | -0.001 (0.022) | 0.981 | 9 | 0.010 (0.020) | 0.620 |
| MMP12 | Alzheimer's disease, *FinnGen 2023* | Weighted Mode | 4 | -0.003 (0.024) | 0.924 | 9 | 0.002 (0.022) | 0.938 |
| MMP12 | Alzheimer's disease, *FinnGen 2023* | Leave-one-out | 4 | -0.001 (0.022) | 0.981 | 9 | 0.010 (0.020) | 0.620 |
| MMP12 | All-cause dementia, *FinnGen 2023* | Inverse variant weighting | 4 | -0.002 (0.020) | 0.923 | 9 | 0.010 (0.019) | 0.586 |
| MMP12 | All-cause dementia, *FinnGen 2023* | MR-Egger | 4 | -0.028 (0.031) | 0.462 | 9 | -0.018 (0.031) | 0.568 |
| MMP12 | All-cause dementia, *FinnGen 2023* | Weighted Median | 4 | -0.006 (0.021) | 0.788 | 9 | 0.001 (0.020) | 0.942 |
| MMP12 | All-cause dementia, *FinnGen 2023* | Maximum-likelihood | 4 | -0.002 (0.020) | 0.923 | 9 | 0.010 (0.019) | 0.584 |
| MMP12 | All-cause dementia, *FinnGen 2023* | Weighted Mode | 4 | -0.006 (0.021) | 0.808 | 9 | 0.000 (0.020) | 0.992 |
| MMP12 | All-cause dementia, *FinnGen 2023* | Leave-one-out | 4 | -0.002 (0.020) | 0.923 | 9 | 0.010 (0.019) | 0.586 |
| MMP12 | Vascular dementia, *FinnGen 2023* | Inverse variant weighting | 4 | -0.033 (0.049) | 0.499 | 9 | 0.004 (0.045) | 0.930 |
| MMP12 | Vascular dementia, *FinnGen 2023* | MR-Egger | 4 | 0.025 (0.076) | 0.771 | 9 | -0.071 (0.075) | 0.371 |
| MMP12 | Vascular dementia, *FinnGen 2023* | Weighted Median | 4 | -0.025 (0.051) | 0.627 | 9 | -0.010 (0.048) | 0.830 |
| MMP12 | Vascular dementia, *FinnGen 2023* | Maximum-likelihood | 4 | -0.033 (0.049) | 0.498 | 9 | 0.004 (0.045) | 0.930 |
| MMP12 | Vascular dementia, *FinnGen 2023* | Weighted Mode | 4 | -0.022 (0.049) | 0.686 | 9 | -0.018 (0.051) | 0.736 |
| MMP12 | Vascular dementia, *FinnGen 2023* | Leave-one-out | 4 | -0.033 (0.049) | 0.499 | 9 | 0.004 (0.045) | 0.930 |

*MR, Mendelian randomization; SNP, single nucleotide polymorphism; SE, standard error.*

**Supplementary Table 5. Enrichment analysis for the identified proteins.**

| Term | P value | FDR-corrected P value | Genes | dataset |
|---|---|---|---|---|
| Calcium Ion Binding (GO:0005509) | 0.214 | 0.242 | MMP12 | GO_MF |
| Core Promoter Sequence-Specific DNA Binding (GO:0001046) | 0.029 | 0.144 | MMP12 | GO_MF |
| DNA Binding (GO:0003677) | 0.071 | 0.152 | MMP12 | GO_MF |
| Endopeptidase Activity (GO:0004175) | 0.201 | 0.242 | MMP12 | GO_MF |
| Metal Ion Binding (GO:0046872) | 0.249 | 0.249 | MMP12 | GO_MF |
| Metalloendopeptidase Activity (GO:0004222) | 0.112 | 0.186 | MMP12 | GO_MF |
| Metallopeptidase Activity (GO:0008237) | 0.138 | 0.207 | MMP12 | GO_MF |
| Protein Serine/Threonine Kinase Activity (GO:0004674) | 0.029 | 0.144 | RPS6KB1 | GO_MF |
| Protein Serine/Threonine/Tyrosine Kinase Activity (GO:0004712) | 0.014 | 0.144 | RPS6KB1 | GO_MF |
| Sequence-Specific DNA Binding (GO:0043565) | 0.057 | 0.152 | MMP12 | GO_MF |
| Serine-Type Endopeptidase Activity (GO:0004252) | 0.071 | 0.152 | MMP12 | GO_MF |
| Serine-Type Peptidase Activity (GO:0008236) | 0.085 | 0.159 | MMP12 | GO_MF |
| Transcription Cis-Regulatory Region Binding (GO:0000976) | 0.043 | 0.152 | MMP12 | GO_MF |
| Transition Metal Ion Binding (GO:0046914) | 0.226 | 0.242 | MMP12 | GO_MF |
| Zinc Ion Binding (GO:0008270) | 0.201 | 0.242 | MMP12 | GO_MF |
| Brain - Cortex Female 50-59 Up | 0.014 | 0.042 | NEFL | GTEx |
| Brain - Cortex Male 30-39 Up | 0.014 | 0.042 | NEFL | GTEx |
| Brain - Cortex Male 70-79 Up | 0.014 | 0.042 | NEFL | GTEx |
| Brain - Frontal Cortex (BA9) Female 40-49 Up | 0.029 | 0.042 | NEFL | GTEx |
| Brain - Frontal Cortex (BA9) Female 50-59 Up | 0.029 | 0.042 | NEFL | GTEx |
| Brain - Frontal Cortex (BA9) Female 60-69 Up | 0.029 | 0.042 | NEFL | GTEx |
| Brain - Frontal Cortex (BA9) Male 20-29 Up | 0.014 | 0.042 | NEFL | GTEx |
| Brain - Frontal Cortex (BA9) Male 40-49 Up | 0.014 | 0.042 | NEFL | GTEx |
| Brain - Frontal Cortex (BA9) Male 50-59 Up | 0.014 | 0.042 | NEFL | GTEx |
| Brain - Frontal Cortex (BA9) Male 60-69 Up | 0.014 | 0.042 | NEFL | GTEx |
| Brain - Frontal Cortex (BA9) Male 70-79 Up | 0.014 | 0.042 | NEFL | GTEx |
| Brain - Hypothalamus Male 20-29 Up | 0.029 | 0.042 | NEFL | GTEx |
| Brain - Substantia Nigra Female 50-59 Up | 0.029 | 0.042 | NEFL | GTEx |
| Brain - Substantia Nigra Male 20-29 Up | 0.029 | 0.042 | NEFL | GTEx |
| Brain - Substantia Nigra Male 50-59 Up | 0.029 | 0.042 | NEFL | GTEx |
| Cells - Cultured Fibroblasts Female 40-49 Up | 0.057 | 0.062 | EDA2R | GTEx |
| Cells - Cultured Fibroblasts Female 50-59 Up | 0.057 | 0.062 | EDA2R | GTEx |
| Cells - Cultured Fibroblasts Male 20-29 Up | 0.071 | 0.071 | EDA2R | GTEx |

| | | | | |
|---|---|---|---|---|
| Cells - Cultured Fibroblasts Male 40-49 Up | 0.057 | 0.062 | EDA2R | GTEx |
| Cells - Cultured Fibroblasts Male 50-59 Up | 0.057 | 0.062 | EDA2R | GTEx |
| Cells - Cultured Fibroblasts Male 70-79 Up | 0.071 | 0.071 | EDA2R | GTEx |
| Kidney - Cortex Female 60-69 Up | 0.014 | 0.042 | HAVCR1 | GTEx |
| Kidney - Cortex Male 30-39 Up | 0.029 | 0.042 | HAVCR1 | GTEx |
| Kidney - Cortex Male 70-79 Up | 0.029 | 0.042 | HAVCR1 | GTEx |
| Minor Salivary Gland Male 20-29 Up | 0.057 | 0.062 | MMP12 | GTEx |
| Small Intestine - Terminal Ileum Female 20-29 Up | 0.043 | 0.059 | MMP12 | GTEx |
| Bosutinib | 0.125 | 0.125 | RPS6KB1 | IDG_DT |
| Crizotinib | 0.125 | 0.125 | RPS6KB1 | IDG_DT |
| Fedratinib | 0.112 | 0.125 | RPS6KB1 | IDG_DT |
| Midostaurin | 0.125 | 0.125 | RPS6KB1 | IDG_DT |
| Nintedanib | 0.125 | 0.125 | RPS6KB1 | IDG_DT |
| Quercetin | 0.098 | 0.125 | MMP12 | IDG_DT |
| Ruboxistaurin | 0.029 | 0.125 | RPS6KB1 | IDG_DT |
| Sunitinib | 0.125 | 0.125 | RPS6KB1 | IDG_DT |
| Vandetanib | 0.085 | 0.125 | RPS6KB1 | IDG_DT |
| AMPK signaling pathway | 0.057 | 0.083 | RPS6KB1 | KEGG |
| Acute myeloid leukemia | 0.014 | 0.083 | RPS6KB1 | KEGG |
| Amyotrophic lateral sclerosis | 0.029 | 0.083 | NEFL | KEGG |
| Apelin signaling pathway | 0.014 | 0.083 | RPS6KB1 | KEGG |
| Autophagy | 0.029 | 0.083 | RPS6KB1 | KEGG |
| Breast cancer | 0.043 | 0.083 | RPS6KB1 | KEGG |
| Chemical carcinogenesis | 0.057 | 0.083 | RPS6KB1 | KEGG |
| Choline metabolism in cancer | 0.029 | 0.083 | RPS6KB1 | KEGG |
| Colorectal cancer | 0.029 | 0.083 | RPS6KB1 | KEGG |
| Cytokine-cytokine receptor interaction | 0.371 | 0.371 | EDA2R | KEGG |
| ErbB signaling pathway | 0.057 | 0.083 | RPS6KB1 | KEGG |
| Fc gamma R-mediated phagocytosis | 0.043 | 0.083 | RPS6KB1 | KEGG |
| Gastric cancer | 0.071 | 0.091 | RPS6KB1 | KEGG |
| HIF-1 signaling pathway | 0.057 | 0.083 | RPS6KB1 | KEGG |
| Hepatocellular carcinoma | 0.029 | 0.083 | RPS6KB1 | KEGG |
| Human cytomegalovirus infection | 0.071 | 0.091 | RPS6KB1 | KEGG |
| Human immunodeficiency virus 1 infection | 0.057 | 0.083 | RPS6KB1 | KEGG |
| Human papillomavirus infection | 0.057 | 0.083 | RPS6KB1 | KEGG |

| | | | | |
|---|---|---|---|---|
| Insulin resistance | 0.043 | 0.083 | RPS6KB1 | KEGG |
| Insulin signaling pathway | 0.029 | 0.083 | RPS6KB1 | KEGG |
| Longevity regulating pathway | 0.043 | 0.083 | RPS6KB1 | KEGG |
| NF-kappa B signaling pathway | 0.085 | 0.100 | EDA2R | KEGG |
| PD-L1 expression and PD-1 checkpoint pathway in cancer | 0.057 | 0.083 | RPS6KB1 | KEGG |
| PI3K-Akt signaling pathway | 0.226 | 0.233 | RPS6KB1 | KEGG |
| Pancreatic cancer | 0.014 | 0.083 | RPS6KB1 | KEGG |
| Pathways in cancer | 0.189 | 0.202 | RPS6KB1 | KEGG |
| Pathways of neurodegeneration | 0.085 | 0.100 | NEFL | KEGG |
| Proteoglycans in cancer | 0.098 | 0.108 | RPS6KB1 | KEGG |
| Shigellosis | 0.071 | 0.091 | RPS6KB1 | KEGG |
| TGF-beta signaling pathway | 0.098 | 0.108 | RPS6KB1 | KEGG |
| Thermogenesis | 0.057 | 0.083 | RPS6KB1 | KEGG |
| mTOR signaling pathway | 0.057 | 0.083 | RPS6KB1 | KEGG |
| LY2584702 Down | 0.014 | 0.014 | RPS6KB1 | PDA |
| Activation Of NMDA Receptors and Postsynaptic Events R-HSA-442755 | 0.043 | 0.076 | NEFL | REACTOME |
| Assembly And Cell Surface Presentation of NMDA Receptors R-HSA-9609736 | 0.014 | 0.060 | NEFL | REACTOME |
| CREB1 Phosphorylation Thru NMDA Receptor-Mediated Activation of RAS Signaling R-HSA-442742 | 0.014 | 0.060 | NEFL | REACTOME |
| Cardiac Conduction R-HSA-5576891 | 0.014 | 0.060 | MMP12 | REACTOME |
| Collagen Degradation R-HSA-1442490 | 0.029 | 0.071 | MMP12 | REACTOME |
| Cytokine Signaling in Immune System R-HSA-1280215 | 0.339 | 0.358 | EDA2R | REACTOME |
| Degradation Of Extracellular Matrix R-HSA-1474228 | 0.085 | 0.125 | MMP12 | REACTOME |
| Disease R-HSA-1643685 | 0.381 | 0.381 | HAVCR1 | REACTOME |
| Early SARS-CoV-2 Infection Events R-HSA-9772572 | 0.014 | 0.060 | HAVCR1 | REACTOME |
| Extracellular Matrix Organization R-HSA-1474244 | 0.189 | 0.226 | MMP12 | REACTOME |
| Immune System R-HSA-168256 | 0.196 | 0.226 | MMP12; EDA2R | REACTOME |
| Infectious Disease R-HSA-5663205 | 0.189 | 0.226 | HAVCR1 | REACTOME |
| Innate Immune System R-HSA-168249 | 0.339 | 0.358 | MMP12 | REACTOME |
| Long-term Potentiation R-HSA-9620244 | 0.029 | 0.071 | NEFL | REACTOME |
| MAPK Family Signaling Cascades R-HSA-5683057 | 0.176 | 0.226 | NEFL | REACTOME |
| MAPK1/MAPK3 Signaling R-HSA-5684996 | 0.176 | 0.226 | NEFL | REACTOME |
| MTOR Signaling R-HSA-165159 | 0.043 | 0.076 | RPS6KB1 | REACTOME |
| Metabolism Of Angiotensinogen to Angiotensin R-HSA-2022377 | 0.029 | 0.071 | MMP12 | REACTOME |
| Metabolism Of Proteins R-HSA-392499 | 0.360 | 0.370 | MMP12 | REACTOME |
| Muscle Contraction R-HSA-397014 | 0.014 | 0.060 | MMP12 | REACTOME |

| | | | | |
|---|---|---|---|---|
| Negative Regulation of NMDA Receptor-Mediated Neuronal Transmission R-HSA-9617324 | 0.014 | 0.060 | NEFL | REACTOME |
| Neuronal System R-HSA-112316 | 0.071 | 0.114 | NEFL | REACTOME |
| Neurotransmitter Receptors and Postsynaptic Signal Transmission R-HSA-112314 | 0.043 | 0.076 | NEFL | REACTOME |
| Neutrophil Degranulation R-HSA-6798695 | 0.214 | 0.239 | MMP12 | REACTOME |
| Peptide Hormone Metabolism R-HSA-2980736 | 0.043 | 0.076 | MMP12 | REACTOME |
| Physiological Factors R-HSA-5578768 | 0.014 | 0.060 | MMP12 | REACTOME |
| Post NMDA Receptor Activation Events R-HSA-438064 | 0.043 | 0.076 | NEFL | REACTOME |
| RAF/MAP Kinase Cascade R-HSA-5673001 | 0.164 | 0.226 | NEFL | REACTOME |
| Ras Activation Upon Ca2+ Influx Thru NMDA Receptor R-HSA-442982 | 0.014 | 0.060 | NEFL | REACTOME |
| SARS-CoV Infections R-HSA-9679506 | 0.029 | 0.071 | HAVCR1 | REACTOME |
| SARS-CoV-2 Infection R-HSA-9694516 | 0.029 | 0.071 | HAVCR1 | REACTOME |
| Signal Transduction R-HSA-162582 | 0.190 | 0.226 | RPS6KB1; NEFL | REACTOME |
| TNFR2 Non-Canonical NF-kB Pathway R-HSA-5668541 | 0.085 | 0.125 | EDA2R | REACTOME |
| TNFs Bind Their Physiological Receptors R-HSA-5669034 | 0.029 | 0.071 | EDA2R | REACTOME |
| Transmission Across Chemical Synapses R-HSA-112315 | 0.057 | 0.096 | NEFL | REACTOME |
| Unblocking Of NMDA Receptors, Glutamate Binding and Activation R-HSA-438066 | 0.014 | 0.060 | NEFL | REACTOME |
| mTORC1-mediated Signaling R-HSA-166208 | 0.043 | 0.076 | RPS6KB1 | REACTOME |

*GO_MF, Gene Ontology – Molecular Function; GTEx, Genotype-Tissue Expression; IDG_DT, Illuminating the Druggable Genome – drug target; PDA, Proteomics Drug Atlas; KEGG, Kyoto Encyclopedia of Genes and Genomes; REACTOME, Reactome Pathway Database.*

**Supplementary Table 6. Drugs linked to the identified proteins associated with dementia, drug type, their potential mechanisms of action, current target disease, and the status and phase of the trials.**

| Drug | Gene | Type | Mechanism Of Action | Disease | Phase | Status |
|---|---|---|---|---|---|---|
| LY-2780301 | *RPS6KB1* | Small molecule | Ribosomal protein S6 kinase (P70S6K) inhibitor | lymphoma | Phase I | Completed |
| LY-2584702 | *RPS6KB1* | Small molecule | Ribosomal protein S6 kinase 1 inhibitor | renal cell carcinoma | Phase I | Terminated |
| LY-2584702 | *RPS6KB1* | Small molecule | Ribosomal protein S6 kinase 1 inhibitor | cancer | Phase I | Completed |
| LY-2584702 | *RPS6KB1* | Small molecule | Ribosomal protein S6 kinase 1 inhibitor | non-small cell lung carcinoma | Phase I | Terminated |
| LY-2584702 | *RPS6KB1* | Small molecule | Ribosomal protein S6 kinase 1 inhibitor | metastasis | Phase I | Terminated |
| XL-418 | *RPS6KB1* | Small molecule | Ribosomal protein S6 kinase (P70S6K) inhibitor | cancer | Phase I | Suspended |
| LY-2584702 | *RPS6KB1* | Small molecule | Ribosomal protein S6 kinase 1 inhibitor | cancer | Phase I | Terminated |
| LY-2584702 | *RPS6KB1* | Small molecule | Ribosomal protein S6 kinase 1 inhibitor | neuroendocrine neoplasm | Phase I | Terminated |
| TAS0612 | *RPS6KB1* | Small molecule | Ribosomal protein S6 kinase (P70S6K) inhibitor | neoplasm | Phase I | Recruiting |
| LY-2780301 | *RPS6KB1* | Small molecule | Ribosomal protein S6 kinase (P70S6K) inhibitor | metastasis | Phase I | Completed |
| MSC-2363318A | *RPS6KB1* | Small molecule | Ribosomal protein S6 kinase (P70S6K) inhibitor | neoplasm | Phase I | Completed |
| MARIMASTAT | *MMP12* | Small molecule | Matrix metalloproteinase 12 inhibitor | lung cancer | Phase III | Completed |
| MARIMASTAT | *MMP12* | Small molecule | Matrix metalloproteinase 12 inhibitor | breast cancer | Phase III | Completed |
| CTS-1027 | *MMP12* | Small molecule | Matrix metalloproteinase 12 inhibitor | hepatitis C virus infection | Phase II | Terminated |
| AZD-1236 | *MMP12* | Small molecule | Matrix metalloproteinase 12 inhibitor | cystic fibrosis | Phase II | Withdrawn |
| AZD-1236 | *MMP12* | Small molecule | Matrix metalloproteinase 12 inhibitor | chronic obstructive pulmonary disease | Phase II | Completed |
| CTS-1027 | *MMP12* | Small molecule | Matrix metalloproteinase 12 inhibitor | hepatitis C virus infection | Phase II | Completed |
| CTS-1027 | *MMP12* | Small molecule | Matrix metalloproteinase 12 inhibitor | chronic hepatitis C virus infection | Phase II | Completed |

**Supplementary Table 7A. International classification of disease codes for definite dementia as recorded in hospital admission and mortality records in ELSA.**

| ICD Code | ICD Description | Dementia Category |
|---|---|---|
| F00 | Dementia in Alzheimer's disease | Definite |
| G30 | Alzheimer's disease | Definite |
| F00.0<br>G30.0 | Dementia in Alzheimer's disease with early onset | Definite |
| F00.1<br>G30.1 | Dementia in Alzheimer disease with late onset | Definite |
| F00.2 | Dementia in Alzheimer disease, atypical or mixed type | Definite |
| G30.8 | Other Alzheimer's disease | Definite |
| F00.9<br>G30.9 | Dementia in Alzheimer's disease, unspecified | Definite |
| F01 | Vascular dementia | Definite |
| F01.0 | Vascular dementia of acute onset | Definite |
| F01.1 | Multi-infarct dementia | Definite |
| F01.2 | Subcortical vascular dementia | Definite |
| F01.3 | Mixed cortical and subcortical vascular dementia | Definite |
| F01.8 | Other vascular dementia | Definite |
| F01.9 | Vascular dementia, unspecified | Definite |
| F02 | Dementia in other diseases classified elsewhere | Definite |
| F02.0 | Dementia in Pick's disease | Definite |
| F02.1 | Dementia in Creutzfeldt-Jakob disease | Definite |
| F02.2 | Dementia in Huntington's disease | Definite |
| F02.3 | Dementia in Parkinson's disease | Definite |
| F02.4 | Dementia in human immunodeficiency virus [HIV] disease | Definite |
| F02.8 | Dementia in other specified diseases classified elsewhere | Definite |
| G31.0 | Frontotemporal dementia | Definite |
| G31.8 | Other specified degenerative diseases of nervous system<br>Grey-matter degeneration [Alpers]<br>Lewy body(ies)(dementia)(disease)<br>Subacute necrotizing encephalopathy [Leigh] | Definite |
| F03 | Unspecified dementia | Definite |
| F05.1 | Delirium superimposed on dementia | Definite |
| I67.3 | Binswanger's disease. Also known as subcortical leukoencephalopathy, is a form of small vessel vascular dementia | Definite |
| F10.7 | Residual and late-onset psychotic disorder: Includes Alcoholic dementia NOS<br>Chronic alcoholic brain syndrome<br>Dementia and other milder forms of persisting impairment of cognitive functions | Definite |
| F04 | Amnestic disorder due to known physiological condition - Korsakov's psychosis or syndrome, non-alcoholic | Definite (listed as dementia by Alzheimer's Association) |
| A81.0 | Creutzfeldt-Jakob disease | Definite (listed as dementia by Alzheimer's Association) |

*References:*

1. *Hayat S, Luben R, Khaw K, et al. Evaluation of routinely collected records for dementia outcomes in UK: a prospective cohort study BMJ Open 2022;12:e060931. doi: 10.1136/bmjopen-2022-060931.*
2. *Wilkinson T, Ly A, Schnier C, Rannikmäe K, Bush K, Brayne C, Quinn TJ, Sudlow CLM. Identifying dementia cases with routinely-collected health data: a systematic review. Alzheimer's & Dementia: The Journal of the Alzheimer's Association.*

**Supplementary Table 7B: International classification of disease codes by dementia sub-type for definite dementia cases in ELSA.**

| Alzheimer's Disease | Vascular dementia |
|---|---|
| F0.0 | F01 |
| F00.0 | F01.0 |
| F00.1 | F01.1 |
| F00.2 | F01.2 |
| F00.9 | F01.3 |
| G30.0 | F01.8 |
| G30 | F01.9 |
| G30.1 | |
| G30.8 | |
| G30.9 | |

**Supplementary Table 8. Summary Information on dementia GWAS datasets.**

| Trait | Late-onset Alzheimer's disease | Alzheimer's disease | Alzheimer's disease (wide definition) | All-cause dementia | Vascular dementia |
|---|---|---|---|---|---|
| **Year** | 2019 | 2022 | 2023 | 2023 | 2023 |
| **Case, n** | 21,982 | 39,106 (clinically diagnosed cases) + 46,828 (proxy cases) | 15,617 | 19,157 | 2,717 |
| **Control, n** | 41,944 | 401,577 | 396,564 | 388,560 | 393,024 |
| **SNP, n** | 10,528,610 | 20,921,626 | 21,306,349 | 21,306,258 | 21,306,039 |
| **Data cohort** | International Genomics of Alzheimer's Project (IGAP consortium): Alzheimer Disease Genetics Consortium (ADGC), European Alzheimer's Disease Initiative (EADI), Cohorts for Heart and Aging Research in Genomic Epidemiology Consortium (CHARGE), Genetic and Environmental Risk in AD/Defining Genetic, Polygenic and Environmental Risk for Alzheimer's Disease Consortium (GERAD/PERADES) | European Alzheimer & Dementia Biobank (EADB) consortium | FinnGen | FinnGen | FinnGen |
| **Country of origin** | Canada, France, Germany, Greece, Iceland, Netherlands, U.K., U.S. | Belgium, Bulgaria, Czech Republic, Denmark, Finland, France, Germany, Greece, Italy, Netherlands, Norway, Portugal, Spain, Sweden, Switzerland, U.K., U.S. | Finland | Finland | Finland |
| **Case ascertainment** | Clinical assessment, magnetic resonance imaging or autopsy-confirmed, and/or diagnosis from health care records | Clinical diagnosis, proxy cases | Clinical diagnosis from health care records, insurance reimbursement records, medication purchase records | Clinical diagnosis from health care records, insurance reimbursement records, medication purchase records | Clinical diagnosis from health care records |
| **Genetic data** | Genome-wide genotyping, imputation using 1000 Genomes project, phase 2 release | Genome-wide genotyping, Affymetrix, Illumina [21101114] imputed | Illumina GWAS arrays, using Finnish specific WGS reference panel of ~9000 individuals | Illumina GWAS arrays, using Finnish specific WGS reference panel of ~9000 individuals | Illumina GWAS arrays, using Finnish specific WGS reference panel of ~9000 individuals |

*GWAS, genome wide association study; SNP, single-nucleotide polymorphism; WGS, whole genome sequence.*

### Supplementary Figures

**Supplementary Figure 1. Flow diagram for ELSA proteomics project sample selection and quality control pipeline.**

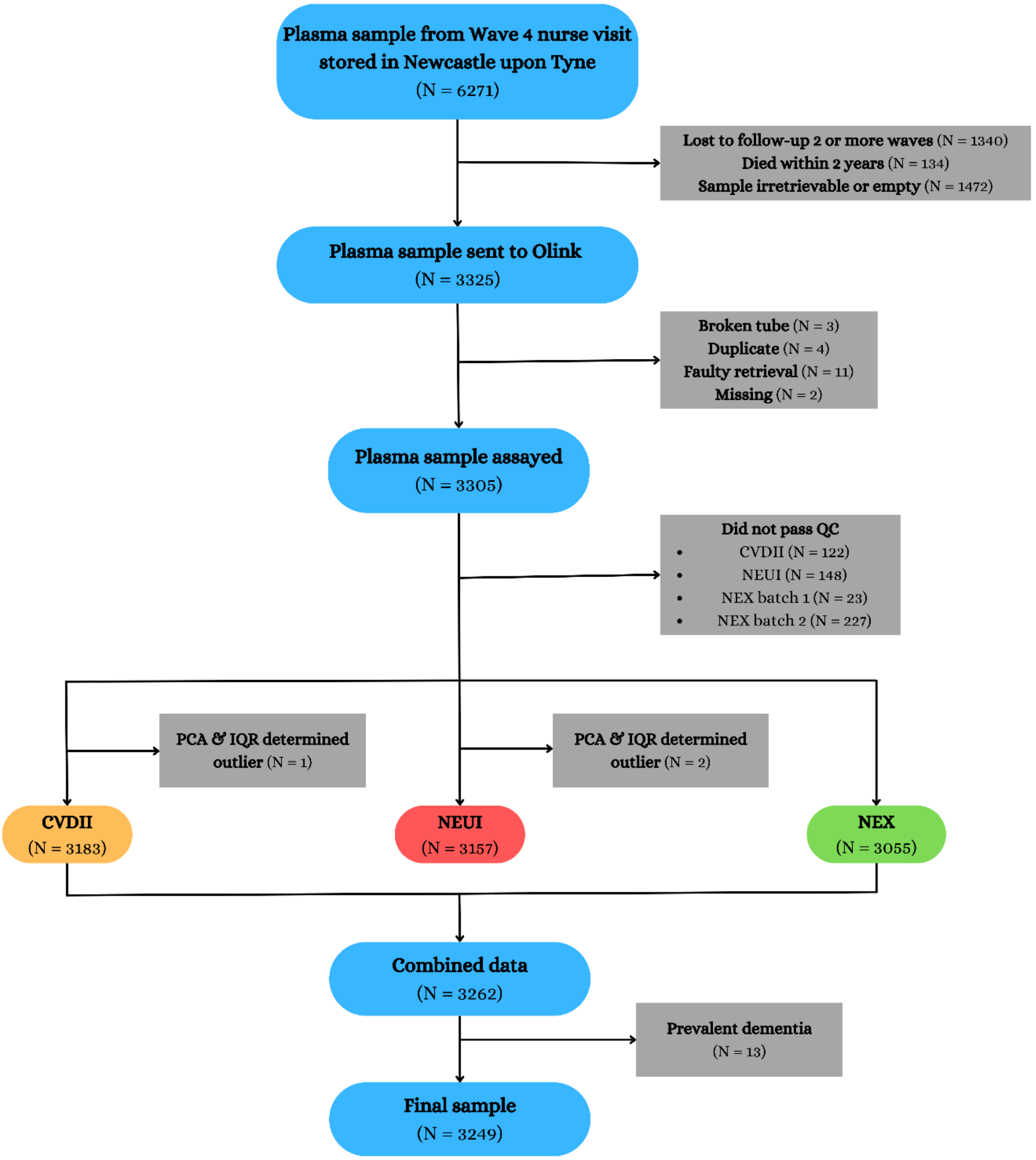


*QC, quality control; PCA, principal component analysis; IQR, interquartile range; CVDII, cardiovascular disease 2 Olink panel; NEU, neurology Olink panel; NEX, neurology exploratory Olink panel.*

**Supplementary Figure 2. Dementia cases by data source in ELSA.**

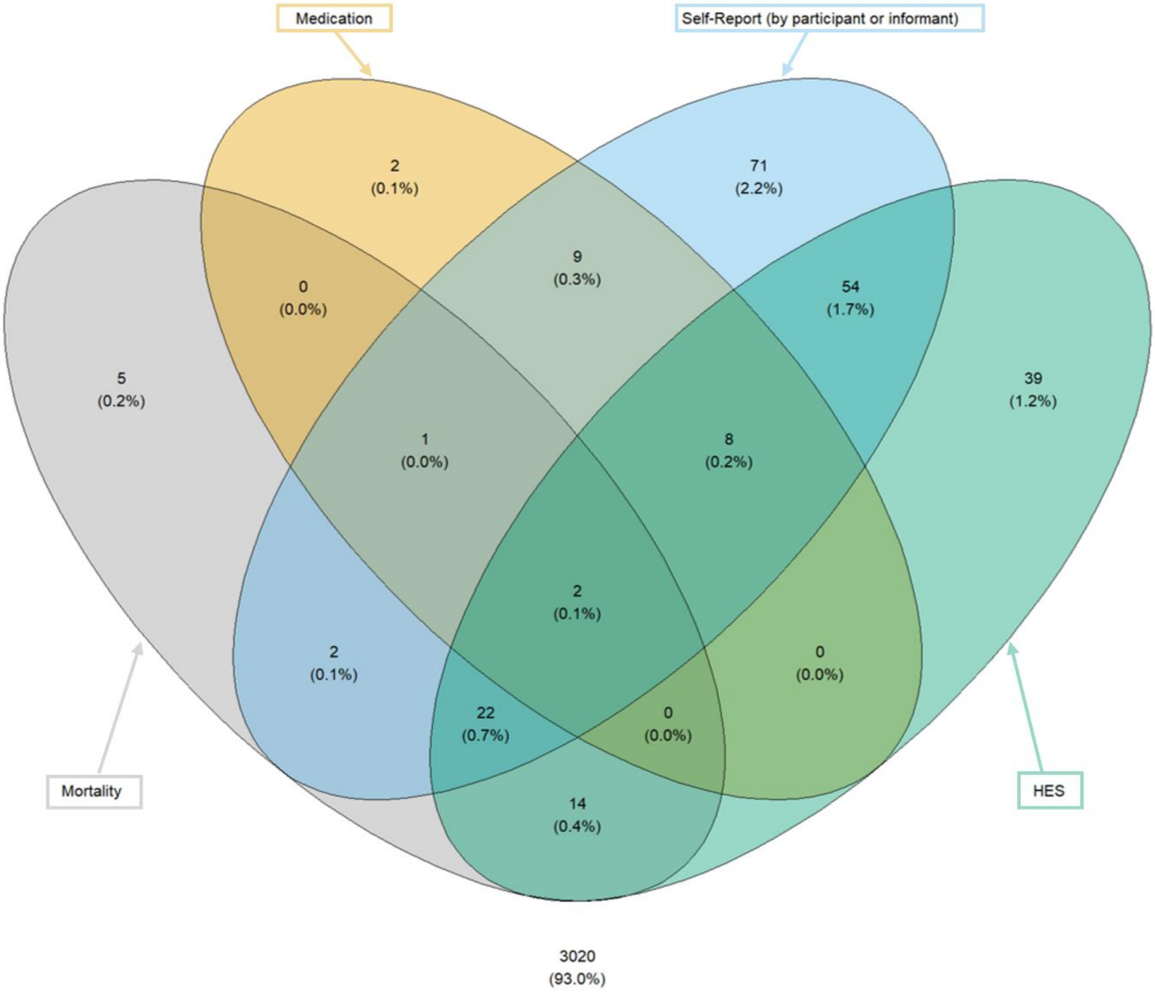


*HES, Hospital Episode Statistics.*

**Supplementary Figure 3. Boxplot for all proteins in ELSA by dementia status.**

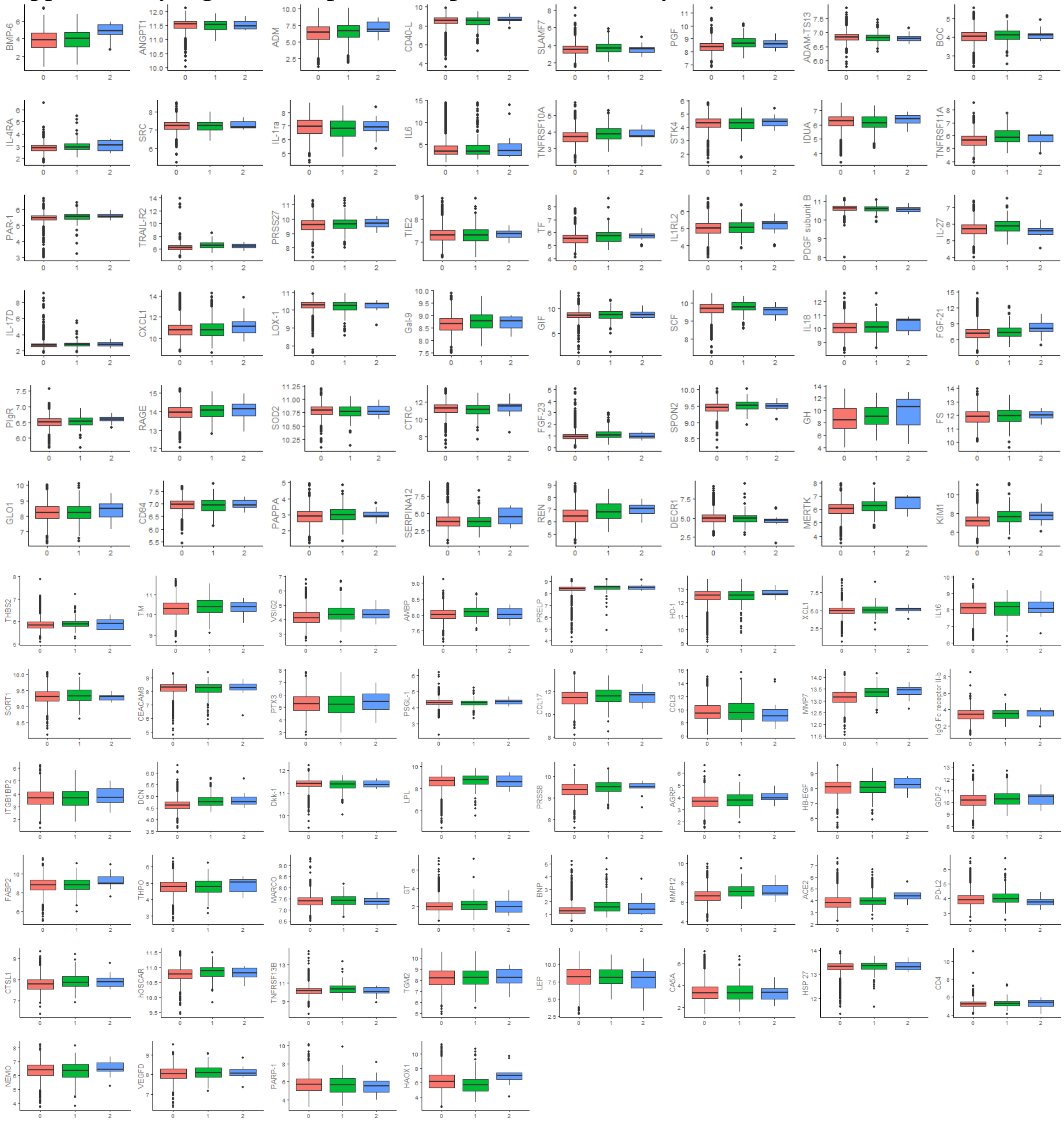

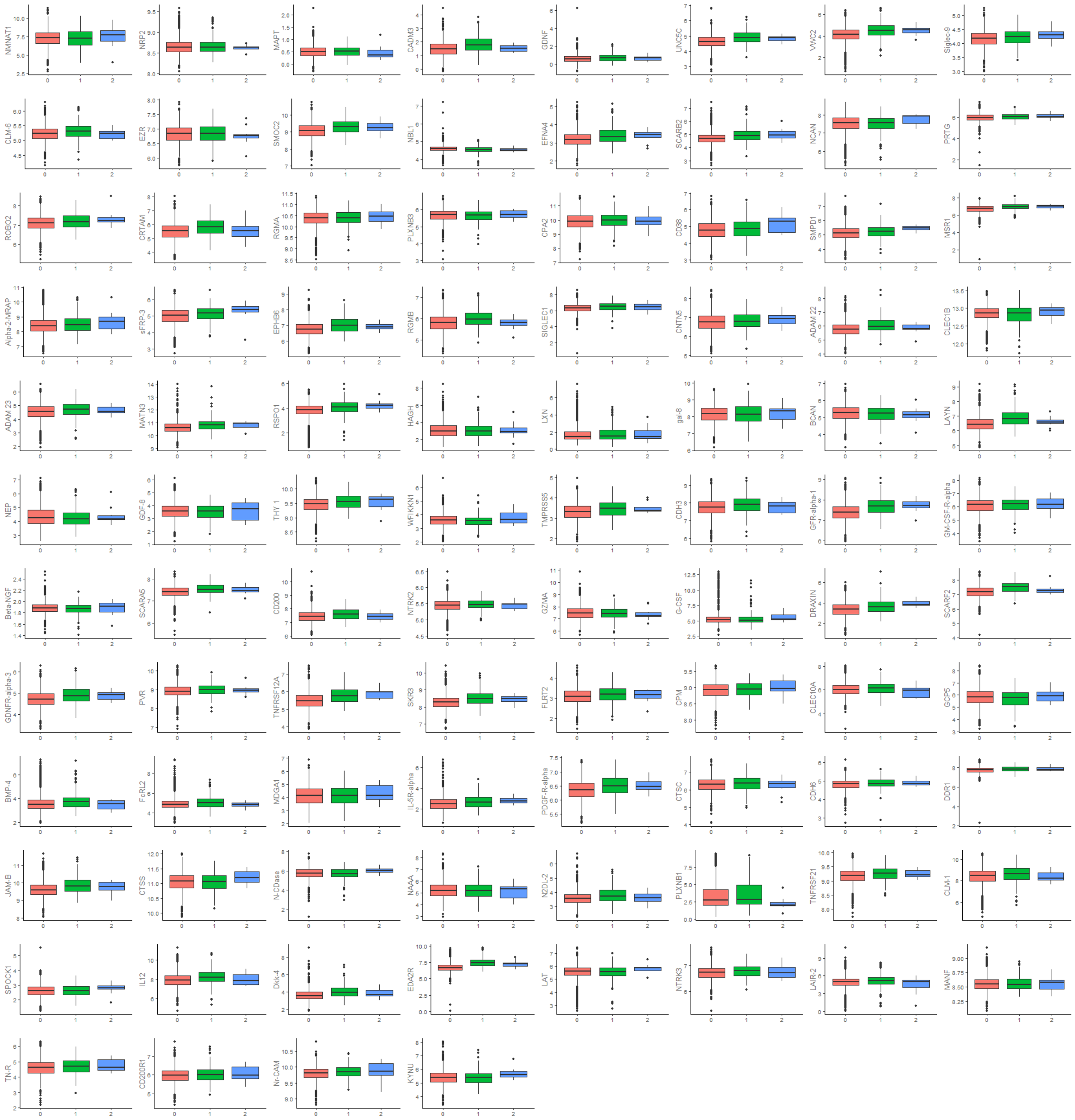
NMNAT1
NRP2
MAPT
CADM3
GDNF
UNC5C
VWC2
Siglec-9
CLM-6
EZR
SMOC2
NBL1
EFNA4
SCARB2
NCAN
PRTG
ROBO2
CRTAM
RGMA
PLXNB3
CPA2
CD38
SMPD1
MSR1
Alpha-2-MRAP
sFRP-3
EPHB6
RGMB
SIGLEC1
CNTN5
ADAM 22
CLEC1B
ADAM 23
MATN3
RSPO1
HAGH
LXN
gal-8
BCAN
LAYN
NEP
GDF-8
THY 1
WFIKKN1
TMPRSS5
CDH3
GFR-alpha-1
GM-CSF-R-alpha
Beta-NGF
SCARA5
CD200
NTRK2
GZMA
G-CSF
DRAXIN
SCARF2
GDNFR-alpha-3
PVR
TNFRSF12A
SKR3
FLRT2
CPM
CLEC10A
GCP5
BMP-4
FcRL2
MDGA1
IL-5R-alpha
PDGF-R-alpha
CTSC
CDH6
DDR1
JAM-B
CTSS
N-CDase
NAAA
N2DL-2
PLXNB1
TNFRSF21
CLM-1
SPOCK1
IL12
Dkk-4
EDA2R
LAT
NTRK3
LAIR-2
MANF
TN-R
CD200R1
Nr-CAM
KYNU

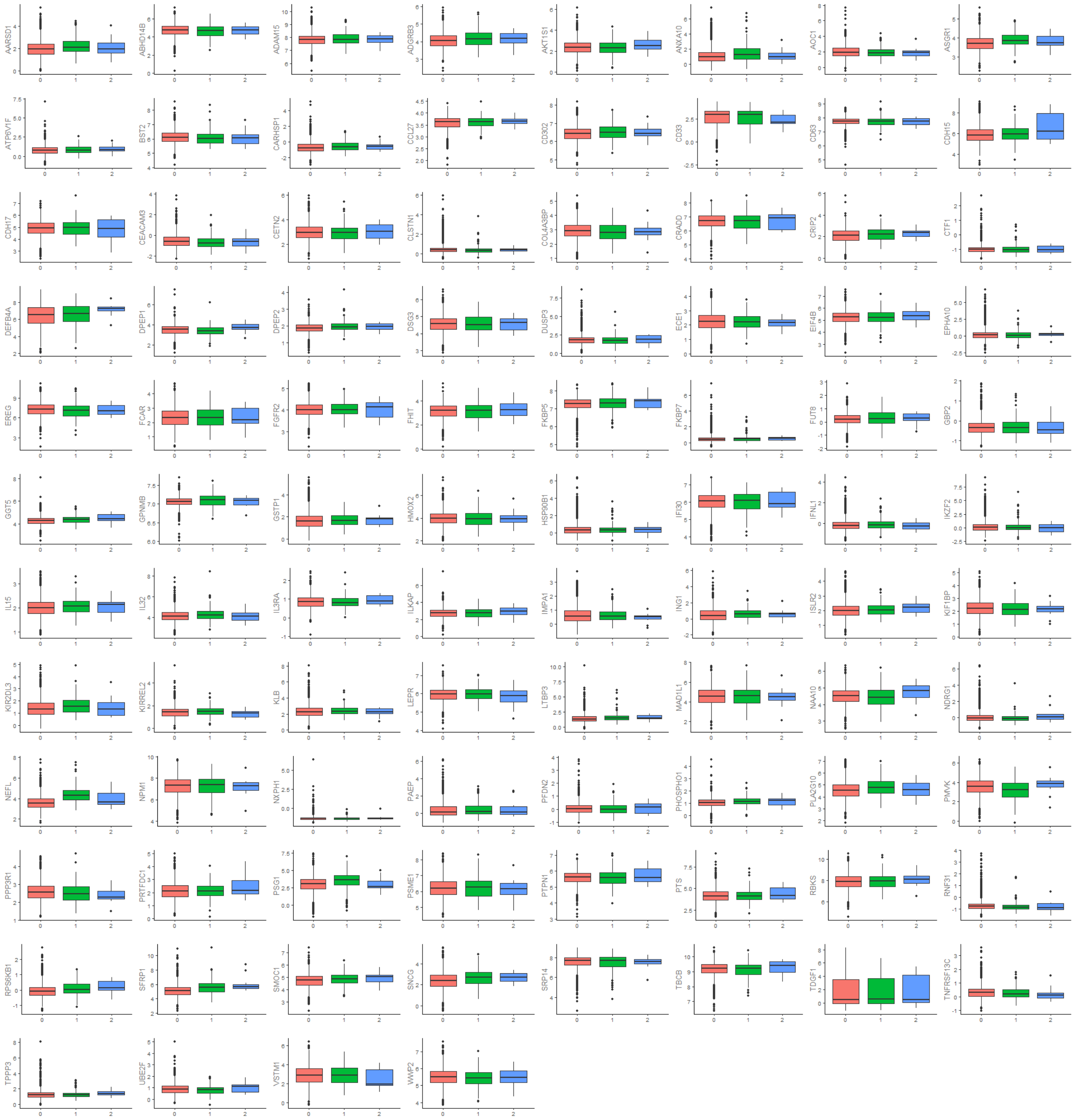


*Red – no dementia; green – incident dementia, blue – prevalent dementia. Y-axis indicates the protein NPX values with error bars.*

**Supplementary Figure 4. Volcano plot showing the unadjusted HR (x axis) and two-sided P values (y axis) for the association between protein concentration with incident all-cause dementia using imputed data.**

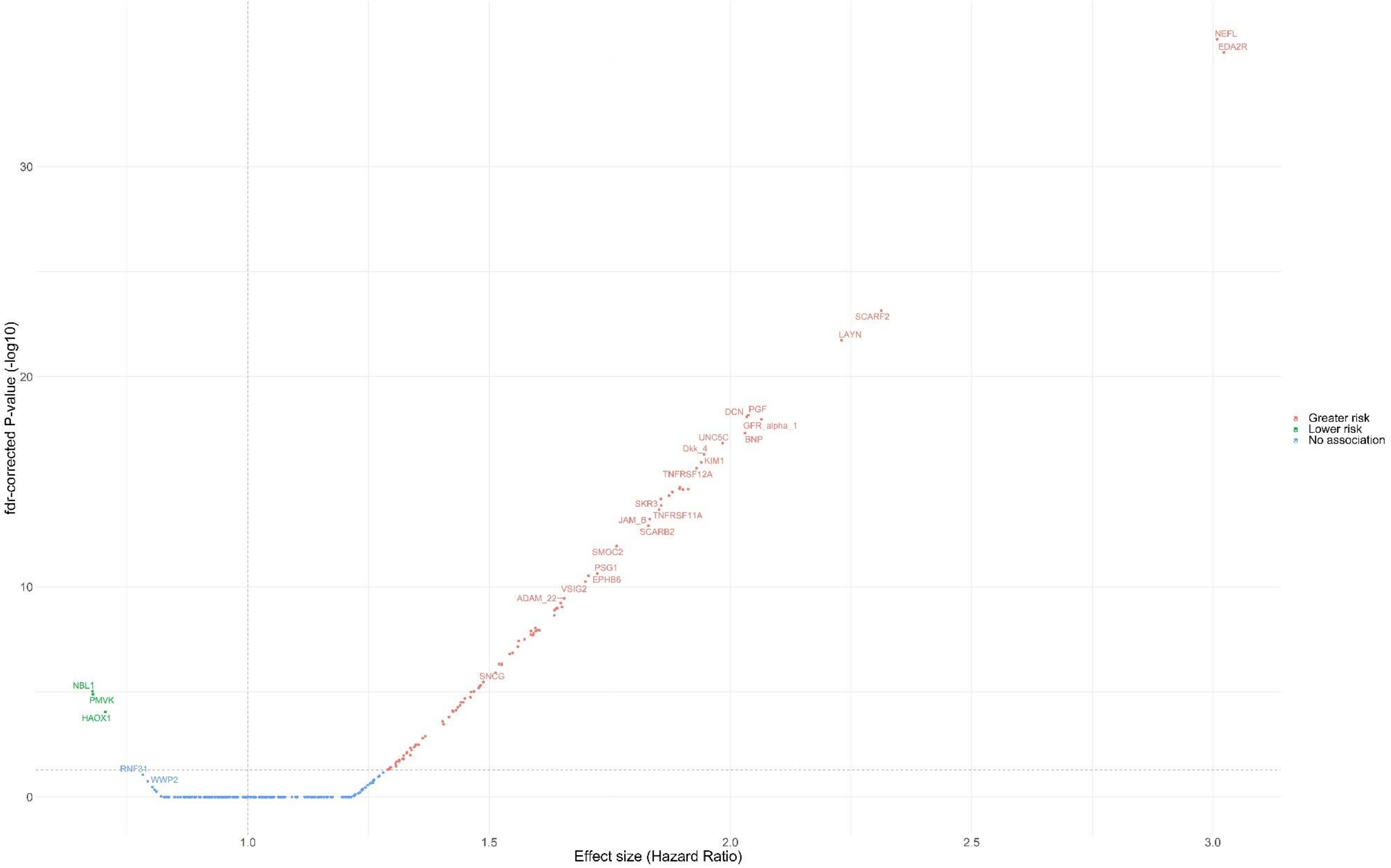


*Proteins above the horizontal dotted grey line were significantly associated with incident dementia FDR-corrected p-value <0.05.*

**Supplementary Figure 5. Volcano plot showing the sex-, age- and ethnicity-adjusted HR (x axis) and two-sided P values (y axis) for the association between protein concentration with incident all-cause dementia using imputed data.**

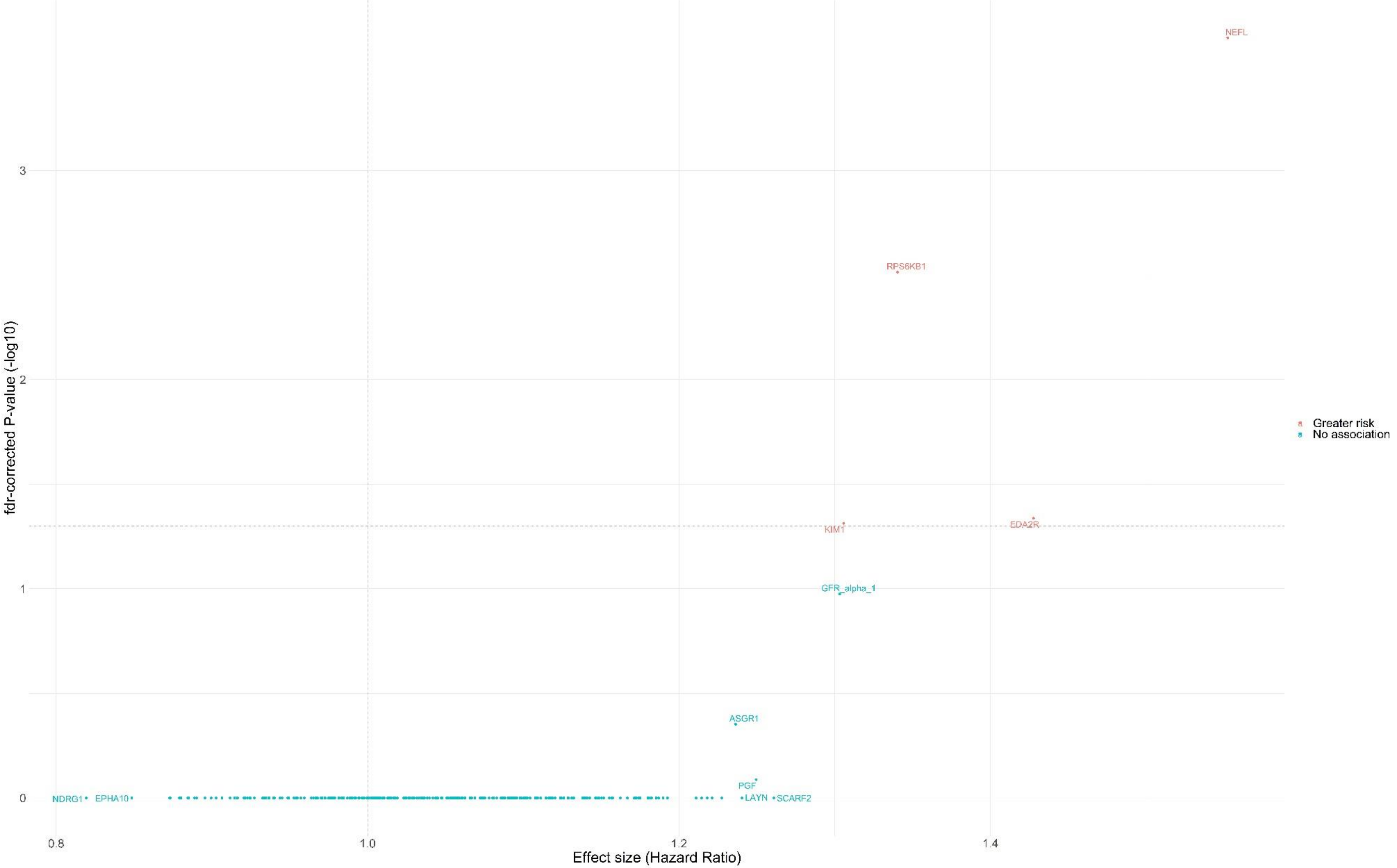


*Proteins above the horizontal dotted grey line were significantly associated with incident dementia FDR-corrected p-value <0.05.*

**Supplementary Figure 6. Volcano plot showing the fully adjusted HR (x axis) and two-sided P values (y axis) for the association between protein concentration with incident all-cause dementia, excluding other ethnic groups using imputed data.**

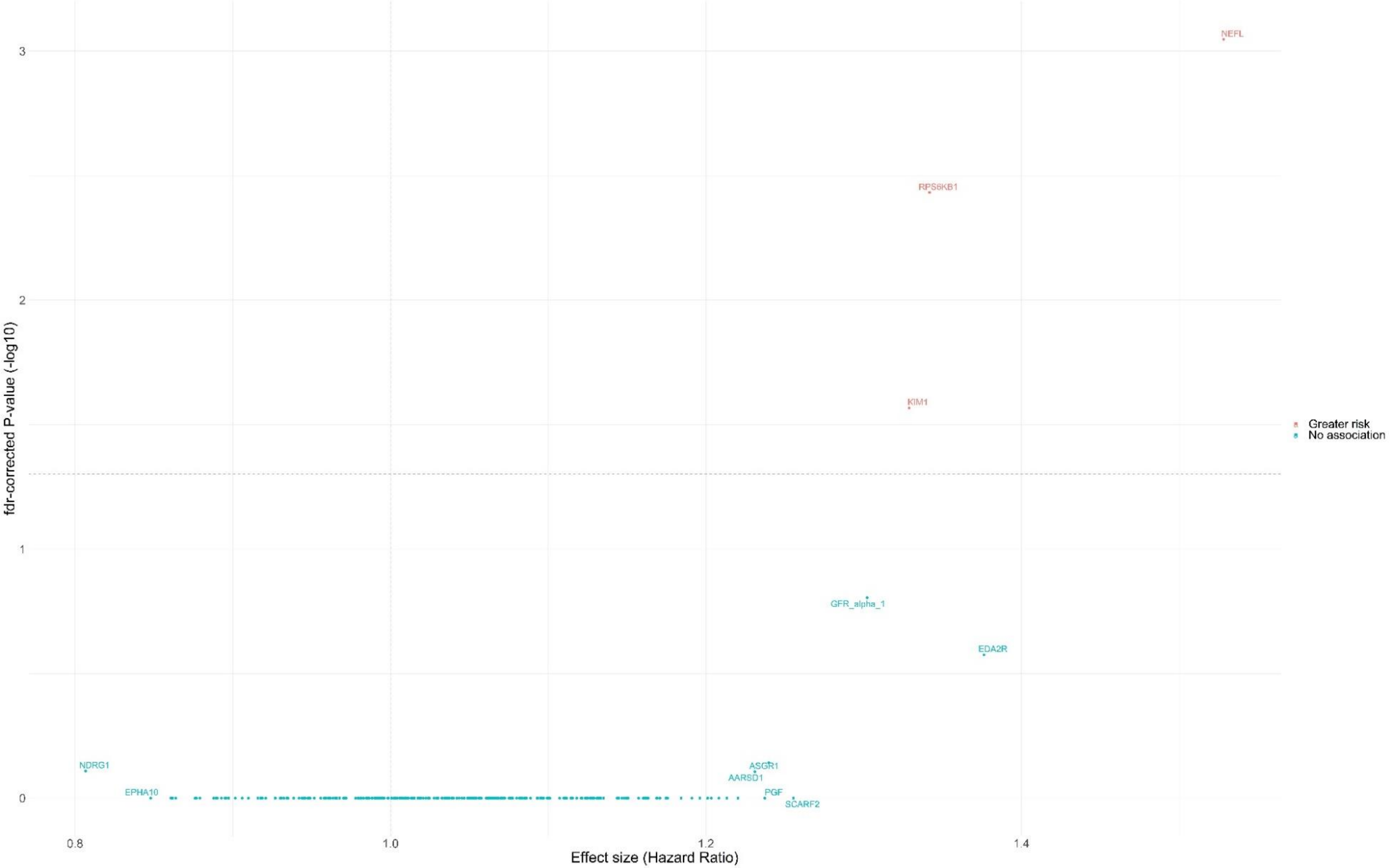


*Adjusted for age, sex, education, ethnicity, smoking status, depression, cardiovascular disease, body mass index, systolic blood pressure, LDL cholesterol. Proteins above the horizontal dotted grey line were significantly associated with incident dementia FDR-corrected p-value <0.05.*

**Supplementary Figure 7. Volcano plot showing the fully adjusted HR (x axis) and two-sided P values (y axis) for the association between protein concentration with incident all-cause dementia, excluding APOE 4 carriers using imputed data.**

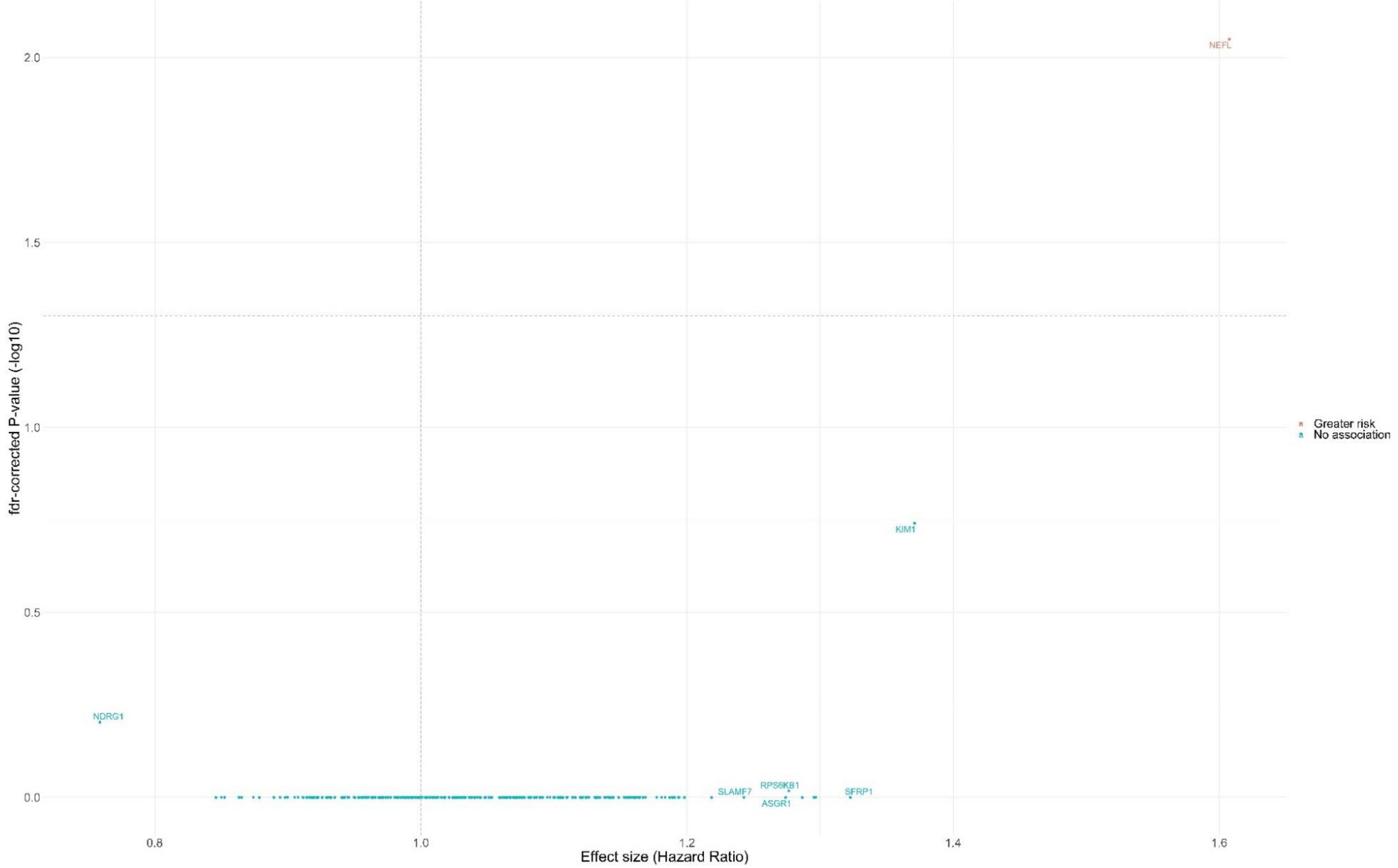


*Adjusted for age, sex, education, ethnicity, smoking status, depression, cardiovascular disease, body mass index, systolic blood pressure, LDL cholesterol. Proteins above the horizontal dotted grey line were significantly associated with incident dementia FDR-corrected p-value <0.05.*

**Supplementary Figure 8. Volcano plot showing the fully adjusted HR (x axis) and two-sided P values (y axis) for the association between protein concentration with incident all-cause dementia, after reducing the possibility of reverse causation bias by excluding all-cause dementia cases that occurred during the first year of follow-up using imputed data.**

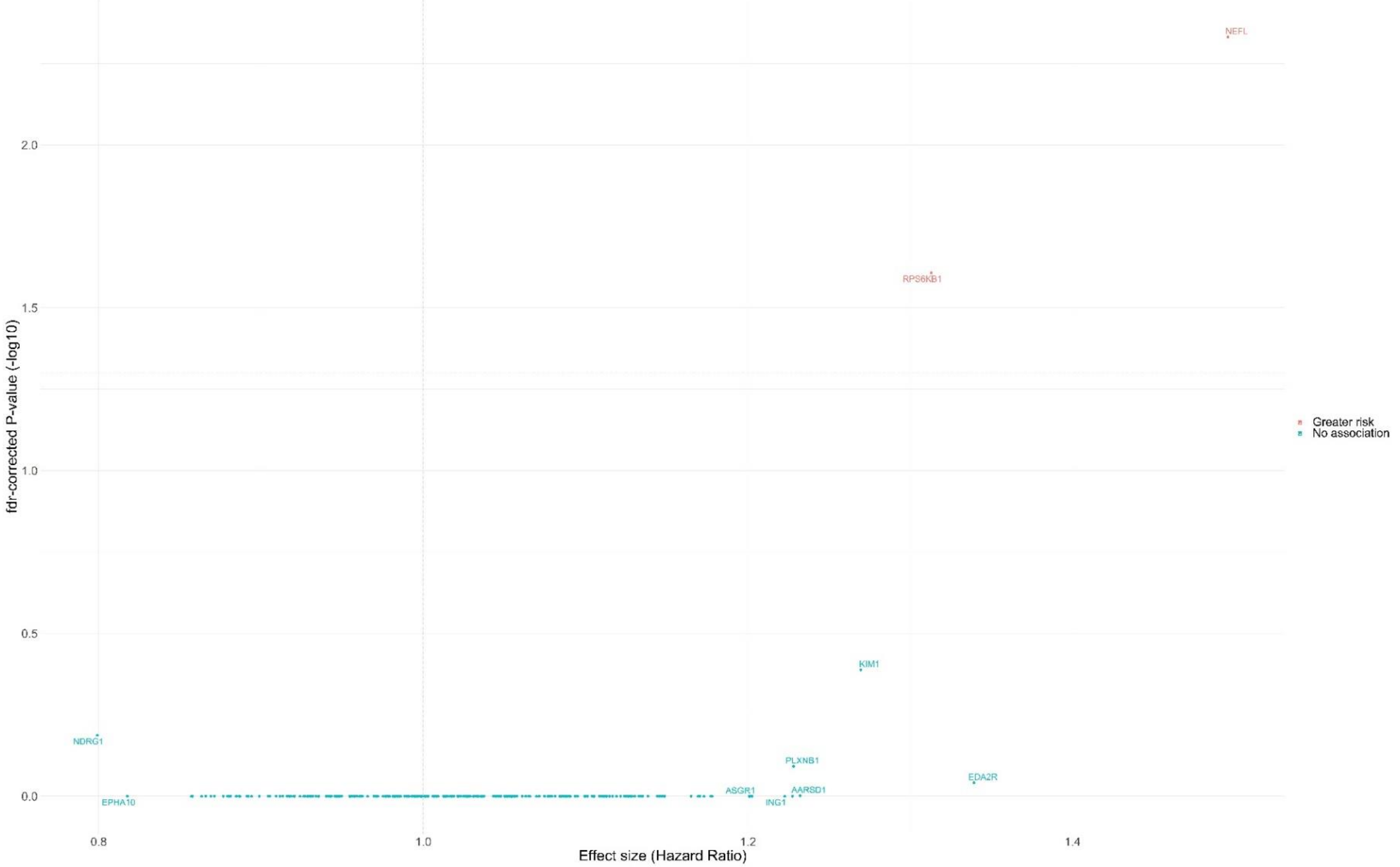


*Adjusted for age, sex, education, ethnicity, smoking status, depression, cardiovascular disease, body mass index, systolic blood pressure, LDL cholesterol. Proteins above the horizontal dotted grey line were significantly associated with incident dementia FDR-corrected p-value < 0.05.*

**Supplementary Figure 9. Volcano plot showing the fully adjusted HR (x axis) and two-sided P values (y axis) for the association between protein concentration with incident all-cause dementia, excluding participants <60 years using imputed data.**

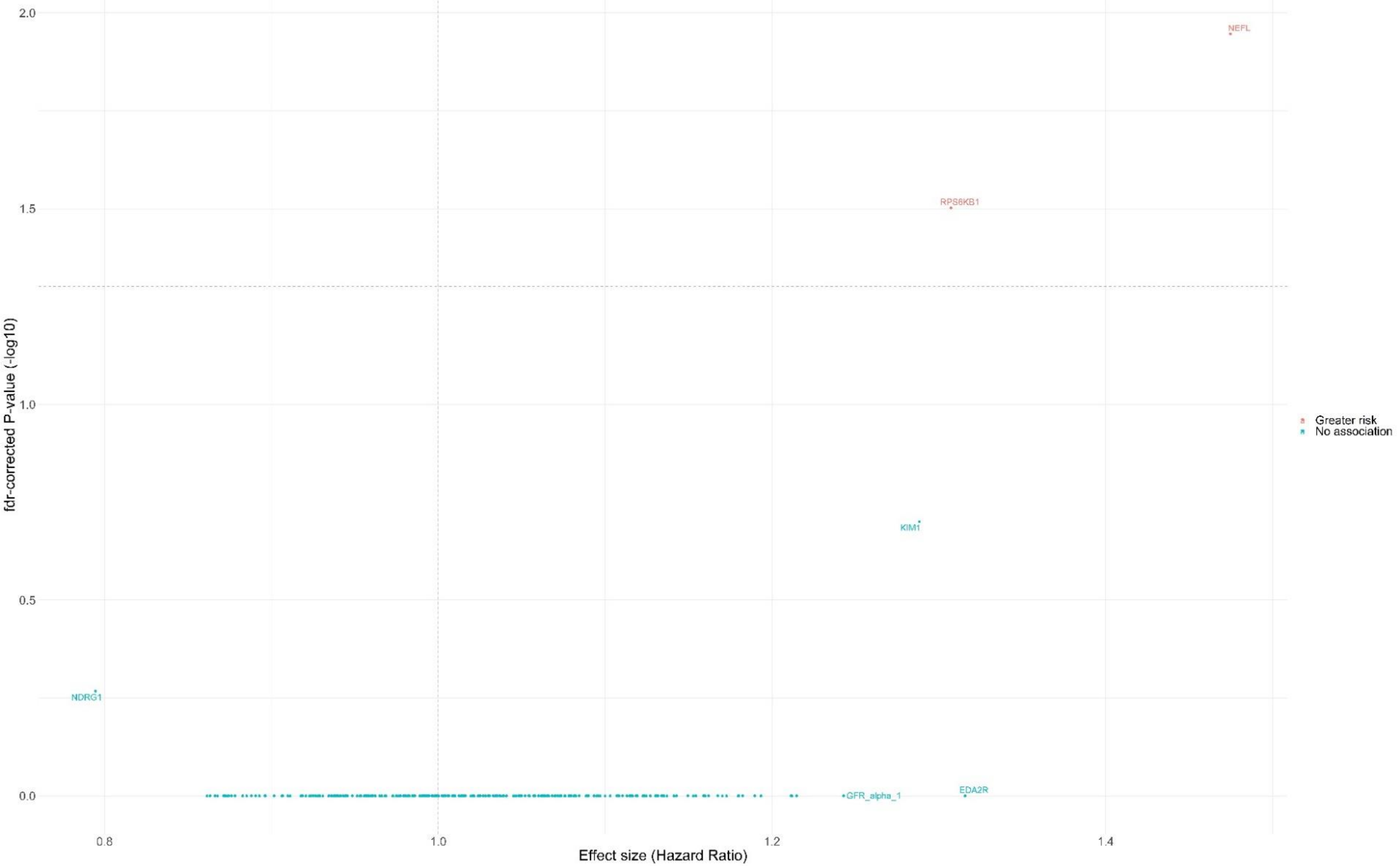


*Adjusted for age, sex, education, ethnicity, smoking status, depression, cardiovascular disease, body mass index, systolic blood pressure, LDL cholesterol. Proteins above the horizontal dotted grey line were significantly associated with incident dementia FDR-corrected p-value < 0.05.*

**Supplementary Figure 10. Volcano plot showing the fully adjusted sub-distribution HR (x axis) and two-sided P values (y axis) for the association between protein concentration with incident all-cause dementia, using Fine-Gray competing risk regression using imputed data.**

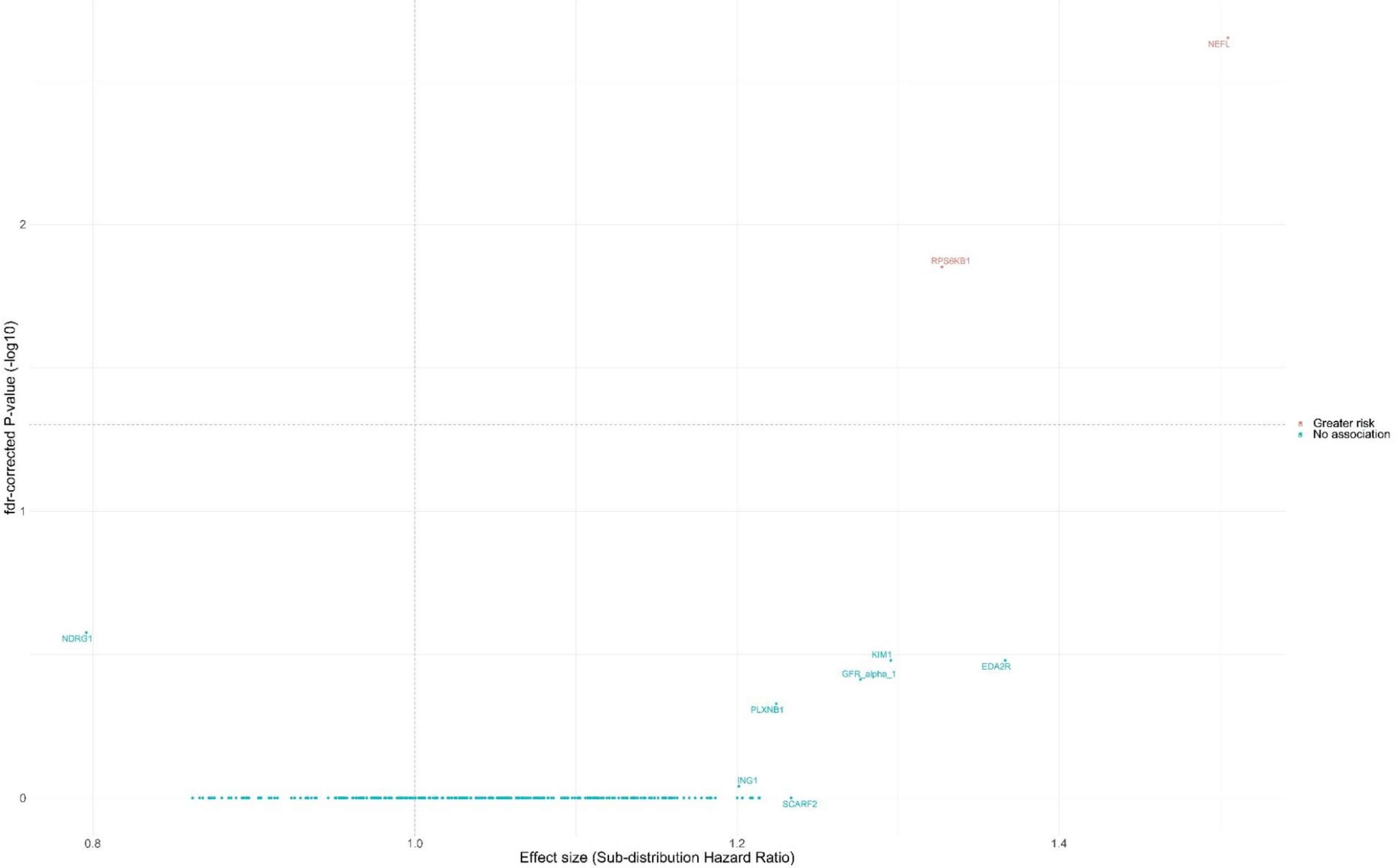


*Adjusted for age, sex, education, ethnicity, smoking status, depression, cardiovascular disease, body mass index, systolic blood pressure, LDL cholesterol. Proteins above the horizontal dotted grey line were significantly associated with incident dementia FDR-corrected p-value < 0.05.*

**Supplementary Figure 11. Volcano plot showing the fully adjusted HR (x axis) and two-sided P values (y axis) for the association between protein concentration with incident Alzheimer's disease using imputed data.**

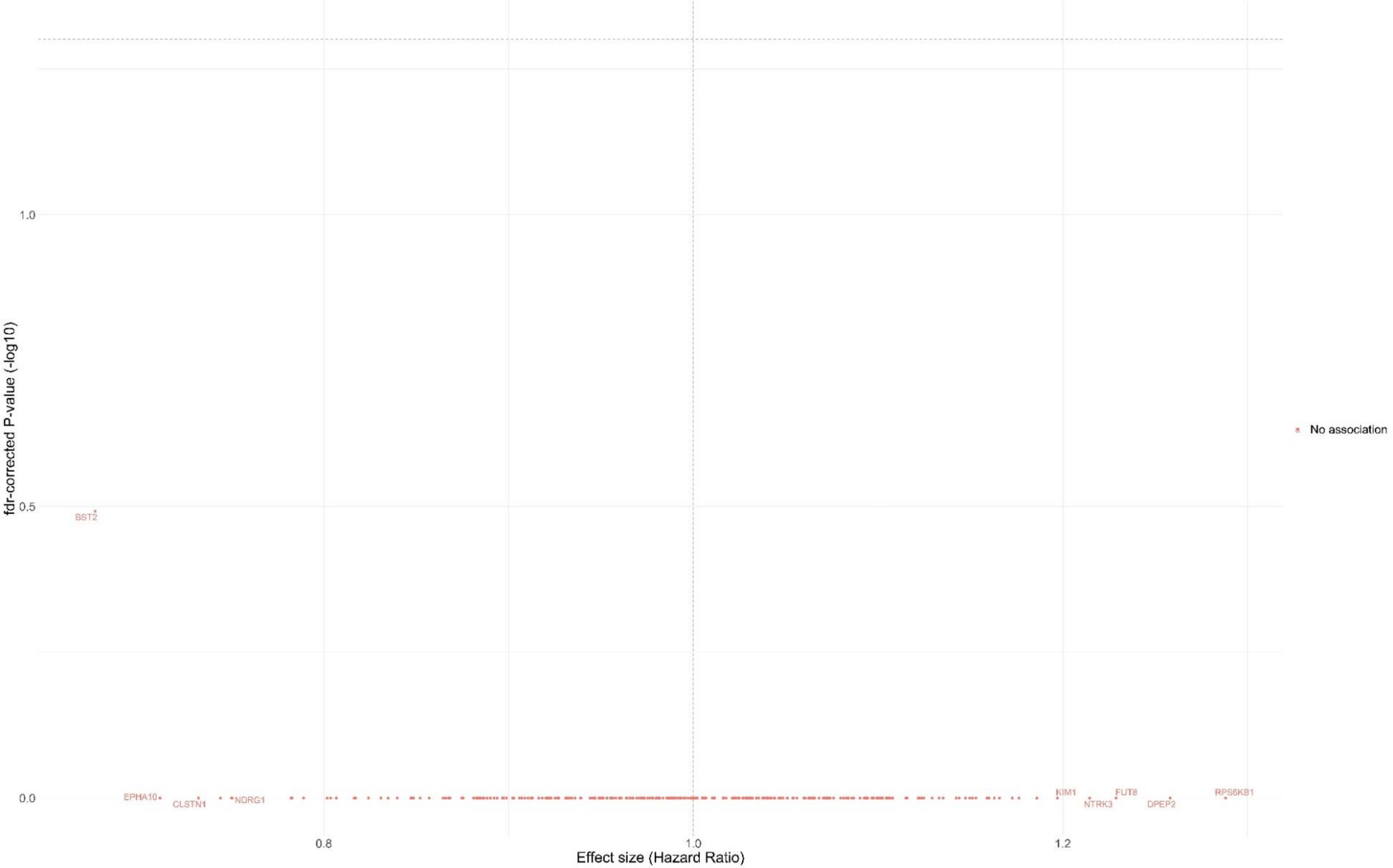


*Adjusted for age, sex, education, ethnicity, smoking status, depression, cardiovascular disease, body mass index, systolic blood pressure, LDL cholesterol. Proteins above the horizontal dotted grey line were significantly associated with incident dementia FDR-corrected p-value < 0.05.*

**Supplementary Figure 12. Volcano plot showing the fully adjusted HR (x axis) and two-sided P values (y axis) for the association between protein concentration with incident vascular dementia using imputed data.**

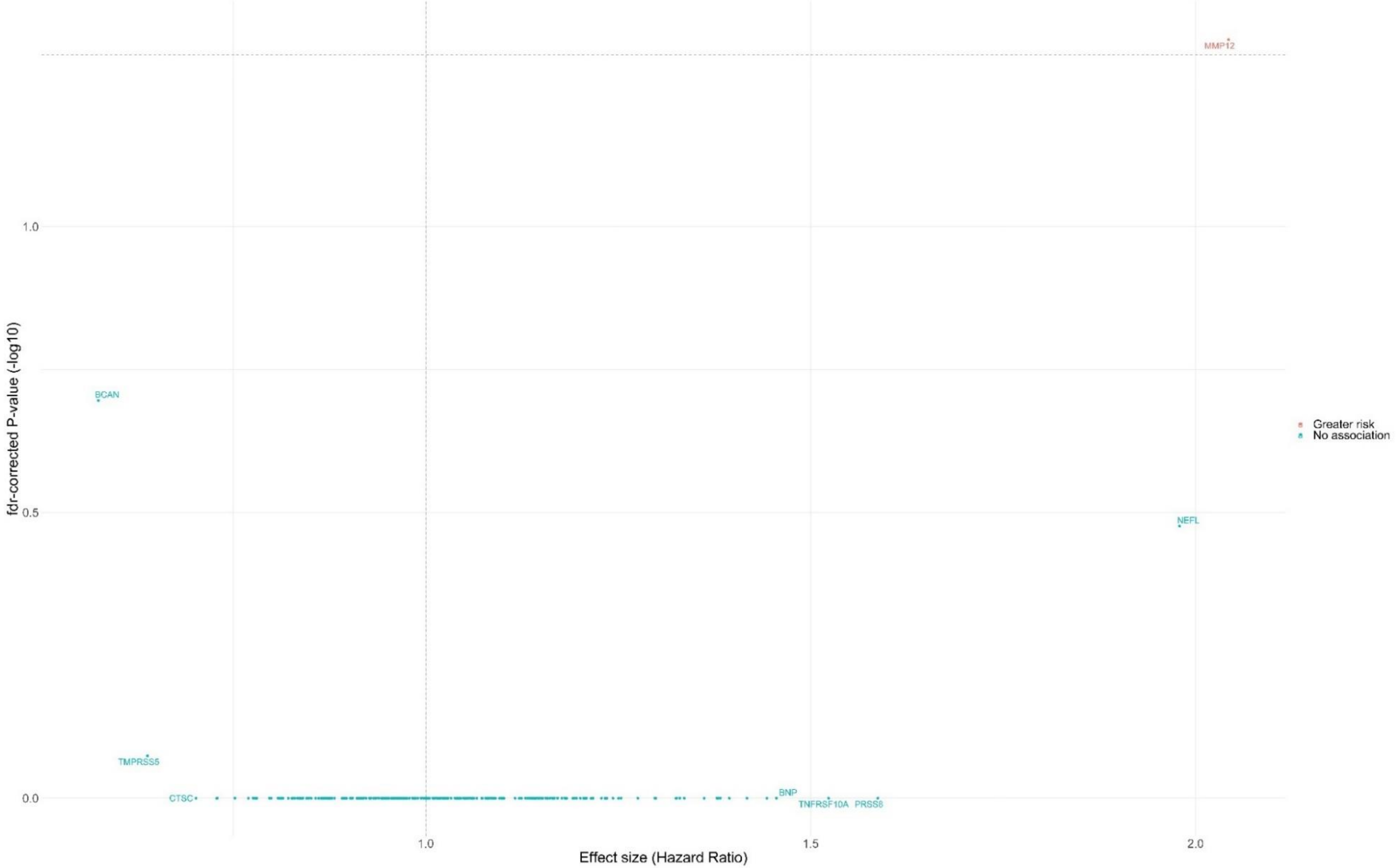


*Adjusted for age, sex, education, ethnicity, smoking status, depression, cardiovascular disease, body mass index, systolic blood pressure, LDL cholesterol. Proteins above the horizontal dotted grey line were significantly associated with incident dementia FDR-corrected p-value < 0.05.*
